# Efficacy and Safety of Sodium-Glucose Co-Transporter 2 Inhibitors in Heart Failure: A Systematic Review and Meta-Analysis of 59 Randomized Controlled Trials

**DOI:** 10.64898/2026.01.28.26345101

**Authors:** Vicky Muller Ferreira, Victor Ayres Muller

## Abstract

**Background:** Sodium-glucose co-transporter 2 (SGLT2) inhibitors have emerged as a cornerstone of heart failure (HF) therapy, yet the totality of randomized evidence — including smaller trials — has not been comprehensively synthesized. We aimed to evaluate the efficacy and safety of SGLT2 inhibitors across the full spectrum of HF.

**Methods:** We searched PubMed, Cochrane CENTRAL, ClinicalTrials.gov, and WHO ICTRP from inception to March 2026 for randomized controlled trials comparing any SGLT2 inhibitor with placebo or standard care in adults with HF. Primary outcomes were all-cause mortality (ACM) and HF hospitalization (HFH). We used random-effects models with Mantel-Haenszel risk ratios and Hartung-Knapp-Sidik-Jonkman confidence intervals.

Certainty of evidence was assessed using GRADE. The protocol was registered prospectively (PROSPERO CRD420251167908).

**Results:** Of 6,239 records identified, 114 studies met inclusion criteria and 59 RCTs (29,692 participants) were included in quantitative synthesis. SGLT2 inhibitors significantly reduced ACM (RR 0.90 [0.83, 0.98], p = 0.016; 26 trials; I^2^ = 0%; low certainty) and HFH (RR 0.74 [0.69, 0.79], p < 0.001; 15 trials; I^2^ = 0%; moderate certainty). The composite of CVD and HFH was reduced (RR 0.80 [0.75, 0.85], p < 0.001; high certainty). Genital infections were significantly increased (RR 3.75 [1.72, 8.19], p = 0.007). Results were robust across 12 sensitivity analyses and 4 alternative statistical models.

**Conclusions:** SGLT2 inhibitors reduce all-cause mortality, HF hospitalization, cardiovascular death, and serious adverse events in adults with HF, with an acceptable safety profile apart from increased genital infections. These findings support the use of SGLT2 inhibitors as a foundational therapy across the HF spectrum.

## Introduction

Heart failure (HF) affects over 64 million people worldwide and remains a leading cause of hospitalization and death, with five-year mortality rates approaching 50% despite advances in pharmacotherapy (1,2). The economic burden is substantial, with global costs exceeding $100 billion annually, driven largely by recurrent hospitalizations (1). Although guideline-directed medical therapy — including renin-angiotensin-aldosterone system inhibitors, beta-blockers, and mineralocorticoid receptor antagonists — has improved outcomes in HF with reduced ejection fraction (HFrEF), progress in HF with preserved ejection fraction (HFpEF) has been more limited (3).

Sodium-glucose co-transporter 2 (SGLT2) inhibitors, originally developed as glucose-lowering agents for type 2 diabetes, have demonstrated unexpected cardiovascular benefits that extend beyond glycemic control (4). The DAPA-HF and EMPEROR-Reduced trials established dapagliflozin and empagliflozin as effective therapies for HFrEF (5,6), while DELIVER and EMPEROR-Preserved expanded these benefits to HFpEF (7,8). Sotagliflozin demonstrated benefits in patients with diabetes and recent worsening HF (9), while empagliflozin improved outcomes when initiated during hospitalization for acute HF (10). Dapagliflozin also improved symptoms and functional status in HFpEF in the PRESERVED-HF trial (11). These landmark trials led to the inclusion of SGLT2 inhibitors as a foundational pillar of HF therapy in both the 2022 AHA/ACC/HFSA and 2023 ESC guidelines, applicable across ejection fraction categories (3,12).

Several meta-analyses have synthesized evidence from the major SGLT2 inhibitor trials in HF (13–15). However, most have focused on a limited number of large, pivotal trials and have not incorporated the growing body of smaller randomized controlled trials (RCTs) that collectively enroll thousands of patients and provide additional data on safety outcomes, quality-of-life measures, and specific patient populations. Furthermore, the landscape continues to evolve, with new trials published through early 2026 that have not been captured in prior syntheses.

The objective of this systematic review and meta-analysis was to comprehensively evaluate the efficacy and safety of SGLT2 inhibitors compared with placebo or standard care in adults with HF, incorporating all available RCTs regardless of sample size, drug, or HF subtype. We aimed to (1) estimate the pooled effects on all-cause mortality, HF hospitalization, cardiovascular death, and other clinical endpoints; (2) explore sources of heterogeneity through subgroup analyses and meta-regression; and (3) assess the certainty of evidence using the GRADE framework.

## Methods

### Protocol and Registration

This systematic review was conducted in accordance with the Preferred Reporting Items for Systematic Reviews and Meta-Analyses (PRISMA) 2020 guidelines (16) and was registered prospectively with PROSPERO (CRD420251167908). The protocol, including all amendments, is available in the supplementary materials.

### Eligibility Criteria

We included RCTs that enrolled adults (age 18 years or older) with a clinical diagnosis of HF of any type — HFrEF (LVEF less than 40%), HFmrEF (LVEF 40–49%), HFpEF (LVEF 50% or greater), or unspecified — and randomized participants to any SGLT2 inhibitor (empagliflozin, dapagliflozin, canagliflozin, sotagliflozin, ertugliflozin, ipragliflozin, luseogliflozin, tofogliflozin, remogliflozin, bexagliflozin, henagliflozin, or licogliflozin) versus placebo or standard care. We excluded observational studies, single-arm trials, studies enrolling exclusively pediatric populations, studies with active comparators, and sub-analyses of already-included trials. No language or date restrictions were applied.

### Information Sources and Search Strategy

We searched PubMed, Cochrane CENTRAL, ClinicalTrials.gov, and WHO ICTRP from inception through March 3, 2026. The search strategy combined three concept blocks: SGLT2 inhibitors (MeSH terms, pharmacological action terms, and free-text synonyms for all individual drugs plus the class wildcard “gliflozin*”), heart failure (MeSH terms and free-text variants), and study design (RCT filter for PubMed; no design filter for CENTRAL or trial registries). Complete search strategies for each database are provided in the supplementary materials.

### Study Selection

Records were managed through a multi-phase screening process. After automated deduplication using exact identifier matching (PMID, DOI, NCT ID) and fuzzy title matching (threshold 95%), title and abstract screening was performed using a four-tier exclusion hierarchy (animal/in vitro, wrong intervention, wrong population, wrong design), with key trial protection to prevent false exclusion of sentinel studies. Full-text screening used an eight-pass pipeline incorporating regex-based screening, API enrichment (PubMed, ClinicalTrials.gov), AI-assisted manual review, trial clustering to identify sub-analyses, block verification, and full-text retrieval with PICO verification. All screening decisions, exclusion codes, and the complete pipeline are documented and reproducible (17).

### Data Extraction

Data were extracted by reading the full-text PDFs of all included studies using a standardized form with approximately 175 fields. Extraction was performed in a structured long format (record ID, field name, value) and subsequently merged into a wide-format analysis dataset. Extracted variables included study design, population characteristics, intervention details, binary outcomes (events and denominators per arm), time-to-event data (hazard ratios with 95% CIs), continuous outcomes (between-group mean differences with 95% CIs), and safety outcomes.

When studies reported medians with interquartile ranges rather than means with standard deviations, we applied the Wan estimator for conversion (18). Standard deviations were derived from standard errors, confidence intervals, or interquartile ranges when necessary. All imputations were documented.

### Risk of Bias Assessment

Risk of bias was assessed for all 59 meta-analysis-included studies using the Cochrane Risk of Bias 2.0 (RoB 2.0) tool (19), evaluating five domains: bias arising from the randomization process (D1), bias due to deviations from intended interventions (D2), bias due to missing outcome data (D3), bias in measurement of the outcome (D4), and bias in selection of the reported result (D5).

### Statistical Analysis

For binary outcomes, we calculated risk ratios (RR) using the Mantel-Haenszel method. As a sensitivity analysis, hazard ratios (HR) were pooled from studies reporting time-to-event data using generic inverse-variance methods. For continuous outcomes, we calculated mean differences (MD). All analyses used random-effects models with restricted maximum likelihood (REML) estimation and Hartung-Knapp-Sidik-Jonkman (HKSJ) confidence intervals (20,21). Heterogeneity was assessed using the Cochran Q test, I^2^ statistic (22), τ^2^, and 95% prediction intervals (when three or more studies were available).

Pre-specified subgroup analyses were performed by HF type, drug class, blinding status, follow-up duration, sample size, risk of bias, and publication era. Twelve filter-based sensitivity analyses and four alternative statistical models (fixed-effect MH-RR, random-effects OR, fixed-effect OR, Peto OR) were examined. Leave-one-out analysis and influence diagnostics were also performed. Meta-regression was conducted when 10 or more studies were available.

Publication bias was assessed using funnel plots (k ≥ 3), Egger’s regression test (23) (k ≥ 10), and the Duval and Tweedie trim-and-fill method (24). Cumulative meta-analysis was conducted for all binary outcomes. Trial sequential analysis (TSA) was performed for primary outcomes at relative risk reduction thresholds of 15% and 20% (25).

The certainty of evidence was assessed using GRADE (26). Number needed to treat (NNT) was calculated as the inverse of the absolute risk reduction.

All analyses were performed in R version 4.5.2 using the meta, metafor, dmetar, and RTSA packages (27,28).

## Results

### Study Selection

The search identified 6,239 records across four databases: PubMed (n = 2,243), Cochrane CENTRAL (n = 2,064), ClinicalTrials.gov (n = 1,637), and WHO ICTRP (n = 295). After removing 1,728 duplicates, 4,511 unique records were screened by title and abstract, of which 2,859 were excluded. Full-text assessment of 1,652 reports led to the exclusion of 1,538 reports, with 114 studies meeting inclusion criteria. Of these, 55 were excluded from quantitative synthesis (19 duplicate registrations, 19 sub-analyses of included trials, 7 registry-only records without published results, 6 conference abstracts, 3 non-RCTs, and 1 study with suspected data fabrication), leaving 59 RCTs in the meta-analysis (Figure 1).

**Figure 1.**
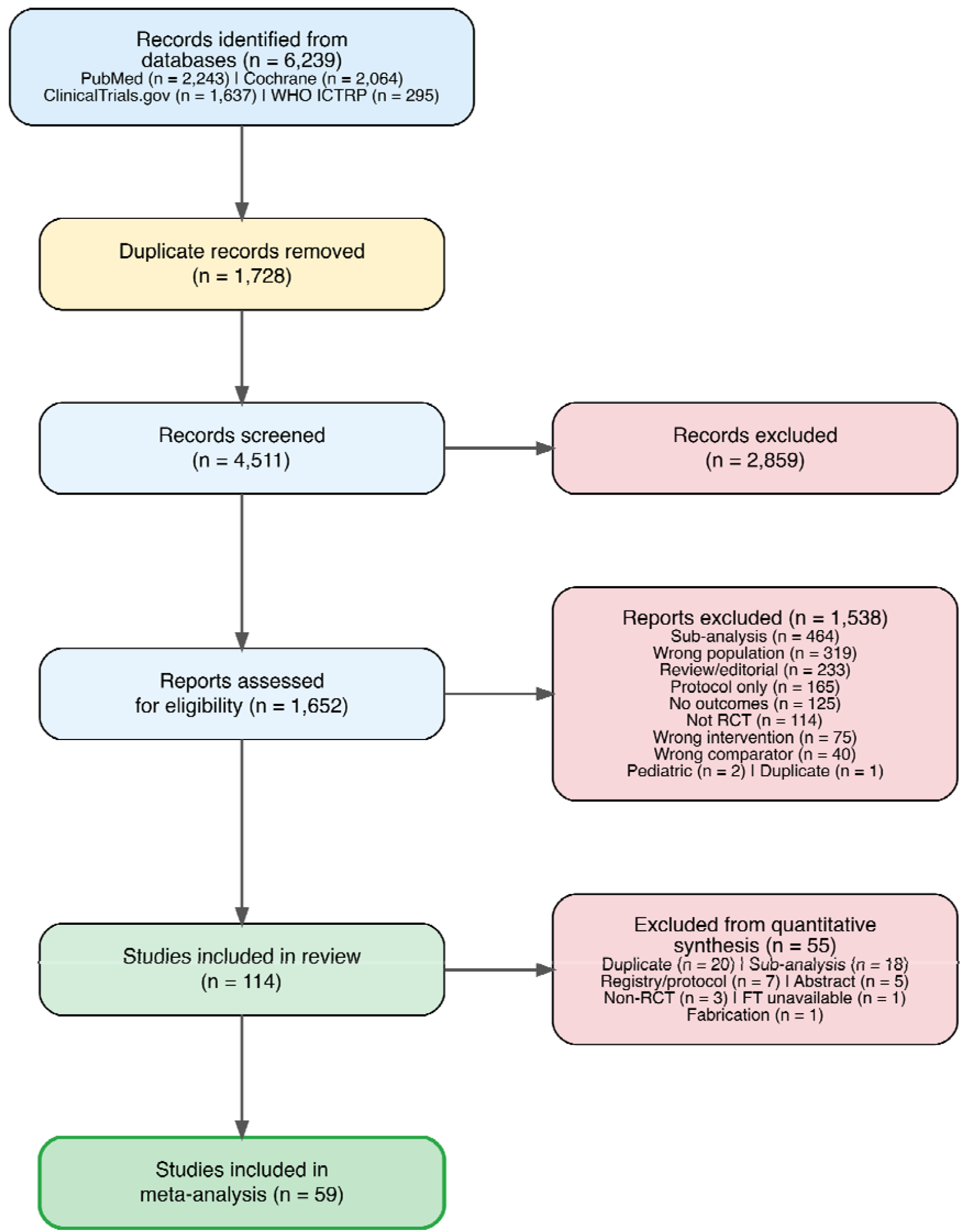
PRISMA 2020 flow diagram.

### Study Characteristics

The 59 included RCTs randomized a total of 29,692 participants (14,848 to SGLT2 inhibitors, 14,805 to control) across 30 countries, published between 2018 and 2026. Empagliflozin was investigated in 25 trials (42%), dapagliflozin in 24 (41%), canagliflozin in 3, sotagliflozin in 1, and other SGLT2 inhibitors in 6. With respect to HF subtype, 17 trials (29%) enrolled exclusively HFrEF patients, 10 (17%) HFpEF, 3 (5%) HFmrEF, and 29 (49%) enrolled mixed populations or did not specify HF type. 36 trials (61%) were double-blind, 22 (37%) were open-label, and 1 was single-blind. The median follow-up was 13 weeks (range 1 to 120 weeks). The median of mean age across trials was 68 years, with a median female proportion of 33%. The median of mean LVEF was 37.2% (Table 1).

**Table 1.** Summary of Study Characteristics.

| Characteristic | Value |
| --- | --- |
| Number of RCTs | 59 |
| Total participants | 29,692 |
| Drug: Empagliflozin | 25 (42%) |
| Drug: Dapagliflozin | 24 (41%) |
| Drug: Canagliflozin | 3 |
| Drug: Sotagliflozin | 1 |
| Drug: Other | 6 |
| HF type: HFrEF | 17 (29%) |
| HF type: HFpEF | 10 (17%) |
| HF type: HFmrEF | 3 (5%) |
| HF type: Mixed/NR | 29 (49%) |
| Design: Double-blind | 36 (61%) |
| Design: Open-label | 22 (37%) |
| Design: Single-blind | 1 |
| RoB: Low | 16 (27%) |
| RoB: Some concerns | 22 (37%) |
| RoB: High | 21 (36%) |
| Median follow-up, weeks | 13 (range 1-120) |
| Median age, years | 68 |
| Median female % | 33 |
| Median LVEF % | 37 |

### Risk of Bias

Among the 59 included RCTs, 16 (27%) were assessed as low risk of bias overall, 22 (37%) as some concerns, and 21 (36%) as high risk of bias. The most common sources of concern were domain D2 (deviations from intended interventions), reflecting the high proportion of open-label trials, and domain D5 (selection of reported results), particularly in smaller single-center studies lacking prospective registration or statistical analysis plans.

### Primary Outcomes

#### All-Cause Mortality

All-cause mortality data were available in 32 trials; 26 with at least one event contributed to the pooled analysis. SGLT2 inhibitors were associated with a significant reduction in ACM (RR 0.90 [0.83, 0.98], p = 0.016), with no heterogeneity (I^2^ = 0%; Figure 2). In a sensitivity analysis using hazard ratios from 4 trials, the result was consistent (HR 0.92 [0.80, 1.06], p = 0.154). The NNT to prevent one death was 78 (based on a control event rate of 12.8% over a weighted mean follow-up of approximately 86 weeks).

**Figure 2.**
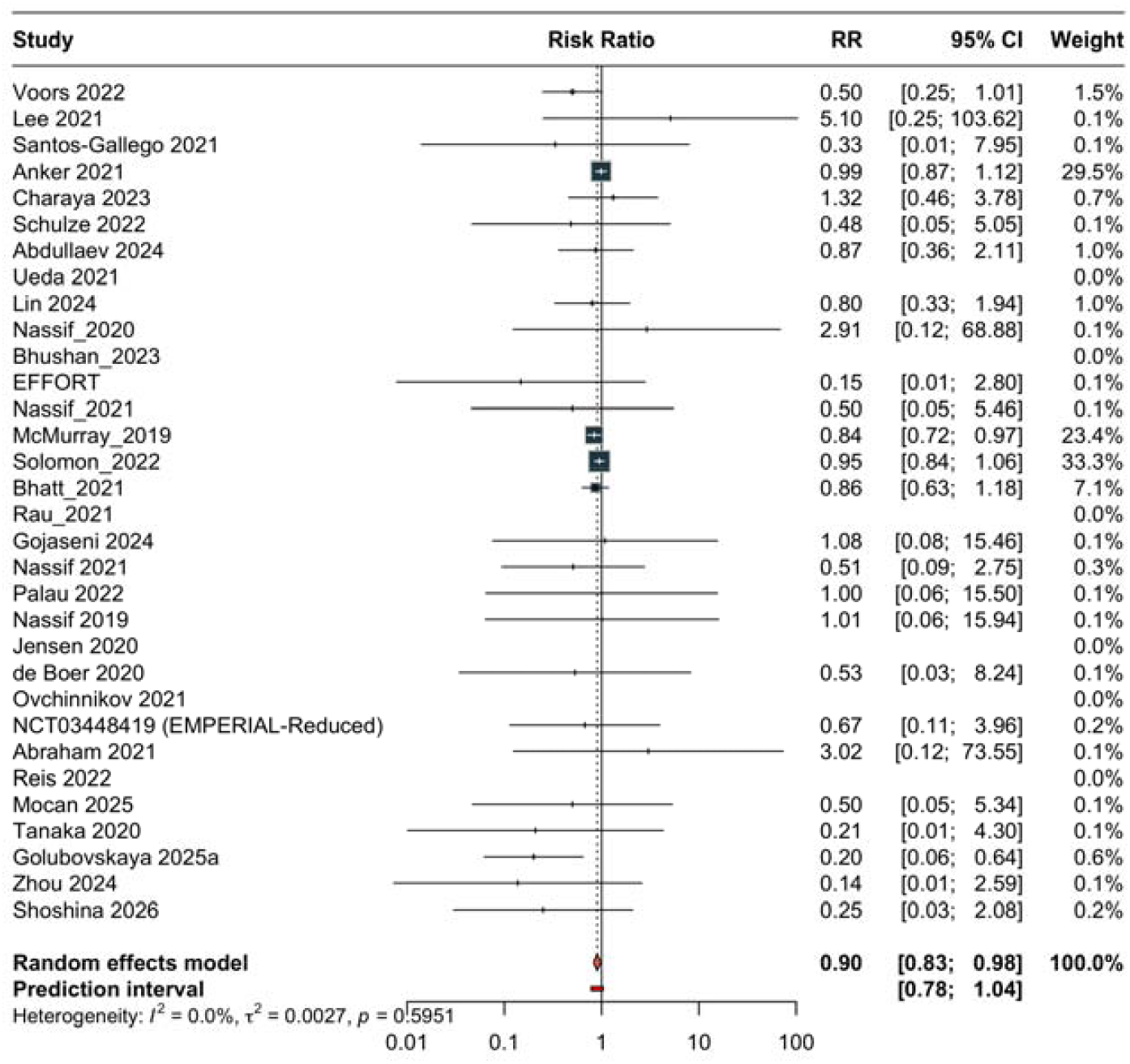
Forest plot for all-cause mortality.

The certainty of evidence was rated as low, downgraded for risk of bias and publication bias (Egger’s test p = 0.028). Trim-and-fill analysis imputed 6 missing studies and yielded an adjusted RR of 0.92.

#### Heart Failure Hospitalization

15 trials reported HFH events. SGLT2 inhibitors significantly reduced HFH (RR 0.74 [0.69, 0.79], p < 0.001), with no heterogeneity (I^2^ = 0%; Figure 3). The HR-based sensitivity analysis confirmed this finding (HR 0.72 [0.66, 0.78], p = 0.001; 4 trials). The NNT was 29 (control event rate 13.5% over a weighted mean follow-up of approximately 91 weeks).

**Figure 3.**
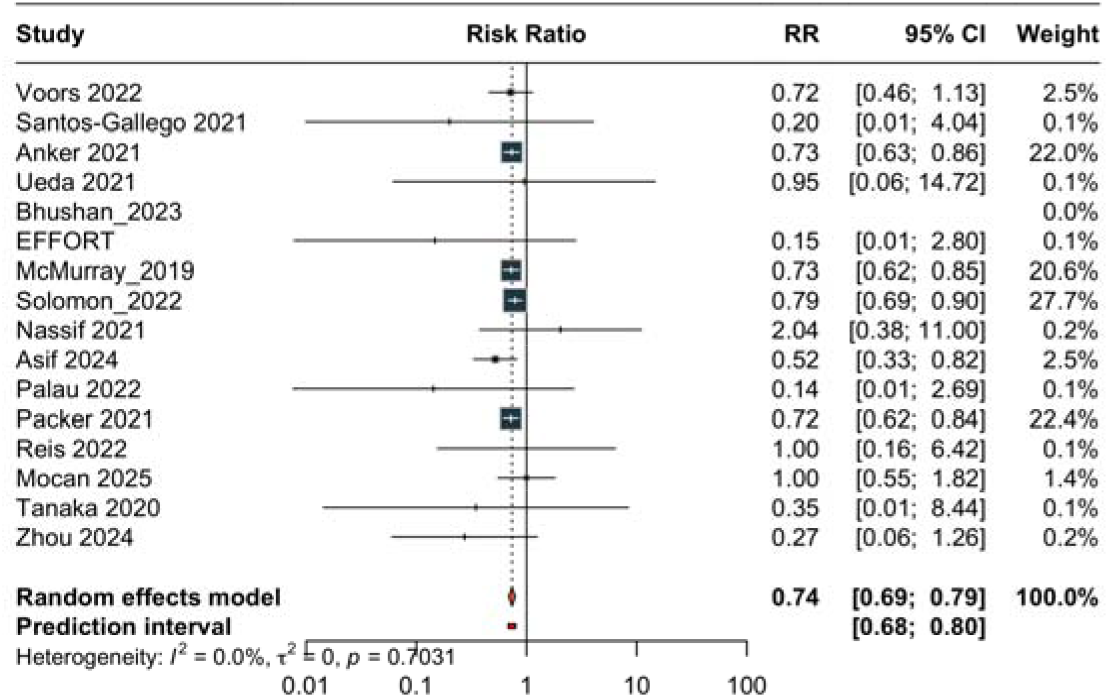
Forest plot for HF hospitalization.

**Figure 4.**
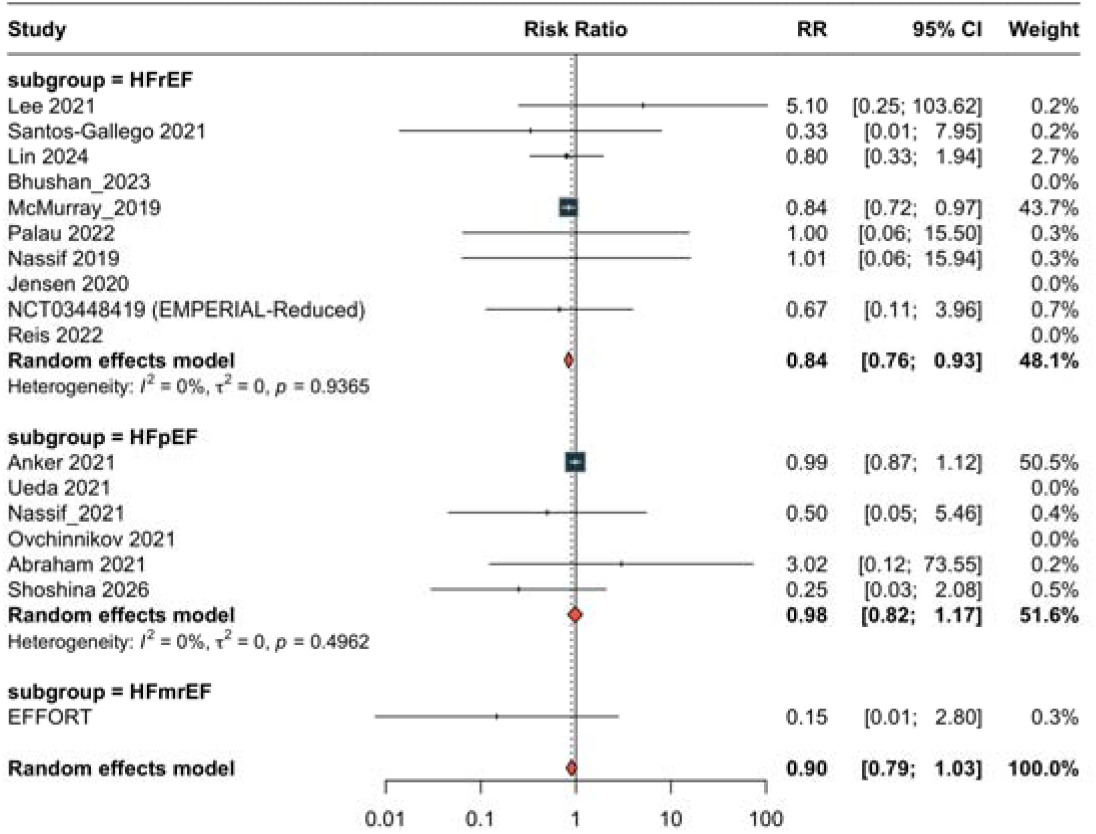
Subgroup analysis for all-cause mortality by HF type.

The certainty of evidence was rated as moderate, downgraded one level for risk of bias.

#### Secondary Outcomes

Cardiovascular death was significantly reduced (RR 0.86 [0.76, 0.98], p = 0.031; 7 trials I^2^ = 18%; moderate certainty; NNT 81). The composite endpoint was significantly reduced (RR 0.80 [0.75, 0.85], p < 0.001; 8 trials; I^2^ = 0%; high certainty; NNT 24).

Serious adverse events were lower (RR 0.94 [0.90, 0.99], p = 0.017; 20 trials; I^2^ = 36%; high certainty; NNT 42).

KCCQ total symptom score improved by 2.6 points (95% CI 1.2 to 4.0; p = 0.002; 9 trials; high certainty). KCCQ overall summary score showed a non-significant trend (MD 2.8 points; 3 trials; moderate certainty).

Six-minute walk distance showed no significant improvement (MD 3.9 meters; 8 trials; I^2^ = 68%; low certainty). NT-proBNP change was not significantly different between groups (MD -30.0 pg/mL, 95% CI -89.0 to 29.0; p = 0.269; 8 trials; I^2^ = 66%; very low certainty); two studies reporting geometric mean ratios rather than absolute differences were excluded from this analysis to avoid scale mixing. Body weight was reduced (MD -1.3 kg; p = 0.048; 9 trials; moderate certainty).

#### Safety Outcomes

Genital infections were significantly more common with SGLT2 inhibitors (RR 3.75 [1.72, 8.19], p = 0.007; 6 trials; I^2^ = 0%). No significant increase was observed for diabetic ketoacidosis (RR 1.47; 4 trials), acute kidney injury (RR 0.89; 13 trials), urinary tract infections (RR 1.01; 13 trials), or hypotension (RR 0.99; 8 trials).

#### Subgroup Analyses

For ACM, the test for subgroup differences was significant by HF type (interaction p = 0.039), with a greater benefit in HFrEF (RR 0.84, 95% CI 0.76-0.93; 7 trials) than HFpEF (RR 0.98, 95% CI 0.82-1.17; 4 trials). Significant interactions were also observed for follow-up duration (interaction p = 0.047), with a stronger effect in trials with less than 12 months of follow-up (RR 0.71) than those with 12 months or more, sample size (interaction p = 0.002), with stronger effects in smaller trials, and publication era (interaction p = 0.009), with more pronounced effects in recent publications. No significant interaction was detected for drug class (p = 0.247), blinding status (p = 0.142), or risk of bias (p = 0.251).

For HFH, the benefit was consistent across all subgroups. A significant interaction was observed for blinding (interaction p = 0.018), with a stronger effect in double-blind trials (RR 0.74) than open-label trials (RR 0.97). No significant interactions were found for HF type (p = 0.132), drug class (p = 0.424), follow-up duration (p = 0.669), sample size (p = 0.246), risk of bias (p = 0.960), or publication era (p = 0.937).

#### Sensitivity Analyses

The ACM result was robust across all 12 filter-based sensitivity analyses, with point estimates ranging from RR 0.68 to 0.98. The effect remained significant when excluding high risk of bias studies (RR 0.91, 95% CI 0.85-0.98), excluding open-label trials (RR 0.91, 95% CI 0.85-0.98), and when restricted to dapagliflozin only (RR 0.89, 95% CI 0.82-0.98). The effect was not significant in HFpEF-only (RR 0.98, p = 0.763) or low RoB-only (RR 0.92, p = 0.056) analyses, though the direction was consistent.

Alternative statistical models yielded consistent results: fixed-effect MH-RR 0.91 (p = 0.005), random-effects OR 0.88 (p = 0.015), fixed-effect OR 0.89 (p = 0.005), and Peto OR 0.89 (p = 0.005).

The HFH result was consistent across all sensitivity analyses and all alternative statistical models (all p < 0.001).

#### Publication Bias

Egger’s test was significant for ACM (p = 0.028), suggesting possible small-study effects. Trim-and-fill analysis imputed 6 studies and yielded an adjusted RR of 0.92, maintaining the direction of benefit. Egger’s test was not significant for HFH (p = 0.189) or SAE (p = 0.431). NT-proBNP (k = 8) did not meet the minimum threshold of 10 studies for Egger’s test. Cumulative meta-analysis showed that the ACM effect estimate stabilized after 2022 and the HFH estimate was stable throughout the accrual of evidence.

#### Certainty of Evidence

The GRADE assessment yielded high certainty for the composite of CVD and HFH, SAE, and KCCQ-TSS; moderate certainty for HFH, CVD, KCCQ-OSS, and body weight; low certainty for ACM, 6MWD, and eGFR; and very low certainty for NT-proBNP and SBP (Table 2).

#### Summary of Findings

**Table 2.** GRADE Summary of Findings.

| Outcome | k | Measure | Effect [95% CI] | p | I<sup>2</sup> | Certainty |
| --- | --- | --- | --- | --- | --- | --- |
| ACM (RR) | 26 | RR | 0.90 [0.83, 0.98] | 0.016 | 0% | Low |
| HFH (RR) | 15 | RR | 0.74 [0.69, 0.79] | < 0.001 | 0% | Moderate |
| CVD (RR) | 7 | RR | 0.86 [0.76, 0.98] | 0.031 | 18% | Moderate |
| Composite (RR) | 8 | RR | 0.80 [0.75, 0.85] | < 0.001 | 0% | High |
| SAE (RR) | 20 | RR | 0.94 [0.90, 0.99] | 0.017 | 36% | High |
| KCCQ-TSS | 9 | MD | 2.6 [1.2, 4.0] | 0.002 | 50% | High |
| KCCQ-OSS | 3 | MD | 2.8 [-1.9, 7.5] | 0.127 | 25% | Moderate |
| 6MWD | 8 | MD | 3.9 [-6.5, 14.4] | 0.406 | 68% | Low |
| NT-proBNP | 8 | MD | -30.0 [-89.0, 29.0] | 0.269 | 66% | Very low |
| eGFR | 3 | MD | -0.1 [-2.3, 2.1] | 0.886 | 0% | Low |
| Weight | 9 | MD | -1.3 [-2.5, -0.0] | 0.048 | 84% | Moderate |
| SBP | 8 | MD | -1.3 [-4.8, 2.2] | 0.398 | 62% | Very low |

## Discussion

### Summary of Evidence

This comprehensive meta-analysis of 59 RCTs enrolling 29,692 participants demonstrates that SGLT2 inhibitors reduce all-cause mortality by 10%, heart failure hospitalization by 26%, cardiovascular death by 14%, the composite of cardiovascular death and HFH by 20%, and serious adverse events by 6% in adults with heart failure. These benefits were accompanied by improved quality of life (KCCQ-TSS) and modest weight reduction, with genital infections as the main safety concern. The effects were robust across multiple sensitivity analyses and alternative statistical approaches.

### Comparison with Previous Meta-Analyses

Our findings are broadly consistent with prior meta-analyses by Zannad et al. (13), Vaduganathan et al. (14), and Salah et al. (15), as well as the large collaborative analysis by the Nuffield-SGLT2i consortium (29), which reported similar magnitudes of benefit for HFH and the composite endpoint. However, our analysis extends the evidence base considerably: by including 59 RCTs compared with 4 to 10 in prior syntheses, we were able to demonstrate a statistically significant reduction in ACM (RR 0.90, p = 0.016) that was not consistently observed in earlier analyses that were limited to the pivotal trials. The inclusion of smaller trials also enabled more informative subgroup and meta-regression analyses, and allowed the assessment of safety outcomes with greater statistical power (e.g., genital infections in 6 trials, AKI in 13 trials).

The ACM finding merits careful interpretation. Although the pooled estimate was statistically significant with zero heterogeneity, the GRADE certainty was rated as low due to risk of bias concerns (36% of trials with high RoB) and evidence of publication bias (Egger p = 0.028). The trim-and-fill adjusted estimate (RR 0.92) remained in the direction of benefit but the CI likely crosses 1.0 in the adjusted analysis, suggesting that small-study effects may have inflated the observed benefit. Notably, the ACM effect was largely driven by HFrEF trials, where the subgroup estimate was strongly significant (RR 0.84, p = 0.005), whereas no benefit was observed in HFpEF (RR 0.98, p = 0.763). This pattern is consistent with the individual trial data from DAPA-HF and EMPEROR-Reduced (mortality benefit) versus DELIVER and EMPEROR-Preserved (no mortality benefit) (5–8).

### HF Subtype Effects

The significant interaction between SGLT2 inhibitor treatment and HF subtype for mortality (p = 0.039) supports the hypothesis that the mechanisms underlying mortality reduction — which may include improved myocardial energetics, reduced cardiac fibrosis, and neurohormonal modulation (4,30) — are more relevant in the failing and remodeled ventricle characteristic of HFrEF. In contrast, the HFH benefit was consistent across HF subtypes (HFrEF RR 0.72, HFpEF RR 0.73), suggesting that the diuretic and hemodynamic effects of SGLT2 inhibitors, including natriuresis, reduced preload, and decreased interstitial congestion (4), benefit patients regardless of ejection fraction.

### Drug Class Effects

No significant heterogeneity was observed between individual SGLT2 inhibitors for either primary outcome. The majority of evidence came from empagliflozin (25 trials) and dapagliflozin (24 trials), with limited data for canagliflozin (3 trials) and sotagliflozin (1 trial). While the absence of a significant drug class interaction supports a class effect, the confidence intervals for the less-studied agents were wide, and firm conclusions about equivalence cannot be drawn.

### Safety Profile

The safety findings were reassuring overall. Serious adverse events were actually lower with SGLT2 inhibitors (RR 0.94, high certainty), confirming that the benefits of therapy are not offset by increased serious harms. The 3.75-fold increase in genital infections is consistent with the known pharmacological mechanism of glycosuria and is a well-characterized class effect. Importantly, no significant increase in diabetic ketoacidosis, acute kidney injury, urinary tract infections, or hypotension was observed. The point estimate for AKI was numerically lower with SGLT2 inhibitors (RR 0.89), consistent with the nephroprotective effects observed in dedicated renal trials (31,32).

### Strengths

This review has several strengths. First, the comprehensive search across four databases with no language or date restrictions, combined with an eight-pass full-text screening pipeline, maximized the completeness of study identification. Second, full-text PDF extraction for all included studies — rather than reliance on abstracts alone — substantially increased the yield of extractable outcome data (e.g., ACM was available in 26 trials versus approximately 11 if limited to abstract reporting). Third, the extensive sensitivity analysis program (12 filter-based, 4 alternative models, leave-one-out, and influence diagnostics) provided robust assessment of result stability. Fourth, the use of HKSJ confidence intervals provides more appropriate coverage than the standard DerSimonian-Laird method, particularly with heterogeneous or small numbers of studies (20,21). Fifth, the application of GRADE, trial sequential analysis, and cumulative meta-analysis adds dimensions of certainty assessment beyond traditional meta-analytic methods (25,26).

### Limitations

Several limitations should be considered. First, approximately 36% of included trials were rated as high risk of bias, primarily due to open-label designs and lack of prospective registration. Although sensitivity analyses excluding these trials did not substantially alter the primary results, the overall certainty for ACM was downgraded accordingly. Second, publication bias was detected for ACM, and while the trim-and-fill adjustment maintained the direction of benefit, small-study effects cannot be fully excluded as a source of bias. Third, the smaller trials that contributed many of the additional events compared with prior meta-analyses may differ systematically from the large multinational pivotal trials in terms of patient populations, care settings, and outcome ascertainment. The significant subgroup interaction by sample size (p = 0.002) supports this concern. Fourth, continuous outcomes were limited by sparse reporting of between-group differences with confidence intervals, requiring imputation in some cases; for NT-proBNP, two studies reporting geometric mean ratios were excluded from the analysis to avoid pooling with absolute mean differences, reducing k from 10 to 8.

Fifth, the median follow-up of 13 weeks is relatively short, and the longer-term effects on mortality may differ from the pooled short-term estimate. Sixth, we could not assess diabetes status as a subgroup variable because this information was inconsistently reported across trials.

### Implications

#### For Clinical Practice

These findings reinforce current guideline recommendations for SGLT2 inhibitors as foundational therapy in HF (3,12). The NNT of 29 for HFH and 24 for the composite endpoint — derived from a pooled control event rate dominated by larger trials with follow-up exceeding one year — underscores the clinical magnitude of benefit.

Clinicians should be aware of the increased risk of genital infections and counsel patients accordingly, while the absence of increased AKI, DKA, or hypotension should provide reassurance when initiating therapy.

#### For Research

Future trials should prioritize longer follow-up to establish durability of the mortality benefit, direct comparisons between SGLT2 inhibitors to assess potential within-class differences, and enrollment of underrepresented populations including those with acute decompensated HF and advanced HF. The role of SGLT2 inhibitors in HFmrEF — where only 3 trials were available — warrants dedicated investigation. Additionally, the emerging agents (henagliflozin, licogliflozin) and dual SGLT1/2 inhibitors (sotagliflozin) deserve further evaluation in HF-specific trials.

## Conclusion

SGLT2 inhibitors significantly reduce all-cause mortality, heart failure hospitalization, cardiovascular death, the composite of cardiovascular death and HFH, and serious adverse events in adults with heart failure, with consistent benefits across drugs and HF subtypes. The mortality benefit is most pronounced in HFrEF. The safety profile is favorable, with genital infections as the only significantly increased adverse event.

These findings support the use of SGLT2 inhibitors as a foundational pillar of heart failure therapy across the ejection fraction spectrum.

## Data Availability

All data and code are available at Zenodo: 10.5281/zenodo.19100266

https://zenodo.org/records/19100266

