## supplement for "Efficacy and Safety of Sodium-Glucose Co-Transporter 2 Inhibitors in Heart Failure: A Systematic Review and Meta-Analysis of 59 Randomized Controlled Trials"

2026-03-18

### eAppendix 1: Search Strategies

#### PubMed

```
# PubMed Search Strategy
# SR: SGLT2 Inhibitors in Heart Failure
# PROSPERO: CRD420251167908
# Date designed: 2026-03-02
# Execute at: https://pubmed.ncbi.nlm.nih.gov/

# === BLOCK 1: Intervention (SGLT2 Inhibitors) ===

("Sodium-Glucose Transporter 2 Inhibitors"[MeSH Terms] OR "Sodium-Glucose Tra
nsporter 2 Inhibitors"[Pharmacological Action] OR "SGLT2 inhibitor*"[tiab] OR
 "SGLT-2 inhibitor*"[tiab] OR "sodium-glucose cotransporter 2 inhibitor*"[tia
b] OR "sodium glucose cotransporter 2 inhibitor*"[tiab] OR "gliflozin*"[tiab]
 OR "empagliflozin"[tiab] OR "dapagliflozin"[tiab] OR "canagliflozin"[tiab] O
R "sotagliflozin"[tiab] OR "ertugliflozin"[tiab] OR "ipragliflozin"[tiab] OR
"luseogliflozin"[tiab] OR "tofogliflozin"[tiab] OR "remogliflozin"[tiab] OR "
bexagliflozin"[tiab] OR "henagliflozin"[tiab] OR "licogliflozin"[tiab])

# === BLOCK 2: Population (Heart Failure — all phenotypes) ===

("Heart Failure"[MeSH Terms] OR "heart failure"[tiab] OR "cardiac failure"[ti
ab] OR "congestive heart failure"[tiab] OR "HFrEF"[tiab] OR "HFpEF"[tiab] OR
"HFmrEF"[tiab] OR "preserved ejection fraction"[tiab] OR "reduced ejection fr
action"[tiab] OR "mildly reduced ejection fraction"[tiab] OR "systolic dysfun
ction"[tiab] OR "diastolic dysfunction"[tiab] OR "ventricular dysfunction"[ti
ab])

# === BLOCK 3: Study Design (RCT filter) ===

("randomized controlled trial"[pt] OR "controlled clinical trial"[pt] OR "ran
domized"[tiab] OR "randomised"[tiab] OR "placebo"[tiab] OR "trial"[tiab])

# === COMBINED SEARCH ===
# Copy-paste the three blocks joined by AND:

("Sodium-Glucose Transporter 2 Inhibitors"[MeSH Terms] OR "Sodium-Glucose Tra
nsporter 2 Inhibitors"[Pharmacological Action] OR "SGLT2 inhibitor*"[tiab] OR
 "SGLT-2 inhibitor*"[tiab] OR "sodium-glucose cotransporter 2 inhibitor*"[tia
b] OR "sodium glucose cotransporter 2 inhibitor*"[tiab] OR "gliflozin*"[tiab]
 OR "empagliflozin"[tiab] OR "dapagliflozin"[tiab] OR "canagliflozin"[tiab] O
R "sotagliflozin"[tiab] OR "ertugliflozin"[tiab] OR "ipragliflozin"[tiab] OR
"luseogliflozin"[tiab] OR "tofogliflozin"[tiab] OR "remogliflozin"[tiab] OR "
bexagliflozin"[tiab] OR "henagliflozin"[tiab] OR "licogliflozin"[tiab]) AND
("Heart Failure"[MeSH Terms] OR "heart failure"[tiab] OR "cardiac failure"[ti
ab] OR "congestive heart failure"[tiab] OR "HFrEF"[tiab] OR "HFpEF"[tiab] OR
```

```
"HFmrEF"[tiab] OR "preserved ejection fraction"[tiab] OR "reduced ejection fr
action"[tiab] OR "mildly reduced ejection fraction"[tiab] OR "systolic dysfun
ction"[tiab] OR "diastolic dysfunction"[tiab] OR "ventricular dysfunction"[ti
ab]) AND ("randomized controlled trial"[pt] OR "controlled clinical trial"[pt]
 OR "randomized"[tiab] OR "randomised"[tiab] OR "placebo"[tiab] OR "trial"[ti
ab])

# Notes:
# - No date or language restrictions
# - MeSH "Sodium-Glucose Transporter 2 Inhibitors" introduced 2016
# - [Pharmacological Action] captures articles indexed by drug mechanism
# - "gliflozin*" wildcard catches all -gliflozin suffix drugs
# - All 12 SGLT2i from PROSPERO protocol listed individually
# - Export as CSV format with all fields
```

#### Cochrane CENTRAL

```
# Cochrane CENTRAL Search Strategy
# SR: SGLT2 Inhibitors in Heart Failure
# PROSPERO: CRD420251167908
# Date designed: 2026-03-02
# Execute at: https://www.cochranelibrary.com/advanced-search/search-manager

# === STEP-BY-STEP (Search Manager) ===

#1 [mh "Sodium-Glucose Transporter 2 Inhibitors"]
#2 ((SGLT2 NEXT inhibitor*) OR (SGLT-2 NEXT inhibitor*) OR (sodium-glucose NE
XT cotransporter NEXT 2 NEXT inhibitor*) OR (sodium NEXT glucose NEXT cotrans
porter NEXT 2 NEXT inhibitor*) OR gliflozin* OR empagliflozin OR dapagliflozi
n OR canagliflozin OR sotagliflozin OR ertugliflozin OR ipragliflozin OR luse
ogliflozin OR tofogliflozin OR remogliflozin OR bexagliflozin OR henagliflozi
n OR licogliflozin):ti,ab,kw
#3 #1 OR #2
#4 [mh "Heart Failure"]
#5 ("heart failure" OR "cardiac failure" OR "congestive heart failure" OR HFr
EF OR HFpEF OR HFmrEF OR "preserved ejection fraction" OR "reduced ejection f
raction" OR "mildly reduced ejection fraction" OR "systolic dysfunction" OR "
diastolic dysfunction" OR "ventricular dysfunction"):ti,ab,kw
#6 #4 OR #5
#7 #3 AND #6

# Filter: Cochrane Central Register of Controlled Trials (CENTRAL) only
# Export: CSV format with all fields
# No date or language restrictions

# === FLAT SEARCH (alternative if Search Manager unavailable) ===
# Paste into simple search box, filter to Trials:
#
# ("SGLT2 inhibitor" OR "SGLT-2 inhibitor" OR "sodium-glucose cotransporter 2
 inhibitor" OR gliflozin OR empagliflozin OR dapagliflozin OR canagliflozin O
```

```
R sotagliflozin OR ertugliflozin OR ipragliflozin OR luseogliflozin OR tofogl
iflozin OR remogliflozin OR bexagliflozin OR henagliflozin OR licogliflozin)
AND ("heart failure" OR "cardiac failure" OR HFrEF OR HFpEF OR HFmrEF OR "ven
tricular dysfunction")

# Notes:
# - [mh] syntax = MeSH descriptor with explode (default)
# - No RCT filter needed — CENTRAL contains only trials
# - Old syntax "MeSH descriptor: [Term] explode all trees" deprecated
# - If [mh] fails, use MeSH browser point-and-click, then combine
```

#### ClinicalTrials.gov

```
# ClinicalTrials.gov Search Strategy
# SR: SGLT2 Inhibitors in Heart Failure
# PROSPERO: CRD420251167908
# Date designed: 2026-03-02
# Execute at: https://clinicaltrials.gov/search

# === WEB INTERFACE ===

Condition or Disease: Heart Failure
Intervention / Treatment: SGLT2 inhibitor OR empagliflozin OR dapagliflozin O
R canagliflozin OR sotagliflozin OR ertugliflozin OR ipragliflozin OR luseogl
iflozin OR tofogliflozin OR remogliflozin OR bexagliflozin OR henagliflozin O
R licogliflozin OR gliflozin
Study Type: Interventional (Clinical Trial)

# === API v2 QUERY ===
# https://clinicaltrials.gov/api/v2/studies?query.cond=Heart+Failure&query.in
tr=SGLT2+inhibitor+OR+empagliflozin+OR+dapagliflozin+OR+canagliflozin+OR+sota
gliflozin+OR+ertugliflozin+OR+ipragliflozin+OR+luseogliflozin+OR+tofogliflozi
n+OR+remogliflozin+OR+bexagliflozin+OR+henagliflozin+OR+licogliflozin+OR+glif
lozin&filter.studyType=INTERVENTIONAL&pageSize=100

# Notes:
# - No date or status restrictions
# - Export as CSV with all fields
# - Check for multi-page results (pageSize=100)
# - ClinicalTrials.gov uses free-text matching, no MeSH
# - Captures registered trials including those without publications
```

#### WHO ICTRP

```
# WHO ICTRP Search Strategy
# SR: SGLT2 Inhibitors in Heart Failure
# PROSPERO: CRD420251167908
# Date designed: 2026-03-02
# Execute at: https://trialsearch.who.int/
```

```
# WHO ICTRP has limited search syntax. Run multiple searches and combine.

# === Search 1 (class term) ===
Title: SGLT2 AND heart failure
Condition: heart failure
Recruitment Status: ALL

# === Search 2 (class term variant) ===
Title: gliflozin AND heart failure
Condition: heart failure
Recruitment Status: ALL

# === Search 3 (empagliflozin) ===
Title: empagliflozin
Condition: heart failure
Recruitment Status: ALL

# === Search 4 (dapagliflozin) ===
Title: dapagliflozin
Condition: heart failure
Recruitment Status: ALL

# === Search 5 (canagliflozin) ===
Title: canagliflozin
Condition: heart failure
Recruitment Status: ALL

# === Search 6 (sotagliflozin) ===
Title: sotagliflozin
Condition: heart failure
Recruitment Status: ALL

# === Search 7 (other SGLT2i — less common) ===
Title: ertugliflozin OR ipragliflozin OR luseogliflozin OR tofogliflozin
Condition: heart failure
Recruitment Status: ALL

# Notes:
# - WHO ICTRP aggregates 17+ registries (ISRCTN, ANZCTR, ChiCTR, JPRN, etc.)
# - Export each search as CSV, combine, then deduplicate
# - Captures trials not registered on ClinicalTrials.gov
# - Particularly important for Japanese (JPRN) and Chinese (ChiCTR) trials
#   where luseogliflozin, tofogliflozin, and henagliflozin are studied
# - No date or language restrictions
```

### eTable 1: Characteristics of Included Studies (n = 59)

| Study | Trial | Year | Country | Blinding | HF Type | N | Drug | Dose (mg) | Comparator | FU (wk) | Age (y) | Female (%) | LVEF (%) | DM (%) | RoB |
|---|---|---|---|---|---|---|---|---|---|---|---|---|---|---|---|
| Abdullaev 2024 | | 2,024 | Russia | Open-label | Mixed (NYHA III-IV) | 119 | Dapagliflozin/Empagliflozin | 10 | Standard diuretic therapy (loop diuretics only) | 4.3 | 71.90 | 45.4 | 37.40 | 100.00 | High |
| Abedi_2024 | Abedi 2024 | 2,024 | Iran | Double-blind | HFrEF and HFmrEF | 80 | Empagliflozin | 10 | Placebo | 24.0 | 60.27 | 22.2 | 24.73 | 29.20 | Some concerns |
| Abraham 2021 | EMPERIAL | 2,021 | USA; Australia; Canada; Germany; Greece; Italy; Norway; Poland; Portugal; Spain; Sweden | Double-blind | HFpEF | 315 | empagliflozin | 10 | placebo | 12.0 | 73.50 | 43.2 | 53.10 | 51.10 | Low |
| Akasaka 2022 | EXCEED | 2,022 | Japan | Open-label | HFpEF | 73 | Ipragliflozin | 50 | Conventional treatment | 24.0 | 71.10 | 39.7 | 60.70 | 100.00 | High |
| Anker 2021 | EMPEROR-Preserved | 2,021 | Multinational (23 countries) | Double-blind | HFpEF | 5,988 | Empagliflozin | 10 | Placebo | 113.7 | 71.90 | 44.6 | 54.30 | 49.10 | Low |
| Asif 2024 | | 2,024 | Pakistan | Double-blind | HFmrEF | 480 | dapagliflozin | 10 | placebo | 4.0 | 52.85 | 44.0 | | 69.38 | High |
| Bhatt_2021 | SOLOIST-WHF | 2,021 | Multinational (32 countries) | Double-blind | Mixed (HFrEF and HFpEF; 79.1% LVEF <50%) | 1,222 | Sotagliflozin | 200 (up to 400) | Placebo | 39.1 | 69.50 | 33.7 | | 100.00 | Some concerns |

| Study | Trial | Year | Country | Blinding | HF Type | N | Drug | Dose (mg) | Comparator | FU (wk) | Age (y) | Female (%) | LVEF (%) | DM (%) | RoB |
|---|---|---|---|---|---|---|---|---|---|---|---|---|---|---|---|
| Bhushan_2023 | Remo Safe-AHF | 2,023 | India | Open-label | HFrEF | 35 | Remogliflozin | | Conventional therapy | 12.0 | | | 29.87 | 100.00 | High |
| Borlaug 2023 | CAMEO-DAPA | 2,023 | United States | Double-blind | HFpEF | 38 | dapagliflozin | 10 | placebo | 25.0 | 67.00 | 66.0 | 62.00 | 18.00 | Low |
| Charaya 2023 | | 2,023 | Russia | Open-label | HFrEF and HFmrEF and HFpEF (ADHF) | 200 | Dapagliflozin | 10 | Standard therapy for ADHF (IV loop diuretics) | 4.0 | 74.00 | 49.0 | 47.00 | 31.00 | High |
| Damman 2020 | EMPA-RESPONSE-AHF | 2,020 | Netherlands | Double-blind | acute HF | 80 | empagliflozin | 10 | placebo | 8.6 | 76.00 | 33.0 | 36.00 | 33.00 | Some concerns |
| EFFORT | 10.1161/CIRCULATIONAHA.124.069144 | 2,020 | South Korea | Double-blind | HFmrEF | 128 | ertugliflozin | | placebo | 52.0 | 66.40 | 39.0 | 42.00 | 12.50 | Low |
| Ejiri 2020 | MUSCAT-HF | 2,020 | Japan | Open-label | HFpEF | 169 | luseogliflozin | 2.5 | voglibose | 24.0 | 73.10 | 37.6 | 57.50 | 100.00 | High |
| Emara 2023 | DAPA-RESPONSE-AHF | 2,023 | | Double-blind | Acute HF | 87 | dapagliflozin | 10 | Placebo | 4.0 | | | | | Some concerns |
| Fatima Gilani 2023 | | 2,023 | Pakistan | Open-label | AHF (LVEF <40%) | 160 | dapagliflozin | 10 | standard medical therapy (placebo-free) | 1.0 | 65.13 | 18.8 | | 51.25 | High |
| Fatima Gilani 2024 | | 2,024 | Pakistan | Open-label | AHF (EF <40%) | 150 | dapagliflozin | 10 | conventional therapy alone | 12.0 | 63.76 | 17.3 | | 48.00 | High |
| Fu_2023 | | 2,02 | China | Double- | HFrEF | 60 | dapagliflozin | 10 | placebo | 52. | 70.5 | 28.3 | 30.9 | 100. | Some con |

| Study | Trial | Year | Country | Blinding | HF Type | N | Drug | Dose (mg) | Comparator | FU (wk) | Age (y) | Female (%) | LVEF (%) | DM (%) | RoB |
|---|---|---|---|---|---|---|---|---|---|---|---|---|---|---|---|
| | | 3 | | blind | | | | | | 0 | 5 | | 5 | 00 | cerns |
| Gojaseni 2024 | CO-0482 | 2,024 | Thailand | Open-label | acute decompensated HF | 32 | dapagliflozin | 10 | standard of care | 4.0 | 67.10 | 51.5 | 42.10 | | High |
| Golubovskaya 2025a | Golubovskaya 2025a | 2,025 | Russia | Open-label | HFrEF/HFmrEF/HFpEF | 92 | empagliflozin | 10 | standard care | 26.0 | | 31.5 | | 43.50 | High |
| Griffin 2020 | Griffin 2020 | 2,020 | USA | Double-blind | Mixed (HFrEF 45% + HFpEF) | 20 | Empagliflozin | 10 | Placebo | 6.0 | 60.00 | 25.0 | 42.90 | 100.00 | Some concerns |
| Hundertmark 2023 | EMPA-VISION | 2,023 | UK | Double-blind | HFrEF+HFpEF | 72 | empagliflozin | 10 | Placebo | 12.0 | 68.33 | 41.7 | | 12.50 | Some concerns |
| Ilyas 2021 | | 2,021 | Australia | Double-blind | HFrEF | 19 | dapagliflozin | 10 | placebo | 8.0 | 73.00 | 26.0 | 35.00 | 100.00 | Some concerns |
| Jensen 2020 | Empire HF | 2,020 | Denmark | Double-blind | HFrEF | 190 | empagliflozin | 10 | placebo | 12.0 | 64.00 | 15.0 | 29.00 | 18.00 | Some concerns |
| Kawanami 2025 | ROAD-ADHF | 2,025 | Japan | Open-label | HFrEF/HFmrEF | 117 | dapagliflozin | 10 | conventional therapy (loop diuretics alone) | 1.0 | 73.00 | 35.0 | 33.00 | | High |
| Kolwelter 2021a | NCT03128528 | 2,021 | Germany | Double-blind | Mixed (HFrEF 61% + HFmrEF) | 75 | Empagliflozin | 10 | Placebo | 13.0 | 66.00 | 15.0 | 39.00 | 23.00 | Some concerns |
| Lee 2021 | SUGAR-DM-HF | 2,021 | United Kingdom (Scotland) | Double-blind | HFrEF | 105 | empagliflozin | 10 | placebo | 36.0 | 68.70 | 26.7 | 32.50 | 78.10 | Low |
| Lin 2024 | | 2,024 | China | Double-blind | HFrEF | 200 | Dapagliflozin | 10 | Placebo | 104.0 | 62.70 | 33.0 | 32.85 | 41.00 | Some concerns |

| Study | Trial | Year | Country | Blinding | HF Type | N | Drug | Dose (mg) | Comparator | FU (wk) | Age (y) | Female (%) | LVEF (%) | DM (%) | RoB |
|---|---|---|---|---|---|---|---|---|---|---|---|---|---|---|---|
| Marton 2024 | DAPA-Shuttle1 | 2,024 | Singapore | Double-blind | HFrEF | 40 | dapagliflozin | 10 | placebo | 4.0 | 59.00 | 10.3 | 31.00 | 48.30 | Some concerns |
| McMurray 2024 | DETERMINE | 2,024 | International (multinational) | Double-blind | HFrEF and HFpEF (two separate cohorts) | 817 | dapagliflozin | 10 | placebo | 16.0 | | | | | Some concerns |
| McMurray_2019 | DAPA-HF | 2,019 | Multinational (20 countries) | Double-blind | HFrEF | 4,744 | Dapagliflozin | 10 | Placebo | 79.0 | 66.35 | 23.8 | 31.05 | 41.80 | Low |
| Mocan 2025 | | 2,025 | Romania | Open-label | AHF (any EF) | 100 | dapagliflozin | 10 | structured IV furosemide alone | 4.0 | 63.63 | 18.4 | 26.03 | 20.40 | High |
| Mordi 2020 | RECEDE-CHF | 2,020 | United Kingdom | Double-blind | HFrEF | 23 | empagliflozin | 25 | placebo | 14.0 | 69.80 | 26.1 | | 100.00 | Low |
| NCT03448419 (EMPERIAL-Reduced) | EMPERIAL-Reduced | 2,018 | USA; Australia; Canada; Germany; Greece; Italy; Norway; Poland; Portugal; Spain; Sweden | Double-blind | HFrEF | 312 | empagliflozin | 10 | placebo | 12.0 | 69.00 | 25.6 | 30.00 | 59.90 | Low |
| Nassif 2019 | DEFINE-HF | 2,019 | United States | Double-blind | HFrEF | 263 | dapagliflozin | 10 | placebo | 13.0 | 61.30 | 27.0 | 26.00 | 62.00 | Some concerns |
| Nassif 2021 | CHIEF-HF | 2,022 | USA | Double-blind | HFrEF+HFpEF | 476 | canagliflozin | 100 | placebo | 12.0 | 63.40 | 44.9 | | 27.90 | Some concerns |
| Nassif_2020 | EMBRACE-HF | 2,0 | United States | Doubl | HFrEF+HFpEF | 65 | empagliflozin | 10 | placebo | 13. | 66. | 37.0 | 44. | 52.0 | Low |

| Study | Trial | Year | Country | Blinding | HF Type | N | Drug | Dose (mg) | Comparator | FU (wk) | Age (y) | Female (%) | LVEF (%) | DM (%) | RoB |
|---|---|---|---|---|---|---|---|---|---|---|---|---|---|---|---|
| | | 20 | | e-blind | | | | | | 0 | 00 | | 00 | 0 | |
| Nassif_2021 | PRESERVED-HF | 2,021 | United States | Double-blind | HFpEF | 324 | dapagliflozin | 10 | placebo | 13.0 | 70.00 | 57.0 | 60.00 | 56.00 | Low |
| Omar_2022 | Empire HF Biomarker | 2,022 | Denmark | Double-blind | HFrEF | 190 | empagliflozin | 10 | placebo | 12.0 | 64.00 | 15.0 | 29.00 | 12.00 | Some concerns |
| Ovchinnikov 2021 | Ovchinnikov 2021 | 2,021 | Russia | Open-label | HFpEF | 60 | empagliflozin | 10 | Standard care | 24.0 | 66.50 | 61.7 | 60.00 | 100.00 | High |
| Ovchinnikov_2025 | 10.1186/s12933-025-02756-y | 2,025 | Russia | Open-label | HFpEF | 70 | empagliflozin | 10 | standard care | 24.0 | 67.10 | 63.0 | 61.00 | 100.00 | High |
| Packer 2021 | EMPEROR-Reduced | 2,021 | Multinational (20 countries) | Double-blind | HFrEF | 3,730 | Empagliflozin | 10 | Placebo | 69.6 | | | | | Low |
| Palau 2022 | DAPA-VO2 | 2,022 | Spain | Double-blind | HFrEF | 90 | dapagliflozin | 10 | placebo | 12.0 | 68.60 | 23.3 | 33.80 | 32.20 | Low |
| Pastore 2024 | DAPA ECHO | 2,024 | Italy | Open-label | HFrEF and HFmrEF | 88 | Dapagliflozin | 10 | Optimal medical therapy (OMT) without SGLT2i | 26.0 | 68.00 | 17.0 | 37.00 | 0.00 | High |
| Rau_2021 | EudraCT 2016-000172-19 | 2,021 | Germany | Double-blind | T2D with ASCVD (43% chronic HF) | 44 | empagliflozin | 10 | placebo | 12.0 | 62.00 | 19.0 | | 100.00 | Some concerns |
| Reis 2022 | | 2,022 | Portugal | Open-label | HFrEF | 40 | Dapagliflozin | 10 | Optimal medical therapy | 26.0 | 60.90 | 17.5 | 34.10 | 0.00 | High |
| Santos- | EMPA-TROPISM | 2, | United | Do | HFrEF | 8 | empagliflozin | 10 | place | 2 | | | 3 | 0. | Lo |

| Study | Trial | Year | Country | Blinding | HF Type | N | Drug | Dose (mg) | Comparator | FU (wk) | Age (y) | Female (%) | LVEF (%) | DM (%) | RoB |
|---|---|---|---|---|---|---|---|---|---|---|---|---|---|---|---|
| Gallego 2021 | | 021 | States | uble-blind | | 4 | | | bo | 4.0 | | | 6.00 | 00 | w |
| Schulze 2022 | EMPAG-HF | 2,022 | Germany | Double-blind | Mixed (HFrEF + HFpEF) | 60 | Empagliflozin | 25 | Placebo | 4.3 | 74.70 | 38.0 | 44.50 | 39.00 | Low |
| Shirakabe_2020 | Shirakabe 2020 | 2,020 | Japan | Open-label | compensated HF (mixed EF) | 60 | empagliflozin | 10-25 | no treatment (standard care) | 26.0 | 74.00 | 17.2 | 55.00 | 100.00 | High |
| Shoshina 2026 | | 2,026 | Russia | Open-label | HFpEF | 50 | dapagliflozin | 10 | Standard care | 26.0 | 63.75 | 42.0 | | 6.00 | High |
| Solomon_2022 | DELIVER | 2,022 | 20 countries (multinational) | Double-blind | HFmrEF/HFpEF (LVEF >40 %) | 6,263 | dapagliflozin | 10 | placebo | 120.0 | 71.65 | 43.9 | 54.15 | 44.80 | Low |
| Tamaki 2021 | Tamaki 2021 | 2,021 | Japan | Open-label | mixed (HFrEF 49%/HFmrEF 14%/HFpEF 37%) | 62 | empagliflozin | 10 | conventional glucose-lowering therapy | 1.0 | | 39.0 | | 100.00 | Some concerns |
| Tanaka 2020 | CANDLE | 2,020 | Japan | Open-label | mixed (71% HFpEF; 29% HFrEF) | 245 | canagliflozin | 100 | glimepiride | 24.0 | 68.60 | 25.0 | 57.60 | 100.00 | Some concerns |
| Thiele 2022 | | 2,022 | Germany | Double-blind | mixed | 19 | empagliflozin | 10 | placebo | 4.3 | 72.10 | 52.6 | 36.00 | 26.30 | Some concerns |
| Ueda 2021 | CANONICAL | 2,021 | Japan | Open-label | HFpEF | 82 | Canagliflozin | 100 | Standard diabetic therapy | 24.0 | 75.70 | 32.9 | 61.50 | 100.00 | High |
| Voors 2022 | EMPULSE | 2,022 | Multinational (15 countries) | Double-blind | Both (HFrEF and HFpEF; acute de novo and decompens | 530 | Empagliflozin | 10 | Placebo | 13.0 | 70.30 | 33.8 | | 45.30 | Low |

| Study | Trial | Year | Country | Blinding | HF Type | N | Drug | Dose (mg) | Comparator | FU (wk) | Age (y) | Female (%) | LVEF (%) | DM (%) | RoB |
|---|---|---|---|---|---|---|---|---|---|---|---|---|---|---|---|
| | | | | | ated chronic) | | | | | | | | | | |
| Wu 2022 | 10.3760/cma.j.cn112148-20220120-00059 | 2,021 | China | Open-label | HFmrEF | 112 | empagliflozin | 10 | conventional therapy | 26.0 | 69.00 | 25.0 | | | High |
| Xie 2024 | DAHOS | 2,024 | China | Open-label | HFrEF | 120 | dapagliflozin | 10 | optimized HF therapy | 12.0 | 62.10 | 27.1 | 33.00 | 16.82 | High |
| Zhou 2024 | | 2,024 | China | Single-blind | HFrEF/HFmrEF | 98 | dapagliflozin | 10 | standard care | 52.0 | 66.63 | 40.8 | 48.67 | 56.12 | Some concerns |
| de Boer 2020 | de Boer 2020 (licogliflozin) | 2,020 | 21 countries | Double-blind | HFrEF + HFpEF (NYHA II-IV; mixed LVEF) | 125 | licogliflozin | 2.5/10/50 | placebo | 12.0 | | 28.2 | | 100.00 | Some concerns |

### eTable 2: Risk of Bias 2.0 Assessments (n = 59)

| Study | D1: Randomization | D2: Deviations | D3: Missing | D4: Measurement | D5: Reporting | Overall |
|---|---|---|---|---|---|---|
| Abdullaev 2024 | Some concerns | Some concerns | Some concerns | Low | Some concerns | Some concerns |
| Abedi_2024 | Low | Low | Some concerns | Low | Low | Low |
| Abraham 2021 | Low | Low | Some concerns | Low | Low | Some concerns |
| Akasaka 2022 | Some concerns | High | Some concerns | Some concerns | Low | High |
| Ali 2024 | Some concerns | Some concerns | Some concerns | Some concerns | Some concerns | High |
| Anker 2021 | Low | Low | Low | Low | Low | Low |
| Asif 2024 | Some concerns | Low | Some concerns | Low | Some concerns | High |
| Bhatt_2021 | Low | Low | Low | Low | Low | Low |
| Bhushan_2023 | Some concerns | Some concerns | Some concerns | Low | Some concerns | High |
| Boorsma 2021 | Low | Low | Some concerns | Low | Low | Some concerns |
| Borlaug 2023 | Low | Low | Low | Low | Low | Low |
| Bosch 2023 | Low | Low | Some concerns | Low | Some concerns | Some concerns |
| Charaya 2022 | Some concerns | High | Some concerns | Low | Low | High |
| Charaya 2023 | Some concerns | Some concerns | Some concerns | Low | Some concerns | Some concerns |
| Charaya 2023 | Some concerns | Some concerns | Some concerns | Low | Low | Some concerns |
| Damman 2020 | Low | Low | Some concerns | Low | Low | Some concerns |
| EASTER-HF | Some concerns | High | Some concerns | Some concerns | Some concerns | High |
| EFFORT | Low | Low | Low | Low | Low | Low |
| EUCTR2016-002280-34-DE 2016 | Some concerns | Some concerns | Some concerns | Some concerns | Some concerns | High |
| EUCTR2021-005446-15-NL | Some concerns | Some concerns | Some concerns | Some concerns | Some concerns | High |
| Ejiri 2019 | Low | Some | Some | Low | Some | Some |

| Study | D1: Randomization | D2: Deviations | D3: Missing | D4: Measurement | D5: Reporting | Overall |
|---|---|---|---|---|---|---|
| | | concerns | concerns | | concerns | concerns |
| Ejiri 2020 | Low | Some concerns | Low | Low | Some concerns | Some concerns |
| Emara 2023 | Low | Low | Some concerns | Low | Low | Some concerns |
| Fatima Gilani 2023 | Some concerns | Some concerns | Some concerns | Some concerns | Some concerns | High |
| Fatima Gilani 2024 | Some concerns | High | Some concerns | Some concerns | Low | High |
| Fu_2023 | Some concerns | Low | Some concerns | Low | Some concerns | Some concerns |
| Gojaseni 2024 | Some concerns | Some concerns | Some concerns | Low | Some concerns | High |
| Golubovskaya 2023 | Some concerns | Some concerns | Some concerns | Some concerns | Some concerns | Some concerns |
| Golubovskaya 2025a | Some concerns | Some concerns | Some concerns | Low | Some concerns | Some concerns |
| Golubovskaya 2025b | Some concerns | Some concerns | Some concerns | Some concerns | Some concerns | Some concerns |
| Griffin 2020 | Some concerns | Low | Some concerns | Low | Some concerns | Some concerns |
| Hundertmark 2023 | Low | Low | Some concerns | Low | Low | Some concerns |
| Ilyas 2021 | Low | Low | Some concerns | Low | Some concerns | Some concerns |
| Jensen 2020 | Low | Low | Some concerns | Low | Low | Some concerns |
| Jones 2020 | Low | Low | Some concerns | Low | Some concerns | Some concerns |
| Kawanami 2024 | Some concerns | Some concerns | Some concerns | Low | Low | Some concerns |
| Kawanami 2025 | Some concerns | Some concerns | Some concerns | Low | Low | Some concerns |
| Kolwelter 2021a | Some concerns | Low | Some concerns | Low | Some concerns | Some concerns |
| Kolwelter 2021b | Low | Low | Some concerns | Low | Some concerns | Some concerns |
| Kolwelter 2021c | Low | Low | Some concerns | Low | Some concerns | Some concerns |
| Kolwelter 2021d | Low | Low | Some concerns | Low | Some concerns | Some concerns |

| Study | D1: Randomization | D2: Deviations | D3: Missing | D4: Measurement | D5: Reporting | Overall |
|---|---|---|---|---|---|---|
| Kolwelter_2023 | Low | Low | Some concerns | Low | Low | Low |
| Kosiborod 2023 | Low | Low | Some concerns | Low | Low | Some concerns |
| Kumar 2024 | Some concerns | Some concerns | Some concerns | Some concerns | Some concerns | Some concerns |
| Lee 2021 | Low | Low | Some concerns | Low | Some concerns | Some concerns |
| Lin 2024 | Some concerns | Low | Some concerns | Low | Some concerns | Some concerns |
| Lorenzo 2023a | Low | Low | Some concerns | Low | Low | Some concerns |
| Lorenzo 2023b | Low | Low | Low | Low | Low | Low |
| Maddukuri_2025 | Some concerns | Some concerns | Some concerns | Some concerns | Some concerns | High |
| Marton 2024 | Some concerns | Low | Some concerns | Low | Some concerns | Some concerns |
| McMurray 2024 | Low | Low | Low | Low | Low | Low |
| McMurray_2019 | Low | Low | Low | Low | Low | Low |
| Mocan 2025 | Some concerns | High | Low | Some concerns | Low | High |
| Mohebi 2024 | Some concerns | Some concerns | Some concerns | Low | Low | Some concerns |
| Montomoli 2024 | Some concerns | High | Some concerns | Some concerns | Some concerns | High |
| Mordi 2020 | Some concerns | Low | Some concerns | Low | Some concerns | Some concerns |
| Mordi 2021 | Low | Low | Some concerns | Low | Some concerns | Some concerns |
| Murakami 2016 | Some concerns | Some concerns | Some concerns | Some concerns | Some concerns | High |
| Murakami 2017 | Some concerns | High | Some concerns | Some concerns | Some concerns | High |
| NCT03030235 2017 | Some concerns | Some concerns | Some concerns | Some concerns | Some concerns | High |
| NCT03036124 2017 | Some concerns | Some concerns | Some concerns | Some concerns | Some concerns | High |
| NCT03057977 2017 | Some concerns | Some concerns | Some concerns | Some concerns | Some concerns | High |
| NCT03448419 | Low | Low | Some | Low | Low | Some |

| Study | D1: Randomization | D2: Deviations | D3: Missing | D4: Measurement | D5: Reporting | Overall |
|---|---|---|---|---|---|---|
| (EMPERIAL-Reduced) | | | concerns | | | concerns |
| NCT03521934 2018 | Some concerns | Some concerns | Some concerns | Some concerns | Some concerns | High |
| NCT03877237 2019 | Some concerns | Some concerns | Some concerns | Some concerns | Some concerns | High |
| NCT04157751 2019 | Some concerns | Some concerns | Some concerns | Some concerns | Some concerns | High |
| NCT04249778 | Some concerns | Some concerns | Some concerns | Some concerns | Some concerns | High |
| NCT04252287 2020 | Some concerns | Some concerns | Some concerns | Some concerns | Some concerns | High |
| NCT04869124 | Some concerns | Some concerns | Some concerns | Some concerns | Some concerns | High |
| NCT05152940 (ERTU-SODIUM) | Some concerns | Some concerns | Some concerns | Some concerns | Some concerns | High |
| NCT06012279 (CODA-HFrEF) | Some concerns | Some concerns | Some concerns | Some concerns | Some concerns | High |
| Naser_2024 | Low | Low | Low | Low | Some concerns | Low |
| Nassif 2019 | Low | Some concerns | Some concerns | Low | Low | Some concerns |
| Nassif 2021 | Low | Low | Low | Low | Low | Low |
| Nassif_2020 | Low | Low | Some concerns | Low | Some concerns | Some concerns |
| Nassif_2021 | Low | Low | Low | Low | Low | Low |
| Omar 2021 | Low | Low | Some concerns | Low | Some concerns | Some concerns |
| Omar_2022 | Low | Low | Some concerns | Low | Some concerns | Some concerns |
| Ovchinnikov 2021 | Some concerns | High | Some concerns | Some concerns | Some concerns | High |
| Ovchinnikov_2025 | Some concerns | Some concerns | Low | Some concerns | Low | Some concerns |
| Packer 2021 | Low | Low | Low | Low | Low | Low |
| Packer 2021 | Low | Low | Low | Low | Low | Low |
| Palau 2022 | Low | Low | Low | Low | Low | Low |
| Pastore 2024 | Some concerns | Some concerns | Some concerns | Some concerns | Some concerns | High |

| Study | D1: Randomization | D2: Deviations | D3: Missing | D4: Measurement | D5: Reporting | Overall |
|---|---|---|---|---|---|---|
| Pastore 2024b | Some concerns | High | Some concerns | Some concerns | Low | High |
| Polat_2025 | Some concerns | Some concerns | Some concerns | Low | Some concerns | High |
| Qin 2023 | Some concerns | Some concerns | Some concerns | Some concerns | Some concerns | High |
| Rau_2021 | Low | Low | Some concerns | Low | Low | Low |
| Reis 2022 | Some concerns | High | Some concerns | Some concerns | Low | High |
| Santos-Gallego 2021 | Some concerns | Low | Some concerns | Low | Some concerns | Some concerns |
| Schulze 2022 | Some concerns | Low | Some concerns | Low | Some concerns | Some concerns |
| Shirakabe_2020 | Some concerns | Some concerns | Some concerns | Some concerns | Some concerns | High |
| Shoshina 2026 | Some concerns | Some concerns | Some concerns | Low | Low | Some concerns |
| Singh 2018 | Low | Low | Low | Low | Low | Low |
| Singh 2019 | Low | Low | Some concerns | Low | Low | Some concerns |
| Solomon_2022 | Low | Low | Low | Low | Low | Low |
| Spinar 2020 | Low | Low | Low | Low | Low | Low |
| Spinar 2020 | Some concerns | Some concerns | Some concerns | Some concerns | Some concerns | High |
| Tada_2024 | Some concerns | Some concerns | Some concerns | Low | Some concerns | High |
| Tamaki 2019 | Some concerns | Some concerns | Some concerns | Some concerns | Some concerns | High |
| Tamaki 2021 | Some concerns | High | Some concerns | Some concerns | Some concerns | High |
| Tamaki_2020 | Some concerns | Some concerns | Some concerns | Some concerns | Some concerns | High |
| Tanaka 2020 | Low | High | Low | Some concerns | Low | High |
| Thiele 2022 | Low | Low | Some concerns | Low | Low | Some concerns |
| Ueda 2021 | Some concerns | Some concerns | Some concerns | Low | Some concerns | Some concerns |
| Voors 2022 | Low | Low | Low | Low | Low | Low |

| Study | D1: Randomization | D2: Deviations | D3: Missing | D4: Measurement | D5: Reporting | Overall |
|---|---|---|---|---|---|---|
| Wu 2022 | Some concerns | Some concerns | Some concerns | Some concerns | Some concerns | High |
| Wu 2022 | Some concerns | Some concerns | Some concerns | Some concerns | Some concerns | Some concerns |
| Xie 2023 | Some concerns | High | Some concerns | Some concerns | Some concerns | High |
| Xie 2024 | Some concerns | Some concerns | Some concerns | Some concerns | Some concerns | High |
| You 2025 | Some concerns | Some concerns | Some concerns | Some concerns | Some concerns | Some concerns |
| Zaoui 2022 | Some concerns | High | Some concerns | Some concerns | Some concerns | High |
| Zhou 2024 | Some concerns | Some concerns | Some concerns | Some concerns | Some concerns | Some concerns |
| de Boer 2020 | Low | Low | Some concerns | Low | Some concerns | Some concerns |

### eTable 3: Studies Excluded from Meta-Analysis (n = 55)

| Study | Record ID | Trial |
|---|---|---|
| Ali 2024 | CO-0729 | |
| Boorsma 2021 | CO-0725 | EMPA-RESPONSE-AHF |
| Bosch 2023 | CO-1540 | ELSI (post hoc) |
| Charaya 2022 | CO-1534 | |
| Charaya 2023 | CO-1533 | |
| EASTER-HF | CO-1273 | 10.1016/j.ihj.2024.06.009 |
| EUCTR2016-002280-34-DE 2016 | CO-1580 | EMPEROR-Reduced |
| EUCTR2021-005446-15-NL | CO-1952 | DAPA-severe-CKD |
| Ejiri 2019 | CO-0131 | MUSCAT-HF |
| Golubovskaya 2023 | CO-1734 | Golubovskaya 2023 |
| Golubovskaya 2025b | CO-1849 | Golubovskaya 2025b |
| Jones 2020 | CO-0989 | |
| Kawanami 2024 | CO-1744 | ROAD-ADHF |
| Kolwelter 2021b | CO-0609 | NCT03128528 |
| Kolwelter 2021c | CO-0687 | NCT03128528 |
| Kolwelter 2021d | CO-0715 | NCT03128528 |
| Kolwelter_2023 | CO-0465 | ELSI |
| Kosiborod 2023 | CO-0748 | SOLOIST-WHF |
| Kumar 2024 | CO-1850 | |
| Lorenzo 2023a | CO-0680 | DAPA-VO2 |
| Lorenzo 2023b | CO-0696 | DAPA-VO2 |
| Maddukuri_2025 | CO-0307 | |
| Mohebi 2024 | CO-0986 | CHIEF-HF |
| Montomoli 2024 | CO-1679 | DAPA-DP |
| Mordi 2021 | CO-0621 | RECEDE-CHF |
| Murakami 2016 | CO-0635 | Murakami 2016 |
| Murakami 2017 | CO-1747 | Murakami 2017 |
| NCT03030235 2017 | CO-1021 | PRESERVED-HF |
| NCT03036124 2017 | CO-1188 | DAPA-HF |

| Study | Record ID | Trial |
|---|---|---|
| NCT03057977 2017 | CO-1032 | EMPEROR-Reduced |
| NCT03521934 2018 | CO-1485 | SOLOIST-WHF |
| NCT03877237 2019 | CO-1029 | DETERMINE-reduced |
| NCT04157751 2019 | CO-0743 | EMPULSE |
| NCT04249778 | CO-0172 | DAPA-Discharge |
| NCT04252287 2020 | CO-1481 | CHIEF-HF |
| NCT04869124 | CO-1334 | DAPA-Volume |
| NCT05152940 (ERTU-SODIUM) | CO-0617 | ERTU-SODIUM |
| NCT06012279 (CODA-HFrEF) | CO-0151 | CODA-HFrEF |
| Naser_2024 | CO-0287 | CAMEO-DAPA (secondary) |
| Omar 2021 | CO-1140 | Omar 2021 (post hoc) |
| Packer 2021 | CO-1064 | EMPEROR-Preserved (WHF) |
| Pastore 2024b | CO-1040 | DAPA ECHO |
| Polat_2025 | CO-0368 | CO-0368 |
| Qin 2023 | CO-0645 | Qin 2023 |
| Singh 2018 | CO-0802 | |
| Singh 2019 | CO-0800 | REFORM |
| Spinar 2020 | CO-0786 | EMPEROR-Reduced |
| Spinar 2020 | CO-0730 | EMPEROR-Reduced |
| Tada_2024 | CO-0285 | |
| Tamaki 2019 | CO-0637 | Tamaki 2019 |
| Tamaki_2020 | CO-0264 | Tamaki 2020 |
| Wu 2022 | CO-0672 | |
| Xie 2023 | CO-1634 | DAHOS |
| You 2025 | CO-1724 | |
| Zaoui 2022 | WH-0205 | 10.1016/j.ancard.2022.08.007 |

### eTable 4: Primary Outcomes — Meta-Analysis Results

| outcome | measure | k | estimate | ci_low | ci_high | p_value | I2 | tau2 |
|---|---|---|---|---|---|---|---|---|
| acm | RR | 26 | 0.8987962 | 0.8256442 | 0.9784294 | 0.0158320409593 | 0.00000 | 0.002728075 |
| acm_hr | HR | 4 | 0.9182126 | 0.7957675 | 1.0594983 | 0.1540394400245 | 17.15995 | 0.002000254 |
| hfh | RR | 15 | 0.7382688 | 0.6893397 | 0.7906709 | 0.0000001777259 | 0.00000 | 0.000000000 |
| hfh_hr | HR | 4 | 0.7206113 | 0.6631949 | 0.7829986 | 0.0010885082781 | 0.00000 | 0.000000000 |

### eTable 5: Secondary Outcomes — Meta-Analysis Results

| outcome | measure | k | estimate | ci_low | ci_high | p_value | I2 | tau2 |
|---|---|---|---|---|---|---|---|---|
| cvd | RR | 7 | 0.86102517 | 0.7555638 | 0.98120686 | 0.031073443740 | 18.42353 | 0.0000000000000 |
| cvd_hr | HR | 4 | 0.86645652 | 0.8011837 | 0.93704718 | 0.010079354541 | 0.00000 | 0.0000000000000 |
| composite | RR | 8 | 0.79565821 | 0.7454168 | 0.84928587 | 0.000072739352 | 0.00000 | 0.0000003427684 |
| composite_hr | HR | 8 | 0.76678035 | 0.7266839 | 0.80908921 | 0.000007571207 | 0.00000 | 0.0000000000000 |
| sae | RR | 20 | 0.94079613 | 0.8957819 | 0.98807235 | 0.017388676975 | 36.10769 | 0.0000080760710 |
| kccq_tss | MD | 9 | 2.61116051 | 1.2251221 | 3.99719896 | 0.002464138391 | 49.90787 | 0.9166396189804 |
| kccq_oss | MD | 3 | 2.79055862 | -1.9488596 | 7.52997681 | 0.126835634305 | 25.22881 | 0.9816244410873 |
| sixmwd | MD | 8 | 3.90532132 | -6.5416808 | 14.35232348 | 0.406058264635 | 68.16114 | 103.0430681359222 |
| ntprobnp | MD | 8 | -29.98859922 | -89.0067617 | 29.02956327 | 0.268616981229 | 66.12065 | 2,098.0954041795335 |
| egfr | MD | 3 | -0.08218558 | -2.2666106 | 2.10223942 | 0.886275753704 | 0.00000 | 0.0000035397612 |
| weight | MD | 9 | -1.27284257 | -2.5320017 | -0.01368345 | 0.048081774451 | 84.48348 | 2.0104206995819 |

| outcome | measure | k | estimate | ci_low | ci_high | p_value | I2 | tau2 |
|---|---|---|---|---|---|---|---|---|
| sbp | MD | 8 | -1.33279461 | -4.8359170 | 2.17032778 | 0.398201095113 | 62.12428 | 9.6084822306209 |

### eTable 6: Sensitivity Analyses — All-Cause Mortality

| analysis | k | estimate | ci_low | ci_high | p_value | I2 | tau2 |
|---|---|---|---|---|---|---|---|
| Main analysis | 26 | 0.8987962 | 0.8256442 | 0.9784294 | 0.015832041 | 0.00000 | 0.002728075 |
| Excluding high RoB | 20 | 0.9111841 | 0.8474686 | 0.9796899 | 0.014640718 | 0.00000 | 0.001545973 |
| Low RoB only | 13 | 0.9191598 | 0.8425783 | 1.0027017 | 0.056407300 | 0.00000 | 0.001814616 |
| Excluding open-label | 18 | 0.9143255 | 0.8537652 | 0.9791816 | 0.013454471 | 0.00000 | 0.001309919 |
| HFrEF only | 7 | 0.8386478 | 0.7587114 | 0.9270060 | 0.005101605 | 0.00000 | 0.000000000 |
| HFpEF only | 4 | 0.9815415 | 0.8200276 | 1.1748674 | 0.763242754 | 0.00000 | 0.000000000 |
| Follow-up >= 12 months | 6 | 0.9262442 | 0.8272375 | 1.0371004 | 0.141938240 | 17.17701 | 0.001121536 |
| N >= 100 | 18 | 0.9155031 | 0.8547979 | 0.9805193 | 0.014713521 | 0.00000 | 0.001120958 |
| N >= 50 | 25 | 0.8983609 | 0.8234066 | 0.9801383 | 0.018008764 | 0.00000 | 0.002787461 |
| Empagliflozin only | 9 | 0.6801095 | 0.3687329 | 1.2544281 | 0.184528148 | 40.38563 | 0.217750331 |
| Dapagliflozin only | 11 | 0.8949349 | 0.8182483 | 0.9788086 | 0.020099713 | 0.00000 | 0.002324827 |
| Excluding imputed data | 20 | 0.8036026 | 0.6215517 | 1.0389756 | 0.090823766 | 0.00000 | 0.038860396 |
| Excluding sotagliflozin | 25 | 0.8961698 | 0.8136503 | 0.9870585 | 0.027797972 | 0.00000 | 0.004039608 |

### eTable 7: Alternative Sensitivity Analyses — All-Cause Mortality

| analysis | k | estimate | ci_low | ci_high | p_value | I2 | tau2 |
|---|---|---|---|---|---|---|---|
| Fixed-effect (MH-RR) | 26 | 0.9056997 | 0.8452130 | 0.9705150 | 0.004975388 | 0.00000 | 0.002728075 |
| Random-effects OR | 26 | 0.8801194 | 0.7960227 | 0.9731006 | 0.014779370 | 0.00000 | 0.003783542 |
| Fixed-effect OR | 26 | 0.8910903 | 0.8222066 | 0.9657451 | 0.004968284 | 0.00000 | 0.003783542 |
| Peto OR | 26 | 0.8908173 | 0.8219194 | 0.9654907 | 0.004877004 | 18.88036 | 0.008635657 |
| Paule-Mandel RR | 26 | 0.9125498 | 0.8513197 | 0.9781839 | 0.011877743 | 0.00000 | 0.000000000 |

### eTable 8: Sensitivity Analyses — HF Hospitalization

| analysis | k | estimate | ci_low | ci_high | p_value | I2 | tau2 |
|---|---|---|---|---|---|---|---|
| Main analysis | 15 | 0.7382688 | 0.6893397 | 0.7906709 | 0.0000001777259 | 0.00000 | 0.000000000000 |
| Excluding high RoB | 11 | 0.7412384 | 0.6904666 | 0.7957435 | 0.0000027865122 | 0.00000 | 0.000000000000 |
| Low RoB only | 8 | 0.7418306 | 0.6934925 | 0.7935380 | 0.0000156917720 | 0.00000 | 0.000000000000 |
| Excluding open-label | 10 | 0.7364540 | 0.6815410 | 0.7957915 | 0.0000090982654 | 0.00000 | 0.000000000000 |
| HFrEF only | 5 | 0.7214719 | 0.6472437 | 0.8042129 | 0.0011253150487 | 0.00000 | 0.000000000000 |
| HFpEF only | 2 | 0.7349113 | 0.6119139 | 0.8826316 | 0.0297724885230 | 0.00000 | 0.000000000000 |
| Follow-up >= 12 months | 6 | 0.7420861 | 0.6820718 | 0.8073808 | 0.0002692598387 | 0.00000 | 0.000006553957 |
| N >= 100 | 10 | 0.7407016 | 0.6883452 | 0.7970403 | 0.0000067451541 | 0.00000 | 0.000000000000 |
| N >= 50 | 14 | 0.7379380 | 0.6870926 | 0.7925459 | 0.0000004713707 | 0.00000 | 0.000000000000 |
| Empagliflozin only | 4 | 0.7258541 | 0.6673937 | 0.7894354 | 0.0012020939583 | 0.00000 | 0.000000000000 |
| Dapagliflozin only | 7 | 0.7485689 | 0.6550936 | 0.8553821 | 0.0018081101751 | 15.20945 | 0.000000000000 |
| Excluding imputed data | 10 | 0.7240112 | 0.5526339 | 0.9485343 | 0.0242080383431 | 1.79611 | 0.026190607820 |
| Excluding sotagliflozin | 15 | 0.7382688 | 0.6893397 | 0.7906709 | 0.0000001777259 | 0.00000 | 0.000000000000 |

### eTable 9: Alternative Sensitivity Analyses — HF Hospitalization

| analysis | k | estimate | ci_low | ci_high | p_value | I2 | tau2 |
|---|---|---|---|---|---|---|---|
| Fixed-effect (MH-RR) | 15 | 0.7356725 | 0.6850310 | 0.7900578 | 0.0000000000000000003289204 | 0 | 0 |
| Random-effects OR | 15 | 0.7061234 | 0.6529543 | 0.7636219 | 0.000000016824696225715034 | 0 | 0 |
| Fixed-effect OR | 15 | 0.7041798 | 0.6491843 | 0.7638344 | 0.0000000000000000002830942 | 0 | 0 |
| Peto OR | 15 | 0.7054893 | 0.6508979 | 0.7646593 | 0.0000000000000000002069455 | 0 | 0 |
| Paule-Mandel RR | 15 | 0.7382688 | 0.6893397 | 0.7906709 | 0.00000017772590218207810 | 0 | 0 |

### eTable 10: Subgroup Analyses — All-Cause Mortality

| outcome | subgroup_var | subgroup | k | estimate | ci_low | ci_high | p_value | I2 | interaction_p |
|---|---|---|---|---|---|---|---|---|---|
| ACM | hf_group | HFrEF | 7 | 0.8386478 | 0.758711448 | 0.9270060 | 0.005101605 | 0.0000000 | 0.038560561 |
| ACM | hf_group | HFpEF | 4 | 0.9815415 | 0.820027637 | 1.1748674 | 0.763242754 | 0.0000000 | 0.038560561 |
| ACM | hf_group | HFmrEF | 1 | 0.1473566 | 0.007766666 | 2.7957893 | 0.202214064 | | 0.038560561 |
| ACM | hf_group | Overall | 12 | 0.9019184 | 0.792823654 | 1.0260249 | 0.105732726 | 0.0000000 | 0.038560561 |
| ACM | drug_class | Empagliflozin | 9 | 0.6801095 | 0.368732920 | 1.2544281 | 0.184528148 | 40.3856327 | 0.246634598 |
| ACM | drug_class | Dapagliflozin | 11 | 0.8949349 | 0.818248321 | 0.9788086 | 0.020099713 | 0.0000000 | 0.246634598 |
| ACM | drug_class | Other | 3 | 0.7322428 | 0.171001488 | 3.1355253 | 0.453891338 | 0.0000000 | 0.246634598 |
| ACM | drug_class | Canagliflozin | 2 | 0.4118703 | 0.003304750 | 51.3313043 | 0.257519655 | 0.0000000 | 0.246634598 |
| ACM | drug_class | Sotagliflozin | 1 | 0.8637033 | 0.632492448 | 1.1794343 | 0.356651622 | | 0.246634598 |
| ACM | drug_class | Overall | 26 | 0.8987962 | 0.825644212 | 0.9784294 | 0.015832041 | 0.0000000 | 0.246634598 |
| ACM | blinding | Double-blind | 18 | 0.9143255 | 0.853765161 | 0.9791816 | 0.013454471 | 0.0000000 | 0.142108969 |
| ACM | blinding | Open-label | 7 | 0.5676610 | 0.264580218 | 1.2179255 | 0.119448601 | 20.3489022 | 0.142108969 |
| ACM | blinding | Single-blind | 1 | 0.1371994 | 0.007275888 | 2.5871322 | 0.184972157 | | 0.142108969 |
| ACM | blinding | Overall | 26 | 0.8987962 | 0.825644212 | 0.9784294 | 0.015832041 | 0.0000000 | 0.142108969 |
| ACM | fu_group | <12 months | 20 | 0.7137838 | 0.550910620 | 0.9248094 | 0.013452357 | 0.0000000 | 0.047256414 |
| ACM | fu_group | ≥12 months | 6 | 0.9262442 | 0.827237485 | 1.0371004 | 0.141938240 | 17.1770079 | 0.047256414 |
| ACM | fu_group | Overall | 26 | 0.8987962 | 0.825644212 | 0.9784294 | 0.015832041 | 0.0000000 | 0.047256414 |
| ACM | size_group | 100-999 | 14 | 0.7156878 | 0.527849531 | 0.9703694 | 0.033694940 | 0.0000000 | 0.001524733 |
| ACM | size_group | <100 | 8 | 0.3445564 | 0.166072626 | 0.7148628 | 0.010660059 | 0.0000000 | 0.001524733 |

| outcome | subgroup_var | subgroup | k | estimate | ci_low | ci_high | p_value | I2 | interaction_p |
|---|---|---|---|---|---|---|---|---|---|
| ACM | size_group | ≥1000 | 4 | 0.9270183 | 0.824910712 | 1.0417649 | 0.130663112 | 0.1626839 | 0.001524733 |
| ACM | size_group | Overall | 26 | 0.8987962 | 0.825644212 | 0.9784294 | 0.015832041 | 0.0000000 | 0.001524733 |
| ACM | overall_judgment | Low | 13 | 0.9191598 | 0.842578321 | 1.0027017 | 0.056407300 | 0.0000000 | 0.251170553 |
| ACM | overall_judgment | High | 6 | 0.5976530 | 0.253058746 | 1.4114868 | 0.184259217 | 28.7294542 | 0.251170553 |
| ACM | overall_judgment | Some concerns | 7 | 0.8166131 | 0.642620011 | 1.0377159 | 0.084021791 | 0.0000000 | 0.251170553 |
| ACM | overall_judgment | Overall | 26 | 0.8987962 | 0.825644212 | 0.9784294 | 0.015832041 | 0.0000000 | 0.251170553 |
| ACM | era | 2022+ | 13 | 0.6962960 | 0.501127123 | 0.9674753 | 0.033650569 | 15.7467899 | 0.008939765 |
| ACM | era | 2020-2021 | 10 | 0.9648090 | 0.864250548 | 1.0770677 | 0.480301131 | 0.0000000 | 0.008939765 |
| ACM | era | Pre-2020 | 3 | 0.8373373 | 0.785279206 | 0.8928464 | 0.006987527 | 0.0000000 | 0.008939765 |
| ACM | era | Overall | 26 | 0.8987962 | 0.825644212 | 0.9784294 | 0.015832041 | 0.0000000 | 0.008939765 |

### eTable 11: Subgroup Analyses — HF Hospitalization

| outcome | subgroup_var | subgroup | k | estimate | ci_low | ci_high | p_value | I2 | interaction_p |
|---|---|---|---|---|---|---|---|---|---|
| HFH | hf_group | HFrEF | 5 | 0.7214719 | 0.647243683 | 0.8042129 | 0.0011253150487 | 0.000000 | 0.13161321 |
| HFH | hf_group | HFpEF | 2 | 0.7349113 | 0.611913943 | 0.8826316 | 0.0297724885230 | 0.000000 | 0.13161321 |
| HFH | hf_group | HFmrEF | 2 | 0.5060828 | 0.046266652 | 5.5357333 | 0.1717040798079 | 0.000000 | 0.13161321 |
| HFH | hf_group | Overall | 9 | 0.7160758 | 0.659592737 | 0.7773957 | 0.0000137329524 | 0.000000 | 0.13161321 |
| HFH | drug_class | Empagliflozin | 4 | 0.7258541 | 0.667393687 | 0.7894354 | 0.0012020939583 | 0.000000 | 0.42383237 |
| HFH | drug_class | Canagliflozin | 3 | 1.2701421 | 0.170221560 | 9.4774187 | 0.6596194392910 | 0.000000 | 0.42383237 |
| HFH | drug_class | Other | 1 | 0.1473566 | 0.007766666 | 2.7957893 | 0.2022140639856 |  | 0.42383237 |
| HFH | drug_class | Dapagliflozin | 7 | 0.7485689 | 0.655093576 | 0.8553821 | 0.0018081101751 | 15.209448 | 0.42383237 |
| HFH | drug_class | Overall | 15 | 0.7382688 | 0.689339695 | 0.7906709 | 0.0000001777259 | 0.000000 | 0.42383237 |
| HFH | blinding | Double-blind | 10 | 0.7364540 | 0.681541002 | 0.7957915 | 0.0000090982654 | 0.000000 | 0.01825964 |
| HFH | blinding | Open-label | 4 | 0.9674451 | 0.696692553 | 1.3434189 | 0.7693996199073 | 0.000000 | 0.01825964 |
| HFH | blinding | Single-blind | 1 | 0.2742857 | 0.059945180 | 1.2550242 | 0.0954743198390 |  | 0.01825964 |
| HFH | blinding | Overall | 15 | 0.7382688 | 0.689339695 | 0.7906709 | 0.0000001777259 | 0.000000 | 0.01825964 |
| HFH | fu_group | <12 months | 9 | 0.6987368 | 0.509454485 | 0.9583449 | 0.0308146914941 | 0.000000 | 0.66919154 |
| HFH | fu_group | ≥12 months | 6 | 0.7420861 | 0.682071847 | 0.8073808 | 0.0002692598387 | 0.000000 | 0.66919154 |
| HFH | fu_group | Overall | 15 | 0.7382688 | 0.689339695 | 0.7906709 | 0.0000001777259 | 0.000000 | 0.66919154 |
| HFH | size_group | 100-999 | 6 | 0.7014878 | 0.455693918 | 1.0798590 | 0.0883130065783 | 14.368275 | 0.24603032 |
| HFH | size_group | <100 | 5 | 0.4079821 | 0.147553928 | 1.1280583 | 0.0706358363085 | 0.000000 | 0.24603032 |
| HFH | size_group | ≥1000 | 4 | 0.7446148 | 0.695717499 | 0.7969488 | 0.0008206206835 | 0.000000 | 0.24603032 |

| outcome | subgroup_var | subgroup | k | estimate | ci_low | ci_high | p_value | I2 | interaction_p |
|---|---|---|---|---|---|---|---|---|---|
| HFH | size_group | Overall | 15 | 0.7382688 | 0.689339695 | 0.7906709 | 0.0000001777259 | 0.000000 | 0.24603032 |
| HFH | overall_judgment | Low | 8 | 0.7418306 | 0.693492486 | 0.7935380 | 0.0000156917720 | 0.000000 | 0.95999020 |
| HFH | overall_judgment | High | 4 | 0.7211758 | 0.397256054 | 1.3092174 | 0.1794170951304 | 5.734242 | 0.95999020 |
| HFH | overall_judgment | Some concerns | 3 | 0.6291579 | 0.034865356 | 11.3533811 | 0.5619235360300 | 36.284054 | 0.95999020 |
| HFH | overall_judgment | Overall | 15 | 0.7382688 | 0.689339695 | 0.7906709 | 0.0000001777259 | 0.000000 | 0.95999020 |
| HFH | era | 2022+ | 8 | 0.7493744 | 0.610054859 | 0.9205107 | 0.0128211502122 | 14.780798 | 0.93698738 |
| HFH | era | 2020-2021 | 6 | 0.7244695 | 0.661494273 | 0.7934400 | 0.0002666807493 | 0.000000 | 0.93698738 |
| HFH | era | Pre-2020 | 1 | 0.7263811 | 0.620634945 | 0.8501448 | 0.0000682157304 |  | 0.93698738 |
| HFH | era | Overall | 15 | 0.7382688 | 0.689339695 | 0.7906709 | 0.0000001777259 | 0.000000 | 0.93698738 |

### eTable 12: Publication Bias Assessment

| outcome | k | egger_p | egger_sig | tf_k_added | tf_estimate | tf_ci_low | tf_ci_high | tf_p |
|---|---|---|---|---|---|---|---|---|
| ACM (RR) | 26 | 0.02759532 | true | 6 | 0.9169688 | 0.8406845 | 1.0001752 | 0.05043603026219 |
| HFH (RR) | 15 | 0.18854637 | false | 3 | 0.7402372 | 0.6906137 | 0.7934264 | 0.00000005640258 |
| CVD (RR) | 7 | | | | | | | |
| Composite (RR) | 8 | | | | | | | |
| SAE (RR) | 20 | 0.43072281 | false | 2 | 0.9418447 | 0.8955244 | 0.9905609 | 0.02213314393173 |
| NT-proBNP | 8 | | | | | | | |
| eGFR | 3 | | | | | | | |
| SBP | 8 | | | | | | | |

### eTable 13: Heterogeneity Assessment

| outcome | k | Q | Q_df | Q_p | I2 | tau2 | tau | predict_low | predict_high | I2_class |
|---|---|---|---|---|---|---|---|---|---|---|
| ACM (RR) | 26 | 22.699039 | 25 | 0.59513257986456 | 0.00000 | 0.0027280748385 | 0.0522309759 | 0.78015508 | 1.0354795 | Low |
| ACM (HR) | 4 | 3.621437 | 3 | 0.30535032131991 | 17.15995 | 0.0020002540030 | 0.0447241993 | 0.74719966 | 1.1283655 | Low |
| HFH (RR) | 15 | 10.781082 | 14 | 0.70314173500638 | 0.00000 | 0.0000000000000 | 0.0000000000 | 0.68277461 | 0.7982734 | Low |
| HFH (HR) | 4 | 1.269008 | 3 | 0.73650513800710 | 0.00000 | 0.0000000000000 | 0.0000000000 | 0.63424531 | 0.8187379 | Low |
| CVD (RR) | 7 | 7.355062 | 6 | 0.28925525088228 | 18.42353 | 0.0000000000000 | 0.0000000000 | 0.76518100 | 0.9688745 | Low |
| CVD (HR) | 4 | 0.720058 | 3 | 0.86847627177707 | 0.00000 | 0.0000000000000 | 0.0000000000 | 0.73844369 | 1.0166610 | Low |
| Composite (RR) | 8 | 6.631423 | 7 | 0.46823694241927 | 0.00000 | 0.0000003427684 | 0.0005854643 | 0.74407384 | 0.8508188 | Low |
| Composite | 8 | 3.953890 | 7 | 0.78507284965291 | 0.00000 | 0.0000000000000 | 0.0000000000 | 0.71389596 | 0.8235823 | Low |

| outcome | k | Q | Q_df | Q_p | I2 | tau2 | tau | predict_low | predict_high | I2_class |
|---|---|---|---|---|---|---|---|---|---|---|
| (HR) | | | | | | | | | | |
| SAE (RR) | 20 | 29.737536 | 19 | 0.05523518048392 | 36.10769 | 0.0000080760710 | 0.0028418429 | 0.90422819 | 0.9788429 | Low-moderate |
| KCCQ-TSS | 9 | 15.970573 | 8 | 0.04280323846921 | 49.90787 | 0.9166396189804 | 0.9574129825 | 0.07148469 | 5.1508363 | Low-moderate |
| KCCQ-OSS | 3 | 2.674827 | 2 | 0.26252382800675 | 25.22881 | 0.9816244410873 | 0.9907696206 | -3.62511439 | 9.2062316 | Low-moderate |
| 6MWD | 8 | 21.985714 | 7 | 0.00255486037527 | 68.16114 | 103.0430681359222 | 10.1510131581 | -22.26630744 | 30.0769501 | Moderate-high |
| NT-proBNP | 8 | 20.661552 | 7 | 0.00430473382068 | 66.12065 | 2,098.0954041795335 | 45.8049713915 | -149.68123765 | 89.7040392 | Moderate-high |
| eGFR | 3 | 1.698493 | 2 | 0.42773720568483 | 0.00000 | 0.0000035397612 | 0.0018814253 | -2.45259288 | 2.2882217 | Low |
| Weight | 9 | 51.557947 | 8 | 0.00000002048437 | 84.48348 | 2.0104206995819 | 1.4178930494 | -4.75165634 | 2.2059712 | High |
| SBP | 8 | 18.481499 | 7 | 0.00997651713723 | 62.12428 | 9.6084822306209 | 3.0997551888 | -9.38807140 | 6.7224822 | Moderate-high |

### eTable 14: Safety Outcomes

| outcome | k | rr | ci_low | ci_high | p_value | I2 |
|---|---|---|---|---|---|---|
| dka | 4 | 1.4726734 | 0.09933338 | 21.833212 | 0.67882126 | 13.89464 |
| aki | 13 | 0.8871977 | 0.62559251 | 1.258199 | 0.46978398 | 24.40960 |
| uti | 13 | 1.0070244 | 0.53872431 | 1.882406 | 0.98094975 | 0.00000 |
| hypotension | 8 | 0.9910533 | 0.68830645 | 1.426961 | 0.95514232 | 0.00000 |
| genital_inf | 6 | 3.7500701 | 1.71743549 | 8.188386 | 0.00735361 | 0.00000 |

### eTable 15: GRADE Summary of Findings

| outcome | k | certainty | downgrades | comments |
|---|---|---|---|---|
| ACM (RR) | 26 | Low | Risk of bias (-1); Publication bias (-1) | 31% high RoB; Egger p = 0.028 |
| HFH (RR) | 15 | Moderate | Risk of bias (-1) | 31% high RoB |
| CVD (RR) | 7 | Moderate | Risk of bias (-1) | 30% high RoB |
| Composite (RR) | 8 | High | | |
| SAE (RR) | 20 | High | | |
| KCCQ-TSS | 9 | High | | |
| KCCQ-OSS | 3 | Moderate | Imprecision (-1) | |
| 6MWD | 8 | Low | Inconsistency (-1); Imprecision (-1) | $I^2$ = 68% |
| NT-proBNP | 8 | Very low | Risk of bias (-1); Inconsistency (-1); Imprecision (-1) | 38% high RoB; $I^2$ = 66% |
| eGFR | 3 | Low | Risk of bias (-1); Imprecision (-1) | 33% high RoB |
| Weight | 9 | Moderate | Inconsistency (-1) | $I^2$ = 84% |
| SBP | 8 | Very low | Risk of bias (-1); Inconsistency (-1); Imprecision (-1) | 38% high RoB; $I^2$ = 62% |

### eTable 16: Number Needed to Treat (NNT)

| outcome | cer | rr | arr | nnt | direction | wmean_fu_weeks | wmean_fu_months |
|---|---|---|---|---|---|---|---|
| ACM | 0.12831858 | 0.8987962 | 0.01298633 | 78 | benefit | 85.70290 | 19.72449 |
| HFH | 0.13524519 | 0.7382688 | 0.03539789 | 29 | benefit | 91.11139 | 20.96925 |
| CVD | 0.0889861 | 0.861025 | 0.0123668 | 81 | benefit | 97.73298 | 22.49321 |

| outcome | cer | rr | arr | nnt | direction | wmean_fu_weeks | wmean_fu_months |
|---|---|---|---|---|---|---|---|
| | 4 | 2 | 3 | | | | |
| Composite | 0.20726942 | 0.7956582 | 0.04235380 | 24 | benefit | 95.25676 | 21.92331 |
| SAE | 0.40446774 | 0.9407961 | 0.02394606 | 42 | benefit | 93.66641 | 21.55729 |

### eTable 17: Trial Sequential Analysis

| outcome | rrr | cer | rr_target | I2 | ris_fixed | ris_adjusted | n_current | pct_ris | ris_reached |
|---|---|---|---|---|---|---|---|---|---|
| ACM | 0.15 | 0.1283186 | 0.85 | 0 | 10,081 | 10,081 | 22,606 | 224.2436 | true |
| HFH | 0.15 | 0.1352452 | 0.85 | 0 | 9,565 | 9,565 | 23,079 | 241.2859 | true |
| ACM | 0.20 | 0.1283186 | 0.80 | 0 | 5,528 | 5,528 | 22,606 | 408.9363 | true |
| HFH | 0.20 | 0.1352452 | 0.80 | 0 | 5,245 | 5,245 | 23,079 | 440.0191 | true |

### eTable 18: Data Imputations

| record_id | study_label | imputation_notes |
|---|---|---|
| CO-0064 | Anker 2021 | eGFR slope reported as annual rate not absolute change; egfr_change_change_sd_int=NEEDS_IMPUTATION(n=2997); egfr_change_change_sd_ctrl=NEEDS_IMPUTATION(n=2991); egfr_change_change_sd_int=NEEDS_IMPUTATION(n=2997); egfr_change_change_sd_ctrl=NEEDS_IMPUTATION(n=2991); egfr_change_change_sd_int=NEEDS_IMPUTATION(n=2997); egfr_change_change_sd_ctrl=NEEDS_IMPUTATION(n=2991) |
| CO-0450 | Bhatt_2021 | NR; age_mean=Wan(median,IQR); age_sd=IQR/1.35; weight_change_change_sd_ctrl=NEEDS_IMPUTATION(n=614); sbp_change_change_sd_int=NEEDS_IMPUTATION(n=608); weight_change_change_sd_ctrl=NEEDS_IMPUTATION(n=614); sbp_change_change_sd_int=NEEDS_IMPUTATION(n=608); weight_change_change_sd_ctrl=NEEDS_IMPUTATION(n=614); sbp_change_change_sd_int=NEEDS_IMPUTATION(n=608); age_mean=Wan(median,IQR); age_sd=IQR/1.35 |
| CO-0399 | EFFORT | NT-proBNP change is mean not median; NT-proBNP diff P=0.22 (NS); egfr_change_change_sd_int=NEEDS_IMPUTATION(n=65); egfr_change_change_sd_int=NEEDS_IMPUTATION(n=65); egfr_change_change_sd_int=NEEDS_IMPUTATION(n=65); egfr_change_change_sd_int=NEEDS_IMPUTATION(n=65) |
| CO-0029 | Ejiri 2020 | NR; age_mean=Wan(median,IQR); age_sd=IQR/1.35; egfr_change_change_sd_ctrl=NEEDS_IMPUTATION(n=83); sbp_change_change_sd_int=NEEDS_IMPUTATION(n=86); egfr_change_change_sd_ctrl=NEEDS_IMPUTATION(n=83); sbp_change_change_sd_int=NEEDS_IMPUTATION(n=86); egfr_change_change_sd_ctrl=NEEDS_IMPUTATION(n=83); sbp_change_change_sd_int=NEEDS_IMPUTATION(n=86) |
| CO-0839 | Jensen 2020 | ACM; HFH; CVD; composite; SAE; 6MWD; eGFR; baseline diabetes%/AF%/medications |
| CO-0432 | McMurray_2019 | NT-proBNP change reported as median; KCCQ-TSS as win ratio not mean change; sixmwd_change_sd_int=NEEDS_IMPUTATION(n=2373); egfr_change_change_sd_int=NEEDS_IMPUTATION(n=2373); weight_change_change_sd_ctrl=NEEDS_IMPUTATION(n=2371); sixmwd_change_sd_int=NEEDS_IMPUTATION(n=2373); egfr_change_change_sd_int=NEEDS_IMPUTATION(n=2373); weight_change_change_sd_ctrl=NEEDS_IMPUTATION(n=2371); sixmwd_change_sd_int=NEEDS_IMPUTATION(n=2373); egfr_change_change_sd_int=NEEDS_IMPUTATION(n=2373); weight_change_change_sd_ctrl=NEEDS_IMPUTATION(n=2371) |
| CO-0408 | Nassif_2021 | age_sd=IQR/1.35; sixmwd_change_sd_int=NEEDS_IMPUTATION(n=162); sixmwd_change_sd_int=NEEDS_IMPUTATION(n=162); sixmwd_change_sd_int=NEEDS_IMPUTATION(n=162); sixmwd_change_sd_int=NEEDS_IMPUTATION(n=162); age_sd=IQR/1.35 |
| CO-1013 | Packer 2021 | Circulation companion paper on worsening HF events; primary NEJM 2020 paper (Packer NEJM 383:1413-24) not available as separate PDF. HFH events=first adjudicated HFH. Composite=ACM or CV hosp. ACM HR from NEJM ref: 0.82 (0.59-1.14). HFH HR=0.69 (0.59-0.81) time-to-first. |

| record_id | study_label | imputation_notes |
|---|---|---|
| CO-0028 | Voors 2022 | PDF; age_mean=Wan(median,IQR); age_sd=IQR/1.35; kccq_tss_change_sd_int=NEEDS_IMPUTATION(n=265); kccq_tss_change_sd_ctrl=NEEDS_IMPUTATION(n=265); kccq_tss_change_sd_int=NEEDS_IMPUTATION(n=265); kccq_tss_change_sd_ctrl=NEEDS_IMPUTATION(n=265); kccq_tss_change_sd_int=NEEDS_IMPUTATION(n=265); kccq_tss_change_sd_ctrl=NEEDS_IMPUTATION(n=265); age_mean=Wan(median,IQR); age_sd=IQR/1.35 |
| WH-0025 | Wu 2022 | age_mean=Wan(median,IQR); age_sd=IQR/1.35; age_sd=IQR/1.35 |

### eAppendix 2: Screening Pipeline — Methodological Details

The study selection process comprised the following sequential steps:

**Step 1 — Automated deduplication.** Records from all four databases were combined and deduplicated using exact matching on persistent identifiers (PMID, DOI, NCT/registry ID) followed by fuzzy title matching (Levenshtein similarity threshold 95%). Candidate duplicate pairs below the threshold were flagged for manual review.

**Step 2 — Title and abstract screening.** A four-tier exclusion hierarchy was applied: (T1) animal or in vitro studies, (T2) wrong intervention (not SGLT2 inhibitor), (T3) wrong population (not heart failure), (T4) wrong design (not RCT). A key trial protection list of 12 sentinel trials prevented false exclusion of known eligible studies. Initial classification was facilitated by a large language model (Claude, Anthropic) to categorize records as likely eligible, likely ineligible, or uncertain. All uncertain records and a random sample of likely ineligible records were independently reviewed by both authors. Records classified as likely eligible proceeded to full-text assessment.

**Step 3 — Full-text screening (8 passes).**

1. *Automated eligibility check*: Regex-based screening of titles and abstracts against exclusion patterns (reviews, protocols, editorials, observational designs).
2. *API enrichment*: PubMed and ClinicalTrials.gov APIs queried to retrieve publication type, study design, and registry status for each record.
3. *Reviewer assessment*: Both reviewers independently assessed full-text reports in blocks of 25 records, applying PICO-based inclusion criteria. Disagreements resolved by consensus.
4. *Trial clustering*: Records grouped by NCT/registry ID and parent trial name to identify companion publications, sub-analyses, and post-hoc analyses.
5. *Sub-analysis exclusion*: Records meeting all three conditions — (a) title mentions a known parent trial, (b) title contains a sub-analysis indicator, (c) record is not the primary publication — were excluded (code FT06).
6. *Block verification*: Remaining records verified in blocks of 25 with independent classification by both reviewers (VERIFIED_INCLUDE, EXCLUDE, FLAG).
7. *Full-text retrieval*: PDFs obtained for all provisionally included records; records without obtainable full text were flagged.
8. *PICO verification*: Final verification of population, intervention, comparator, and outcomes against eligibility criteria using the full-text PDF.

All exclusion codes, decision logs, and the complete reproducible pipeline (Python scripts) are available in the data repository.

### eFigure 1: PRISMA 2020 Flow Diagram

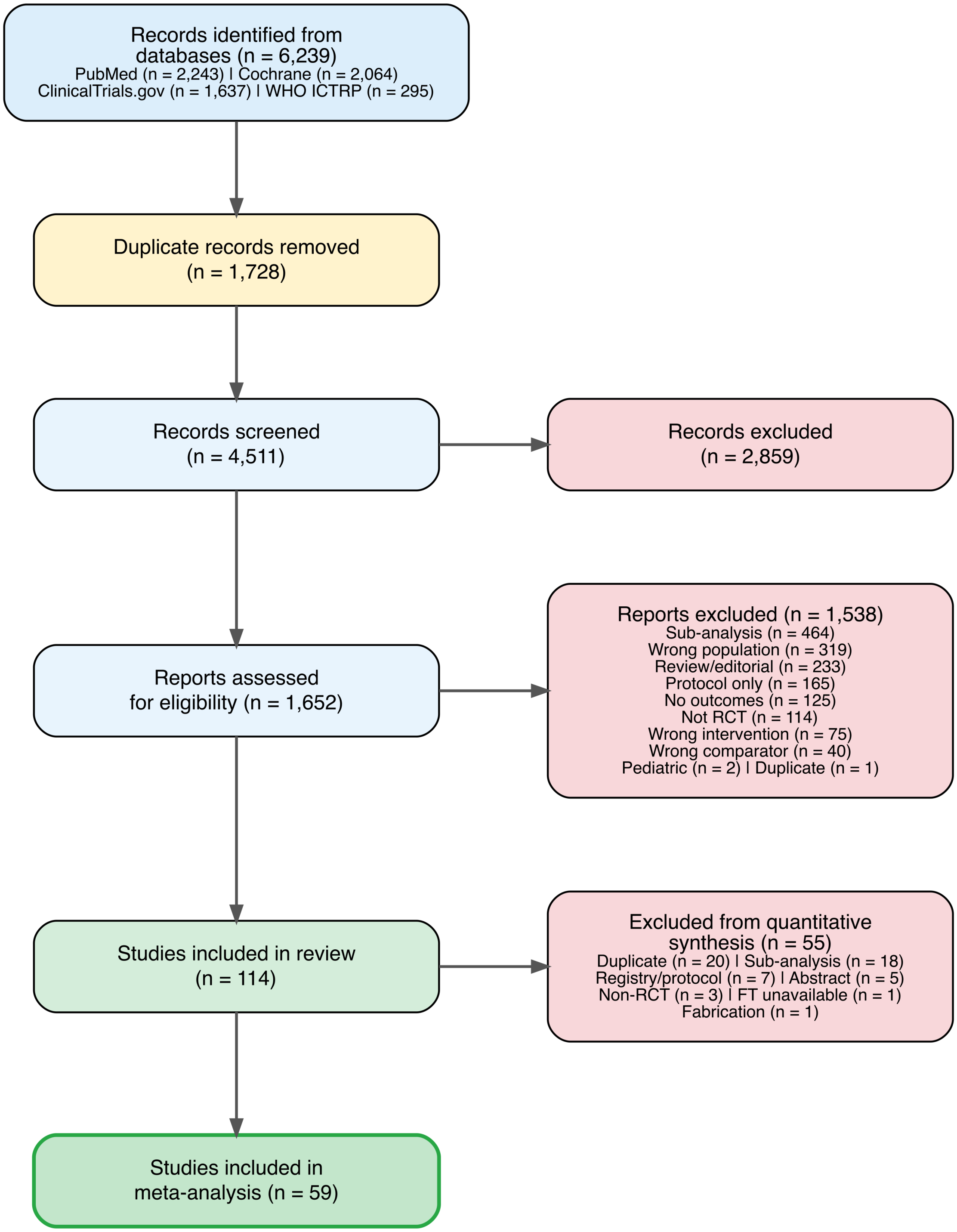


*PRISMA 2020 flow diagram.*

### eFigure 2: Risk of Bias Summary

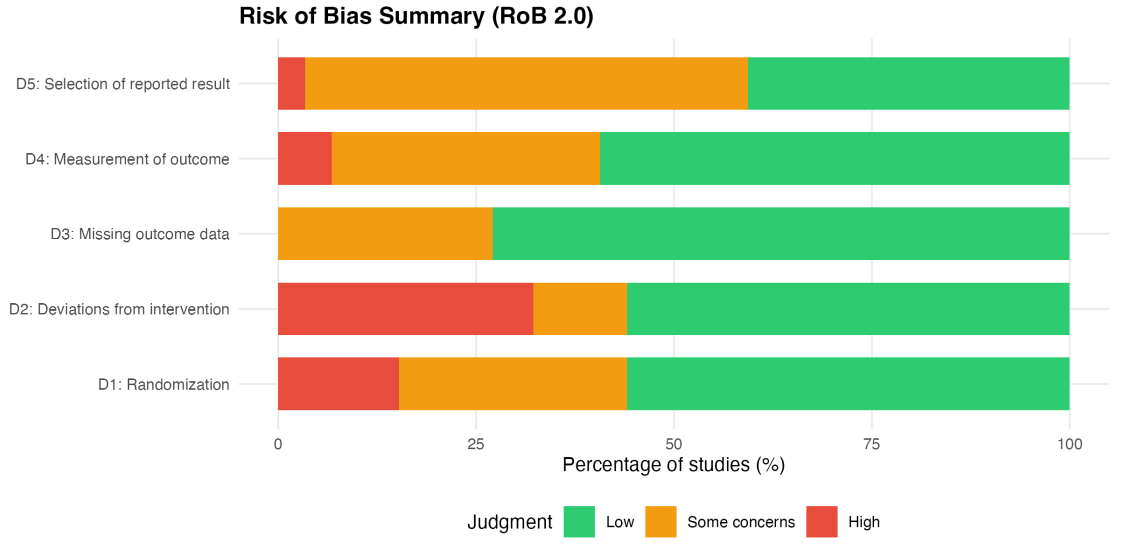


*Risk of Bias 2.0 summary across domains.*

### Primary Outcome Forest Plots

#### eFigure 3: All-Cause Mortality (RR)

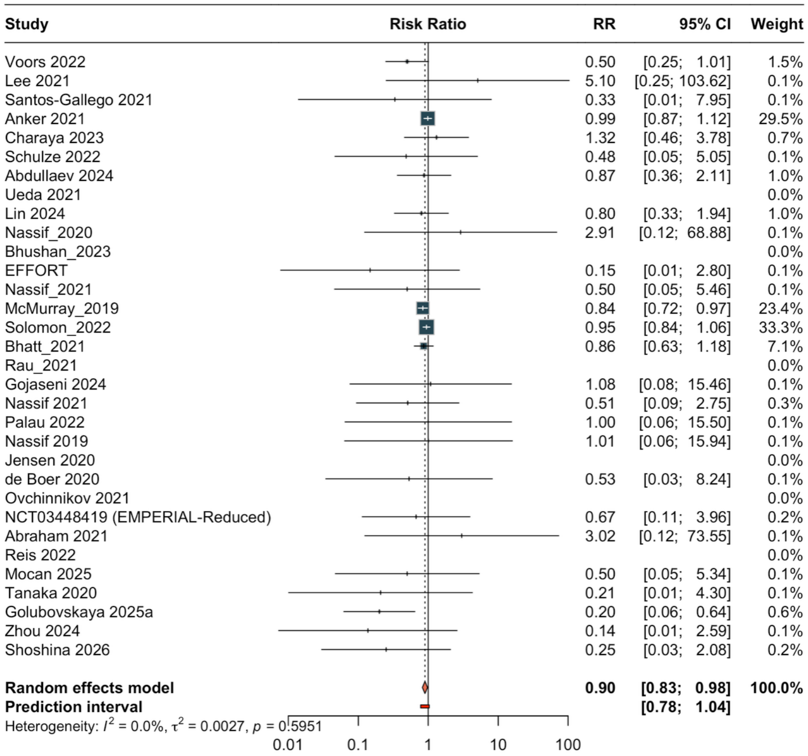

### eFigure 4: All-Cause Mortality (HR)

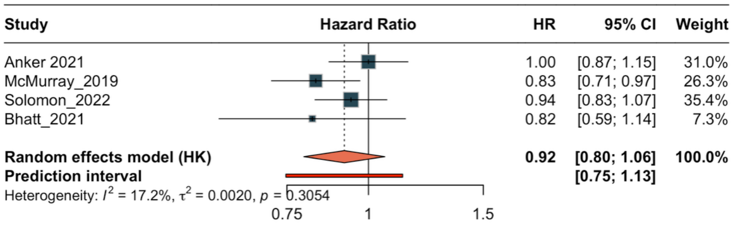

### eFigure 5: HF Hospitalization (RR)

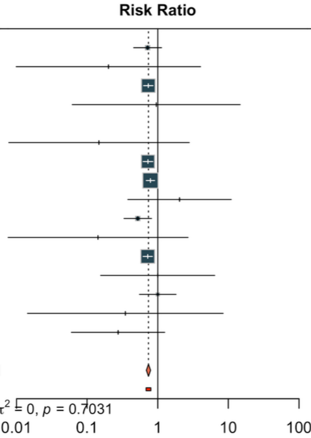


| Study | RR | 95% CI | Weight |
|---|---|---|---|
| Voors 2022 | 0.72 | [0.46; 1.13] | 2.5% |
| Santos-Gallego 2021 | 0.20 | [0.01; 4.04] | 0.1% |
| Anker 2021 | 0.73 | [0.63; 0.86] | 22.0% |
| Ueda 2021 | 0.95 | [0.06; 14.72] | 0.1% |
| Bhushan_2023 | | | 0.0% |
| EFFORT | 0.15 | [0.01; 2.80] | 0.1% |
| McMurray_2019 | 0.73 | [0.62; 0.85] | 20.6% |
| Solomon_2022 | 0.79 | [0.69; 0.90] | 27.7% |
| Nassif 2021 | 2.04 | [0.38; 11.00] | 0.2% |
| Asif 2024 | 0.52 | [0.33; 0.82] | 2.5% |
| Palau 2022 | 0.14 | [0.01; 2.69] | 0.1% |
| Packer 2021 | 0.72 | [0.62; 0.84] | 22.4% |
| Reis 2022 | 1.00 | [0.16; 6.42] | 0.1% |
| Mocan 2025 | 1.00 | [0.55; 1.82] | 1.4% |
| Tanaka 2020 | 0.35 | [0.01; 8.44] | 0.1% |
| Zhou 2024 | 0.27 | [0.06; 1.26] | 0.2% |
| **Random effects model** | **0.74** | **[0.69; 0.79]** | **100.0%** |
| **Prediction interval** | | **[0.68; 0.80]** | |

Heterogeneity: $I^2 = 0.0\%$, $\tau^2 = 0$, $p = 0.7031$

### eFigure 6: HF Hospitalization (HR)

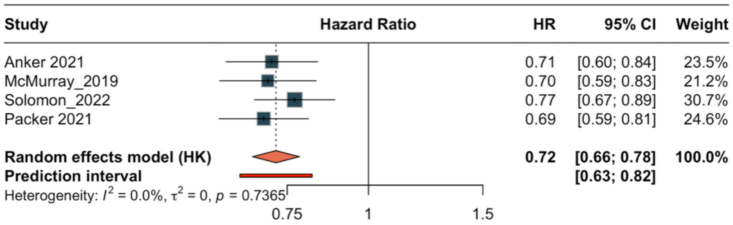

### Secondary Outcome Forest Plots

#### eFigure 7: Cardiovascular Death (RR)

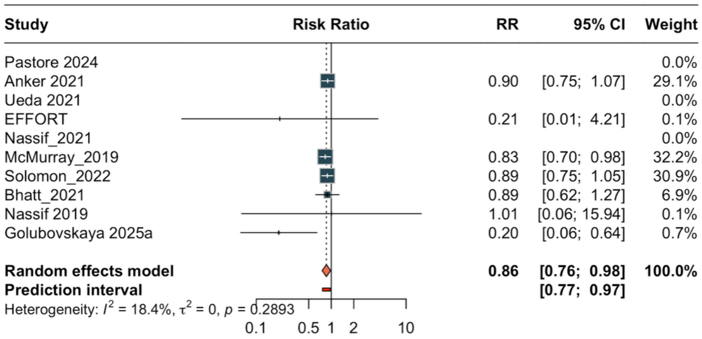

### eFigure 8: Cardiovascular Death (HR)

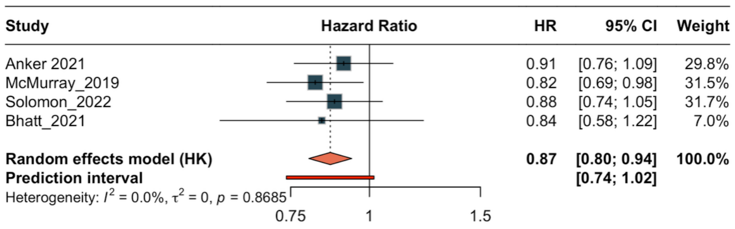

### eFigure 9: CV Death + HF Hospitalization Composite (RR)

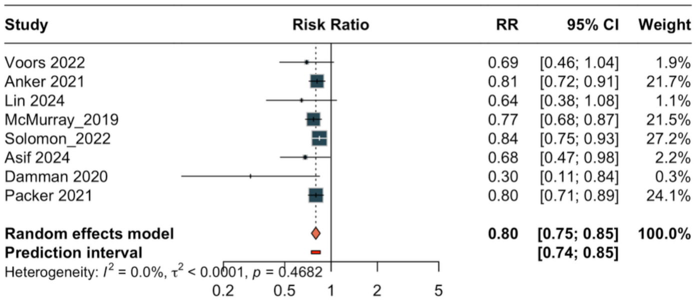

### eFigure 10: Composite (HR)

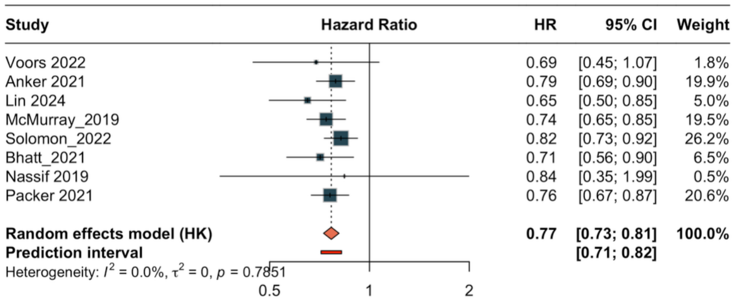

### eFigure 11: Serious Adverse Events

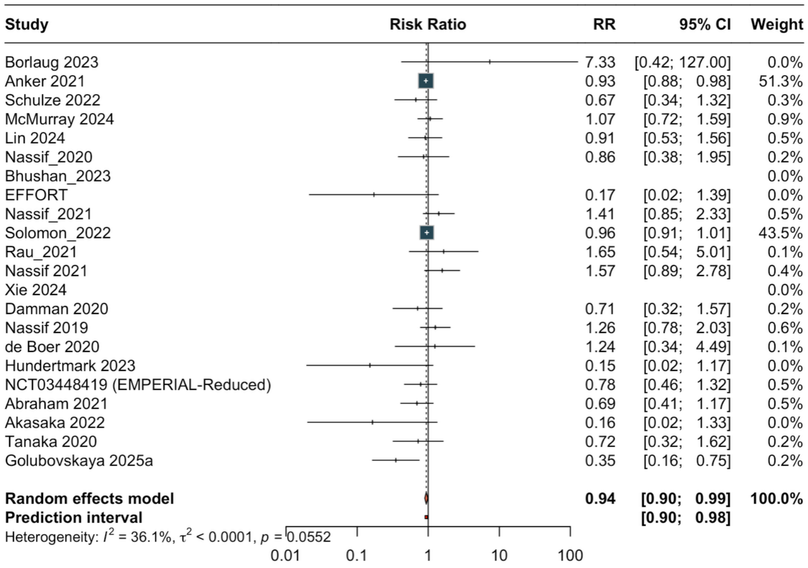

### eFigure 12: KCCQ Total Symptom Score

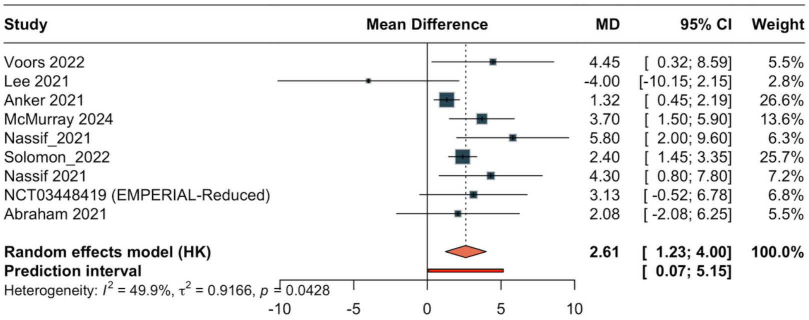

### eFigure 13: KCCQ Overall Summary Score

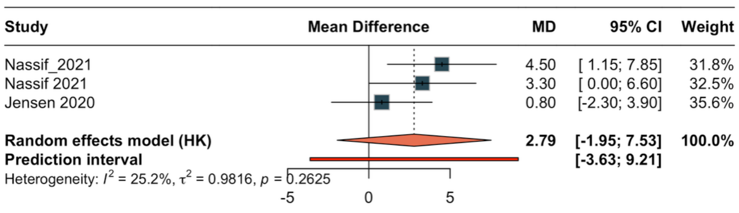

### eFigure 14: 6-Minute Walk Distance

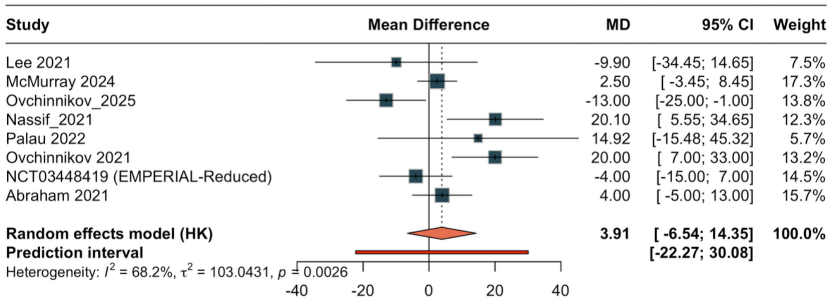

### eFigure 15: NT-proBNP Change

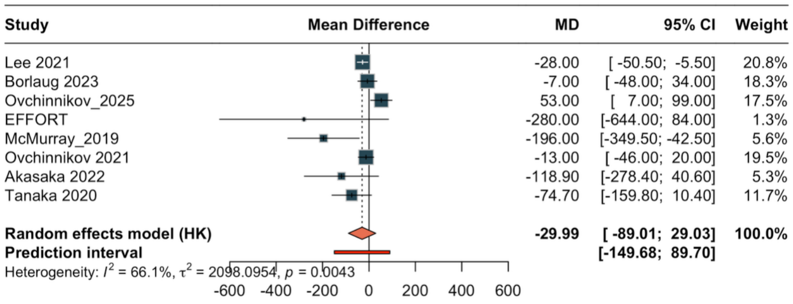

### eFigure 16: eGFR Change

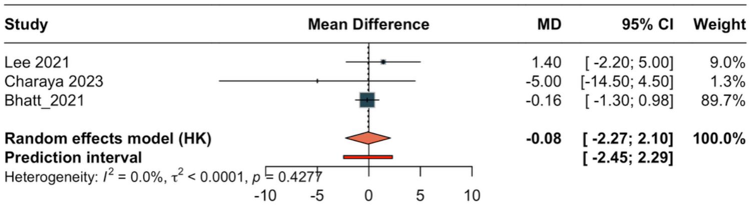

### eFigure 17: Weight Change

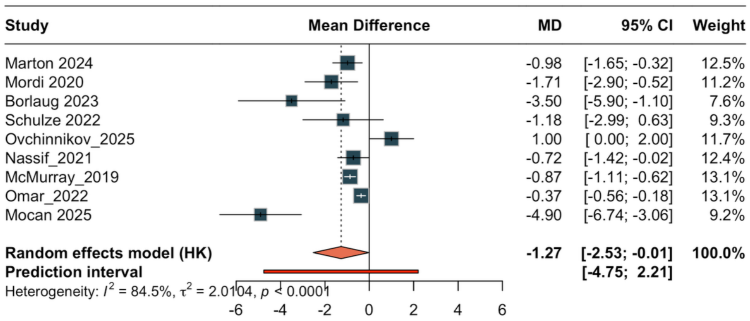

### eFigure 18: Systolic Blood Pressure Change

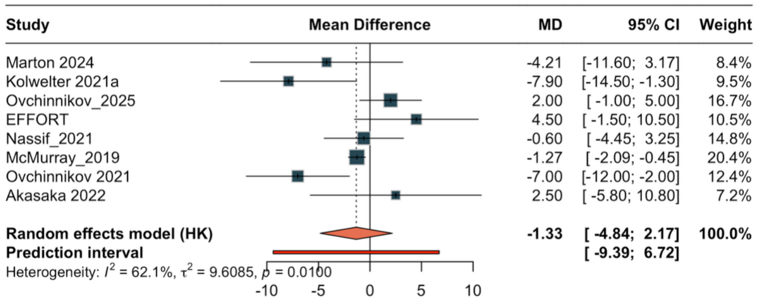

### Safety Forest Plots

#### eFigure 19: Diabetic Ketoacidosis

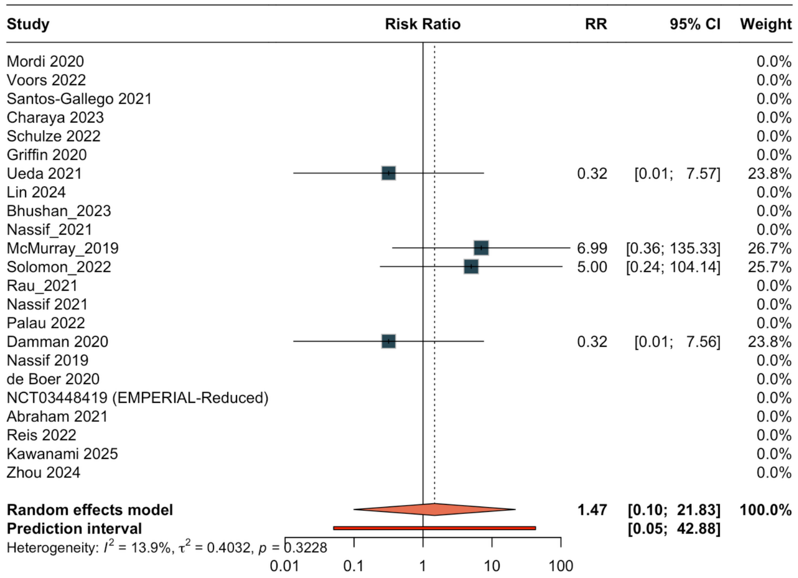

### eFigure 20: Acute Kidney Injury

| Study | RR | 95% CI | Weight |
|---|---|---|---|
| Mordi 2020 | 5.00 | [0.25; 98.64] | 1.3% |
| Charaya 2023 | 1.35 | [0.61; 2.99] | 11.4% |
| Schulze 2022 | 0.32 | [0.10; 1.07] | 6.5% |
| Nassif_2020 | 0.48 | [0.05; 5.09] | 2.0% |
| Nassif_2021 | 1.00 | [0.30; 3.39] | 6.3% |
| McMurray_2019 | 0.58 | [0.39; 0.87] | 20.5% |
| Rau_2021 | | | 0.0% |
| Gojaseni 2024 | 0.72 | [0.27; 1.95] | 8.6% |
| Damman 2020 | 1.27 | [0.30; 5.31] | 4.9% |
| Nassif 2019 | 1.01 | [0.06; 15.94] | 1.5% |
| Fatima Gilani 2023 | 0.76 | [0.43; 1.35] | 15.9% |
| NCT03448419 (EMPERIAL-Reduced) | 1.25 | [0.34; 4.57] | 5.8% |
| Abraham 2021 | 0.60 | [0.15; 2.48] | 5.0% |
| Mocan 2025 | 2.50 | [1.06; 5.92] | 10.3% |
| **Random effects model** | **0.89** | **[0.63; 1.26]** | **100.0%** |
| **Prediction interval** | | **[0.39; 2.04]** | |

Heterogeneity: $I^2 = 24.4\%$, $\tau^2 = 0.1140$, $p = 0.1970$

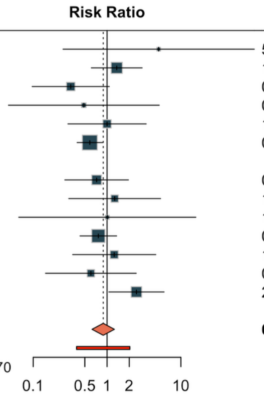

### eFigure 21: Urinary Tract Infection

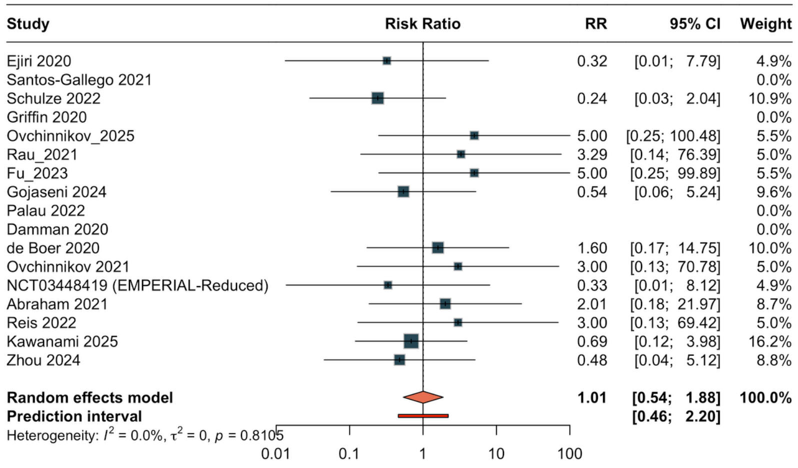

### eFigure 22: Hypotension

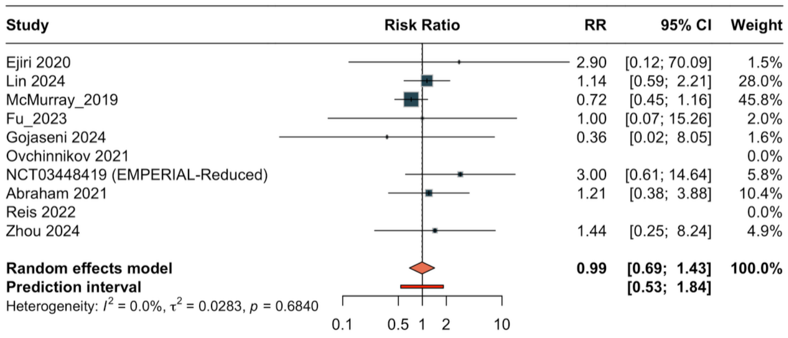

### eFigure 23: Genital Infection

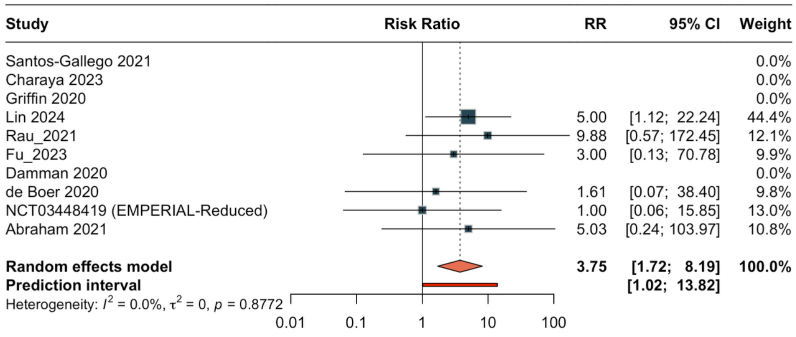

### Subgroup Analyses — All-Cause Mortality

#### eFigure 24: ACM by HF Type

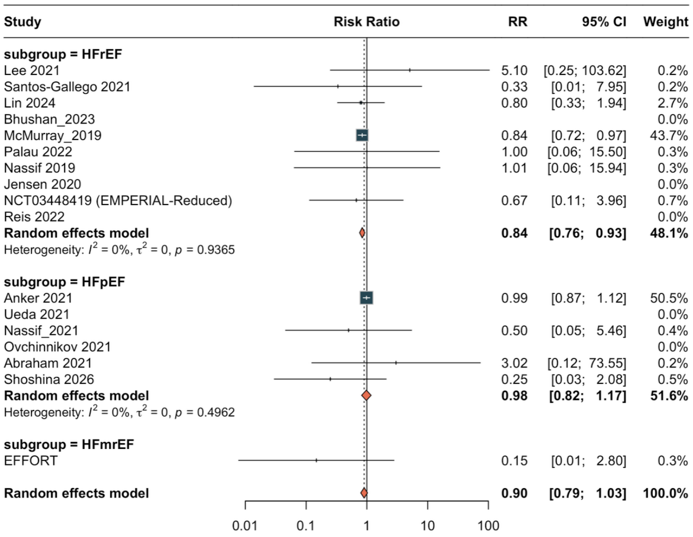

eFigure 25: ACM by Drug Class

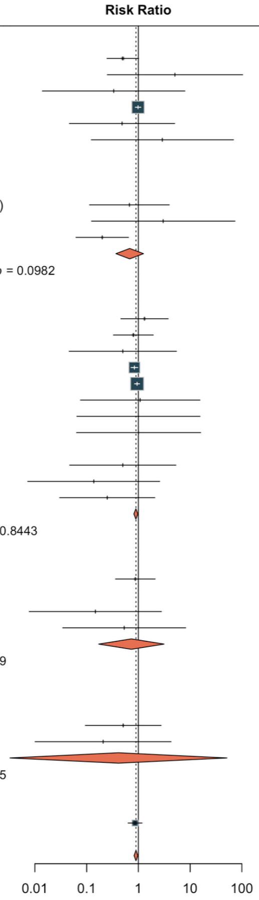

| Study | RR | 95% CI | Weight |
|---|---|---|---|
| **subgroup = Empagliflozin** | | | |
| Voors 2022 | 0.50 | [0.25; 1.01] | 1.5% |
| Lee 2021 | 5.10 | [0.25; 103.62] | 0.1% |
| Santos-Gallego 2021 | 0.33 | [0.01; 7.95] | 0.1% |
| Anker 2021 | 0.99 | [0.87; 1.12] | 29.5% |
| Schulze 2022 | 0.48 | [0.05; 5.05] | 0.1% |
| Nassif_2020 | 2.91 | [0.12; 68.88] | 0.1% |
| Rau_2021 | | | 0.0% |
| Jensen 2020 | | | 0.0% |
| Ovchinnikov 2021 | | | 0.0% |
| NCT03448419 (EMPERIAL-Reduced) | 0.67 | [0.11; 3.96] | 0.2% |
| Abraham 2021 | 3.02 | [0.12; 73.55] | 0.1% |
| Golubovskaya 2025a | 0.20 | [0.06; 0.64] | 0.6% |
| **Random effects model** | **0.68** | **[0.37; 1.25]** | **32.3%** |
| Heterogeneity: $I^2 = 40.4\%$, $\tau^2 = 0.2178$, $p = 0.0982$ | | | |
| **subgroup = Dapagliflozin** | | | |
| Charaya 2023 | 1.32 | [0.46; 3.78] | 0.7% |
| Lin 2024 | 0.80 | [0.33; 1.94] | 1.0% |
| Nassif_2021 | 0.50 | [0.05; 5.46] | 0.1% |
| McMurray_2019 | 0.84 | [0.72; 0.97] | 23.4% |
| Solomon_2022 | 0.95 | [0.84; 1.06] | 33.3% |
| Gojaseni 2024 | 1.08 | [0.08; 15.46] | 0.1% |
| Palau 2022 | 1.00 | [0.06; 15.50] | 0.1% |
| Nassif 2019 | 1.01 | [0.06; 15.94] | 0.1% |
| Reis 2022 | | | 0.0% |
| Mocan 2025 | 0.50 | [0.05; 5.34] | 0.1% |
| Zhou 2024 | 0.14 | [0.01; 2.59] | 0.1% |
| Shoshina 2026 | 0.25 | [0.03; 2.08] | 0.2% |
| **Random effects model** | **0.89** | **[0.82; 0.98]** | **59.1%** |
| Heterogeneity: $I^2 = 0\%$, $\tau^2 = 0.0023$, $p = 0.8443$ | | | |
| **subgroup = Other** | | | |
| Abdullaev 2024 | 0.87 | [0.36; 2.11] | 1.0% |
| Bhushan_2023 | | | 0.0% |
| EFFORT | 0.15 | [0.01; 2.80] | 0.1% |
| de Boer 2020 | 0.53 | [0.03; 8.24] | 0.1% |
| **Random effects model** | **0.73** | **[0.17; 3.14]** | **1.2%** |
| Heterogeneity: $I^2 = 0\%$, $\tau^2 = 0$, $p = 0.5099$ | | | |
| **subgroup = Canagliflozin** | | | |
| Ueda 2021 | | | 0.0% |
| Nassif 2021 | 0.51 | [0.09; 2.75] | 0.3% |
| Tanaka 2020 | 0.21 | [0.01; 4.30] | 0.1% |
| **Random effects model** | **0.41** | **[0.00; 51.33]** | **0.4%** |
| Heterogeneity: $I^2 = 0\%$, $\tau^2 = 0$, $p = 0.6135$ | | | |
| **subgroup = Sotagliflozin** | | | |
| Bhatt_2021 | 0.86 | [0.63; 1.18] | 7.1% |
| **Random effects model** | **0.90** | **[0.83; 0.98]** | **100.0%** |

#### eFigure 26: ACM by Blinding

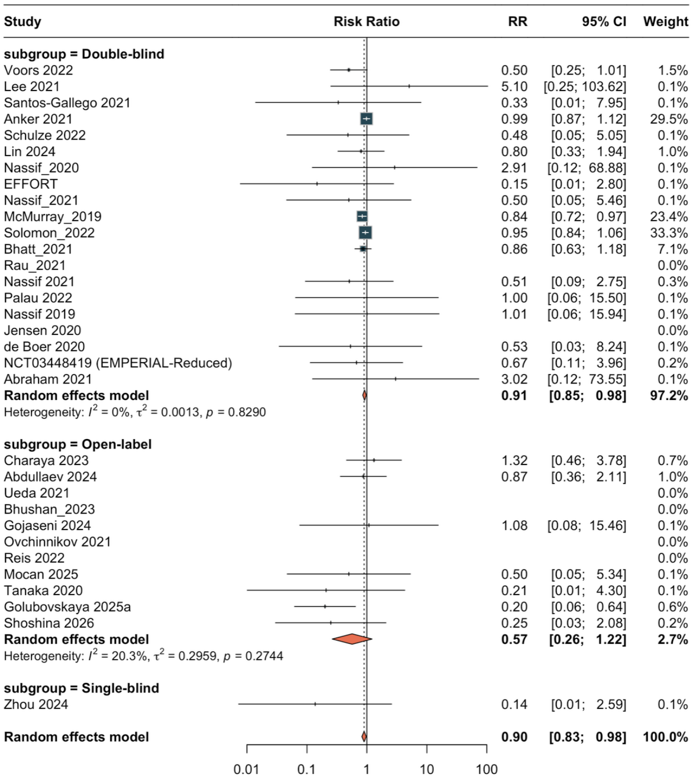
Study
Risk Ratio
RR
95% CI
Weight
subgroup = Double-blind
Voors 2022 0.50 [0.25; 1.01] 1.5%
Lee 2021 5.10 [0.25; 103.62] 0.1%
Santos-Gallego 2021 0.33 [0.01; 7.95] 0.1%
Anker 2021 0.99 [0.87; 1.12] 29.5%
Schulze 2022 0.48 [0.05; 5.05] 0.1%
Lin 2024 0.80 [0.33; 1.94] 1.0%
Nassif_2020 2.91 [0.12; 68.88] 0.1%
EFFORT 0.15 [0.01; 2.80] 0.1%
Nassif_2021 0.50 [0.05; 5.46] 0.1%
McMurray_2019 0.84 [0.72; 0.97] 23.4%
Solomon_2022 0.95 [0.84; 1.06] 33.3%
Bhatt_2021 0.86 [0.63; 1.18] 7.1%
Rau_2021 0.0%
Nassif 2021 0.51 [0.09; 2.75] 0.3%
Palau 2022 1.00 [0.06; 15.50] 0.1%
Nassif 2019 1.01 [0.06; 15.94] 0.1%
Jensen 2020 0.0%
de Boer 2020 0.53 [0.03; 8.24] 0.1%
NCT03448419 (EMPERIAL-Reduced) 0.67 [0.11; 3.96] 0.2%
Abraham 2021 3.02 [0.12; 73.55] 0.1%
Random effects model 0.91 [0.85; 0.98] 97.2%
Heterogeneity: $I^2$ = 0%, $\tau^2$ = 0.0013, $p$ = 0.8290
subgroup = Open-label
Charaya 2023 1.32 [0.46; 3.78] 0.7%
Abdullaev 2024 0.87 [0.36; 2.11] 1.0%
Ueda 2021 0.0%
Bhushan_2023 0.0%
Gojaseni 2024 1.08 [0.08; 15.46] 0.1%
Ovchinnikov 2021 0.0%
Reis 2022 0.0%
Mocan 2025 0.50 [0.05; 5.34] 0.1%
Tanaka 2020 0.21 [0.01; 4.30] 0.1%
Golubovskaya 2025a 0.20 [0.06; 0.64] 0.6%
Shoshina 2026 0.25 [0.03; 2.08] 0.2%
Random effects model 0.57 [0.26; 1.22] 2.7%
Heterogeneity: $I^2$ = 20.3%, $\tau^2$ = 0.2959, $p$ = 0.2744
subgroup = Single-blind
Zhou 2024 0.14 [0.01; 2.59] 0.1%
Random effects model 0.90 [0.83; 0.98] 100.0%
0.01
0.1
1
10
100

#### eFigure 27: ACM by Risk of Bias

| Study | RR | 95% CI | Weight |
|---|---|---|---|
| **subgroup = Low** | | | |
| Voors 2022 | 0.50 | [0.25; 1.01] | 1.5% |
| Lee 2021 | 5.10 | [0.25; 103.62] | 0.1% |
| Santos-Gallego 2021 | 0.33 | [0.01; 7.95] | 0.1% |
| Anker 2021 | 0.99 | [0.87; 1.12] | 29.5% |
| Schulze 2022 | 0.48 | [0.05; 5.05] | 0.1% |
| Nassif_2020 | 2.91 | [0.12; 68.88] | 0.1% |
| EFFORT | 0.15 | [0.01; 2.80] | 0.1% |
| Nassif_2021 | 0.50 | [0.05; 5.46] | 0.1% |
| McMurray_2019 | 0.84 | [0.72; 0.97] | 23.4% |
| Solomon_2022 | 0.95 | [0.84; 1.06] | 33.3% |
| Palau 2022 | 1.00 | [0.06; 15.50] | 0.1% |
| NCT03448419 (EMPERIAL-Reduced) | 0.67 | [0.11; 3.96] | 0.2% |
| Abraham 2021 | 3.02 | [0.12; 73.55] | 0.1% |
| **Random effects model** | **0.92** | **[0.84; 1.00]** | **88.6%** |
| Heterogeneity: $I^2 = 0\%$, $\tau^2 = 0.0018$, $p = 0.5633$ | | | |
| **subgroup = High** | | | |
| Charaya 2023 | 1.32 | [0.46; 3.78] | 0.7% |
| Abdullaev 2024 | 0.87 | [0.36; 2.11] | 1.0% |
| Ueda 2021 | | | 0.0% |
| Bhushan_2023 | | | 0.0% |
| Gojaseni 2024 | 1.08 | [0.08; 15.46] | 0.1% |
| Ovchinnikov 2021 | | | 0.0% |
| Reis 2022 | | | 0.0% |
| Mocan 2025 | 0.50 | [0.05; 5.34] | 0.1% |
| Golubovskaya 2025a | 0.20 | [0.06; 0.64] | 0.6% |
| Shoshina 2026 | 0.25 | [0.03; 2.08] | 0.2% |
| **Random effects model** | **0.60** | **[0.25; 1.41]** | **2.6%** |
| Heterogeneity: $I^2 = 28.7\%$, $\tau^2 = 0.3131$, $p = 0.2195$ | | | |
| **subgroup = Some concerns** | | | |
| Lin 2024 | 0.80 | [0.33; 1.94] | 1.0% |
| Bhatt_2021 | 0.86 | [0.63; 1.18] | 7.1% |
| Rau_2021 | | | 0.0% |
| Nassif 2021 | 0.51 | [0.09; 2.75] | 0.3% |
| Nassif 2019 | 1.01 | [0.06; 15.94] | 0.1% |
| Jensen 2020 | | | 0.0% |
| de Boer 2020 | 0.53 | [0.03; 8.24] | 0.1% |
| Tanaka 2020 | 0.21 | [0.01; 4.30] | 0.1% |
| Zhou 2024 | 0.14 | [0.01; 2.59] | 0.1% |
| **Random effects model** | **0.82** | **[0.64; 1.04]** | **8.7%** |
| Heterogeneity: $I^2 = 0\%$, $\tau^2 = 0$, $p = 0.8403$ | | | |
| **Random effects model** | **0.90** | **[0.83; 0.98]** | **100.0%** |

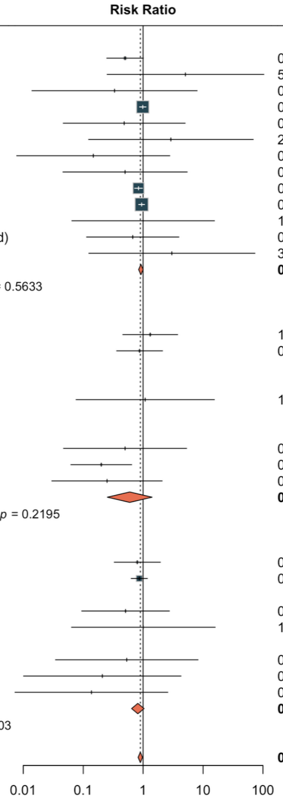

### eFigure 28: ACM by Follow-Up Duration

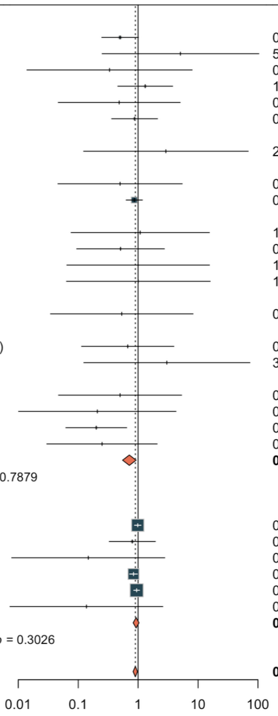


| Study | Risk Ratio | RR | 95% CI | Weight |
|---|---|---|---|---|
| **subgroup = <12 months** | | | | |
| Voors 2022 | | 0.50 | [0.25; 1.01] | 1.5% |
| Lee 2021 | | 5.10 | [0.25; 103.62] | 0.1% |
| Santos-Gallego 2021 | | 0.33 | [0.01; 7.95] | 0.1% |
| Charaya 2023 | | 1.32 | [0.46; 3.78] | 0.7% |
| Schulze 2022 | | 0.48 | [0.05; 5.05] | 0.1% |
| Abdullaev 2024 | | 0.87 | [0.36; 2.11] | 1.0% |
| Ueda 2021 | | | | 0.0% |
| Nassif_2020 | | 2.91 | [0.12; 68.88] | 0.1% |
| Bhushan_2023 | | | | 0.0% |
| Nassif_2021 | | 0.50 | [0.05; 5.46] | 0.1% |
| Bhatt_2021 | | 0.86 | [0.63; 1.18] | 7.1% |
| Rau_2021 | | | | 0.0% |
| Gojaseni 2024 | | 1.08 | [0.08; 15.46] | 0.1% |
| Nassif 2021 | | 0.51 | [0.09; 2.75] | 0.3% |
| Palau 2022 | | 1.00 | [0.06; 15.50] | 0.1% |
| Nassif 2019 | | 1.01 | [0.06; 15.94] | 0.1% |
| Jensen 2020 | | | | 0.0% |
| de Boer 2020 | | 0.53 | [0.03; 8.24] | 0.1% |
| Ovchinnikov 2021 | | | | 0.0% |
| NCT03448419 (EMPERIAL-Reduced) | | 0.67 | [0.11; 3.96] | 0.2% |
| Abraham 2021 | | 3.02 | [0.12; 73.55] | 0.1% |
| Reis 2022 | | | | 0.0% |
| Mocan 2025 | | 0.50 | [0.05; 5.34] | 0.1% |
| Tanaka 2020 | | 0.21 | [0.01; 4.30] | 0.1% |
| Golubovskaya 2025a | | 0.20 | [0.06; 0.64] | 0.6% |
| Shoshina 2026 | | 0.25 | [0.03; 2.08] | 0.2% |
| **Random effects model** | | **0.71** | **[0.55; 0.92]** | **12.8%** |
| Heterogeneity: $I^2 = 0\%$, $\tau^2 = 0.0319$, $p = 0.7879$ | | | | |
| **subgroup = ≥12 months** | | | | |
| Anker 2021 | | 0.99 | [0.87; 1.12] | 29.5% |
| Lin 2024 | | 0.80 | [0.33; 1.94] | 1.0% |
| EFFORT | | 0.15 | [0.01; 2.80] | 0.1% |
| McMurray_2019 | | 0.84 | [0.72; 0.97] | 23.4% |
| Solomon_2022 | | 0.95 | [0.84; 1.06] | 33.3% |
| Zhou 2024 | | 0.14 | [0.01; 2.59] | 0.1% |
| **Random effects model** | | **0.93** | **[0.83; 1.04]** | **87.2%** |
| Heterogeneity: $I^2 = 17.2\%$, $\tau^2 = 0.0011$, $p = 0.3026$ | | | | |
| **Random effects model** | | **0.90** | **[0.83; 0.98]** | **100.0%** |

#### eFigure 29: ACM by Sample Size

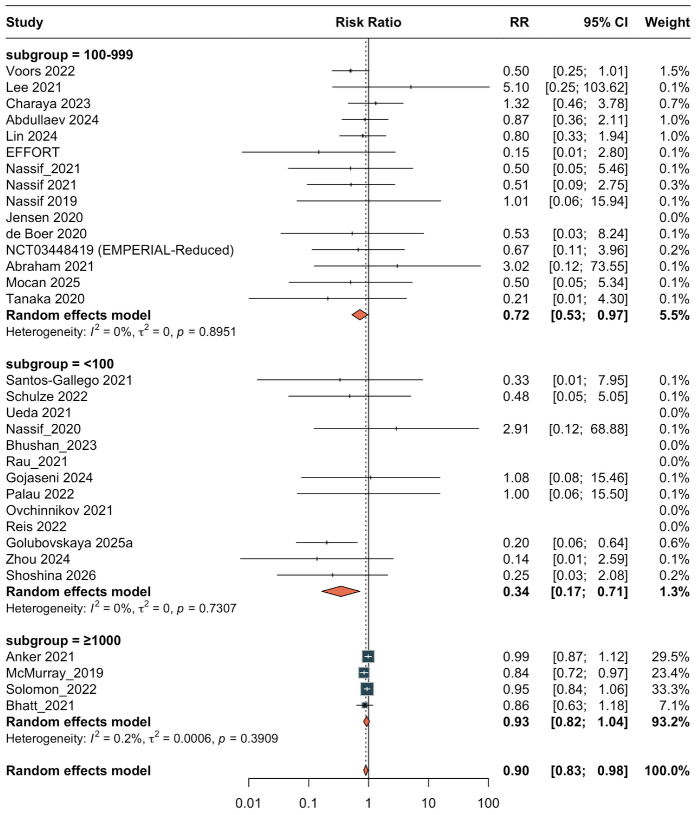
Study
Risk Ratio
RR
95% CI
Weight
subgroup = 100-999
Voors 2022 0.50 [0.25; 1.01] 1.5%
Lee 2021 5.10 [0.25; 103.62] 0.1%
Charaya 2023 1.32 [0.46; 3.78] 0.7%
Abdullaev 2024 0.87 [0.36; 2.11] 1.0%
Lin 2024 0.80 [0.33; 1.94] 1.0%
EFFORT 0.15 [0.01; 2.80] 0.1%
Nassif_2021 0.50 [0.05; 5.46] 0.1%
Nassif 2021 0.51 [0.09; 2.75] 0.3%
Nassif 2019 1.01 [0.06; 15.94] 0.1%
Jensen 2020 0.0%
de Boer 2020 0.53 [0.03; 8.24] 0.1%
NCT03448419 (EMPERIAL-Reduced) 0.67 [0.11; 3.96] 0.2%
Abraham 2021 3.02 [0.12; 73.55] 0.1%
Mocan 2025 0.50 [0.05; 5.34] 0.1%
Tanaka 2020 0.21 [0.01; 4.30] 0.1%
Random effects model 0.72 [0.53; 0.97] 5.5%
Heterogeneity: I2 = 0%, τ2 = 0, p = 0.8951
subgroup = <100
Santos-Gallego 2021 0.33 [0.01; 7.95] 0.1%
Schulze 2022 0.48 [0.05; 5.05] 0.1%
Ueda 2021 0.0%
Nassif_2020 2.91 [0.12; 68.88] 0.1%
Bhushan_2023 0.0%
Rau_2021 0.0%
Gojaseni 2024 1.08 [0.08; 15.46] 0.1%
Palau 2022 1.00 [0.06; 15.50] 0.1%
Ovchinnikov 2021 0.0%
Reis 2022 0.0%
Golubovskaya 2025a 0.20 [0.06; 0.64] 0.6%
Zhou 2024 0.14 [0.01; 2.59] 0.1%
Shoshina 2026 0.25 [0.03; 2.08] 0.2%
Random effects model 0.34 [0.17; 0.71] 1.3%
Heterogeneity: I2 = 0%, τ2 = 0, p = 0.7307
subgroup = ≥1000
Anker 2021 0.99 [0.87; 1.12] 29.5%
McMurray_2019 0.84 [0.72; 0.97] 23.4%
Solomon_2022 0.95 [0.84; 1.06] 33.3%
Bhatt_2021 0.86 [0.63; 1.18] 7.1%
Random effects model 0.93 [0.82; 1.04] 93.2%
Heterogeneity: I2 = 0.2%, τ2 = 0.0006, p = 0.3909
Random effects model 0.90 [0.83; 0.98] 100.0%
0.01
0.1
1
10
100

#### eFigure 30: ACM by Publication Era

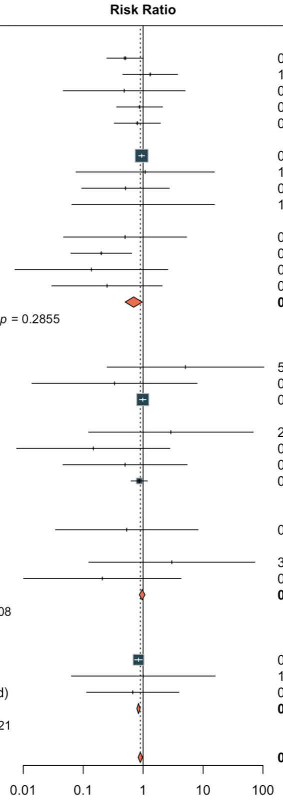


| Study | RR | 95% CI | Weight |
|---|---|---|---|
| **subgroup = 2022+** | | | |
| Voors 2022 | 0.50 | [0.25; 1.01] | 1.5% |
| Charaya 2023 | 1.32 | [0.46; 3.78] | 0.7% |
| Schulze 2022 | 0.48 | [0.05; 5.05] | 0.1% |
| Abdullaev 2024 | 0.87 | [0.36; 2.11] | 1.0% |
| Lin 2024 | 0.80 | [0.33; 1.94] | 1.0% |
| Bhushan_2023 | | | 0.0% |
| Solomon_2022 | 0.95 | [0.84; 1.06] | 33.3% |
| Gojaseni 2024 | 1.08 | [0.08; 15.46] | 0.1% |
| Nassif 2021 | 0.51 | [0.09; 2.75] | 0.3% |
| Palau 2022 | 1.00 | [0.06; 15.50] | 0.1% |
| Reis 2022 | | | 0.0% |
| Mocan 2025 | 0.50 | [0.05; 5.34] | 0.1% |
| Golubovskaya 2025a | 0.20 | [0.06; 0.64] | 0.6% |
| Zhou 2024 | 0.14 | [0.01; 2.59] | 0.1% |
| Shoshina 2026 | 0.25 | [0.03; 2.08] | 0.2% |
| **Random effects model** | **0.70** | **[0.50; 0.97]** | **39.0%** |
| Heterogeneity: $I^2$ = 15.7%, $\tau^2$ = 0.0887, $p$ = 0.2855 | | | |
| **subgroup = 2020-2021** | | | |
| Lee 2021 | 5.10 | [0.25; 103.62] | 0.1% |
| Santos-Gallego 2021 | 0.33 | [0.01; 7.95] | 0.1% |
| Anker 2021 | 0.99 | [0.87; 1.12] | 29.5% |
| Ueda 2021 | | | 0.0% |
| Nassif_2020 | 2.91 | [0.12; 68.88] | 0.1% |
| EFFORT | 0.15 | [0.01; 2.80] | 0.1% |
| Nassif_2021 | 0.50 | [0.05; 5.46] | 0.1% |
| Bhatt_2021 | 0.86 | [0.63; 1.18] | 7.1% |
| Rau_2021 | | | 0.0% |
| Jensen 2020 | | | 0.0% |
| de Boer 2020 | 0.53 | [0.03; 8.24] | 0.1% |
| Ovchinnikov 2021 | | | 0.0% |
| Abraham 2021 | 3.02 | [0.12; 73.55] | 0.1% |
| Tanaka 2020 | 0.21 | [0.01; 4.30] | 0.1% |
| **Random effects model** | **0.96** | **[0.86; 1.08]** | **37.3%** |
| Heterogeneity: $I^2$ = 0%, $\tau^2$ = 0, $p$ = 0.7208 | | | |
| **subgroup = Pre-2020** | | | |
| McMurray_2019 | 0.84 | [0.72; 0.97] | 23.4% |
| Nassif 2019 | 1.01 | [0.06; 15.94] | 0.1% |
| NCT03448419 (EMPERIAL-Reduced) | 0.67 | [0.11; 3.96] | 0.2% |
| **Random effects model** | **0.84** | **[0.79; 0.89]** | **23.7%** |
| Heterogeneity: $I^2$ = 0%, $\tau^2$ = 0, $p$ = 0.9621 | | | |
| **Random effects model** | **0.90** | **[0.83; 0.98]** | **100.0%** |

### Subgroup Analyses — HF Hospitalization

#### eFigure 31: HFH by HF Type

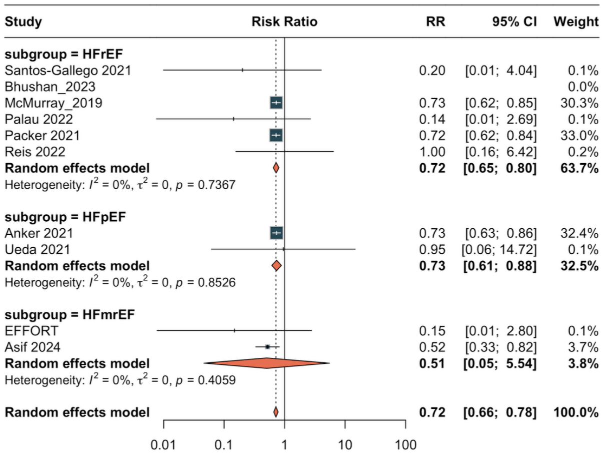

### eFigure 32: HFH by Drug Class

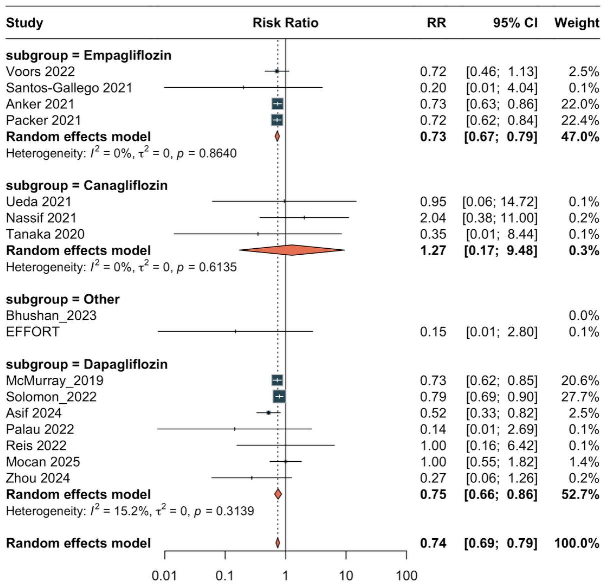

### eFigure 33: HFH by Blinding

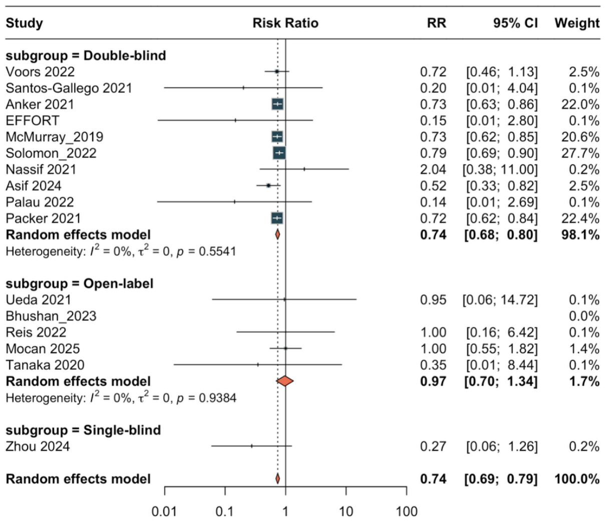

### eFigure 34: HFH by Risk of Bias

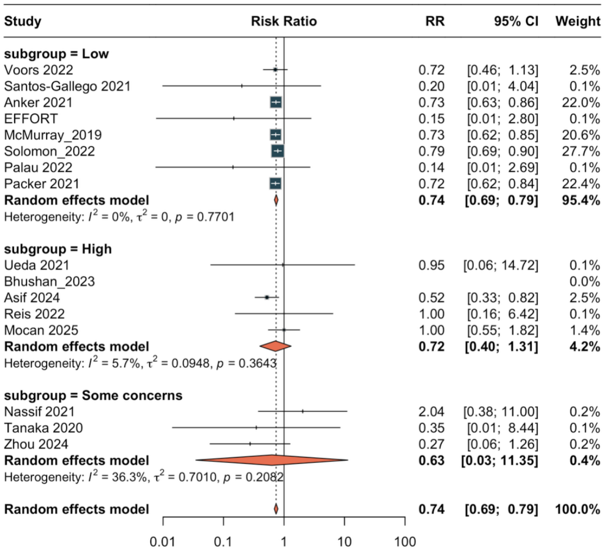

### eFigure 35: HFH by Follow-Up Duration

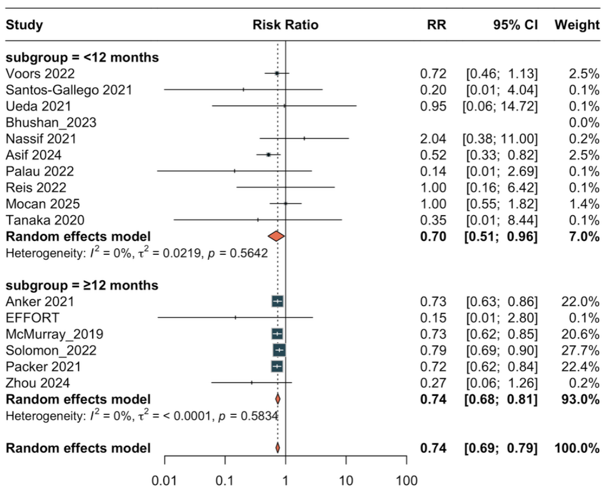

### eFigure 36: HFH by Sample Size

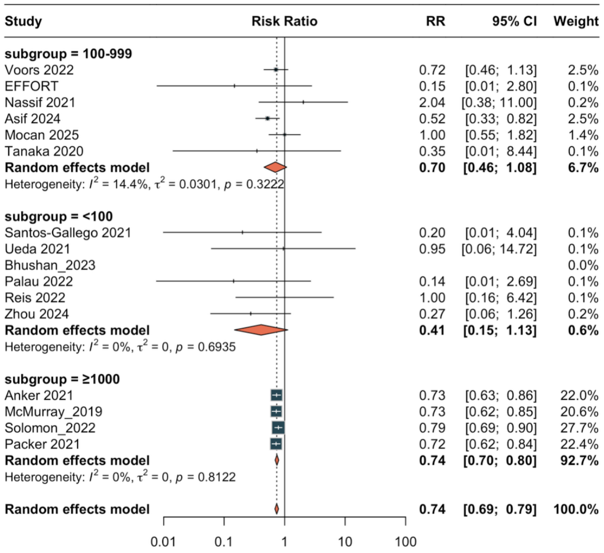

### eFigure 37: HFH by Publication Era

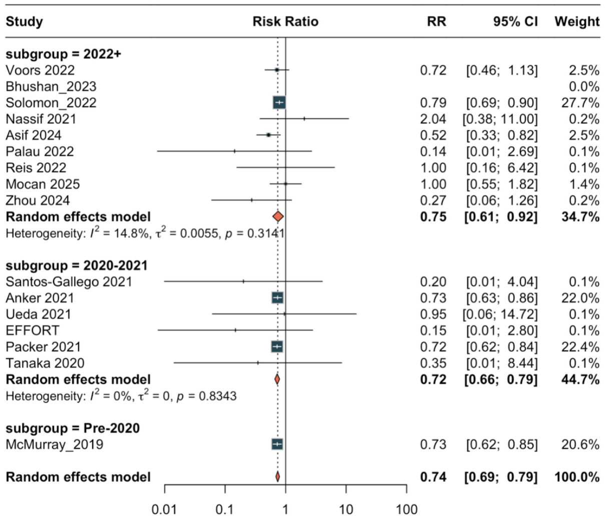

### Publication Bias

#### eFigure 38: Funnel Plot — ACM

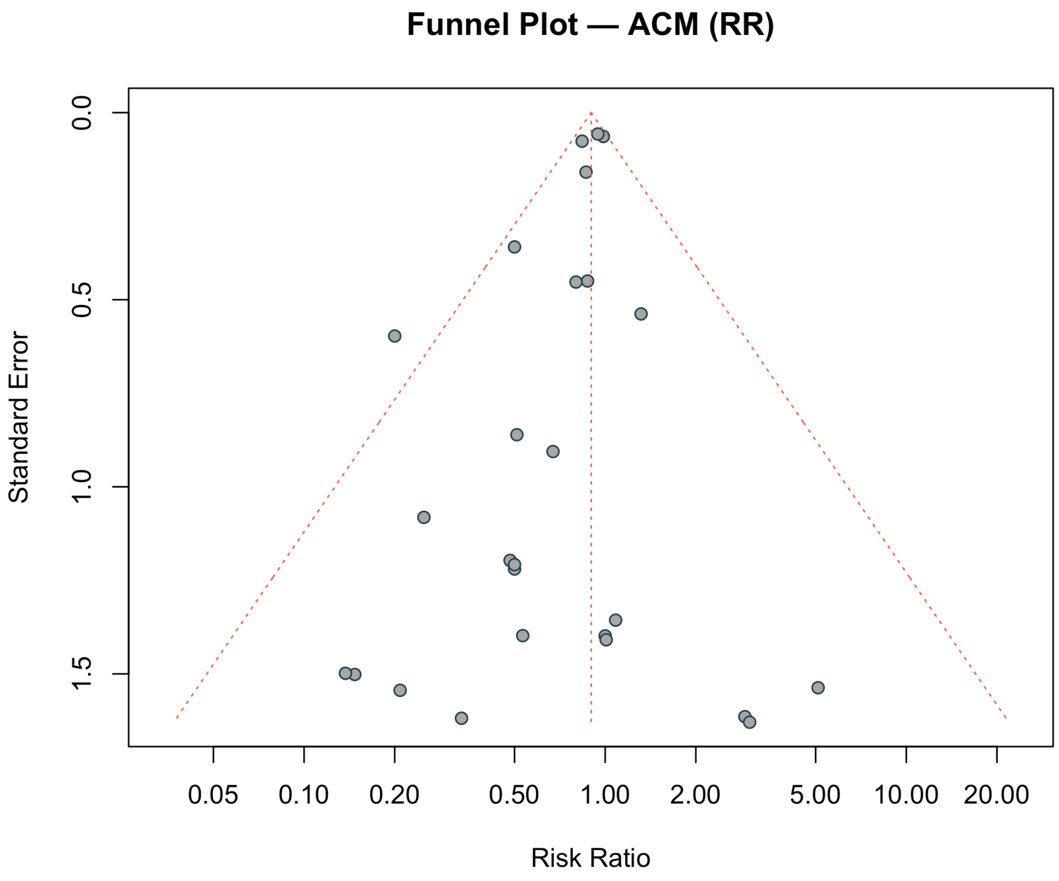

#### eFigure 39: Funnel Plot — HFH

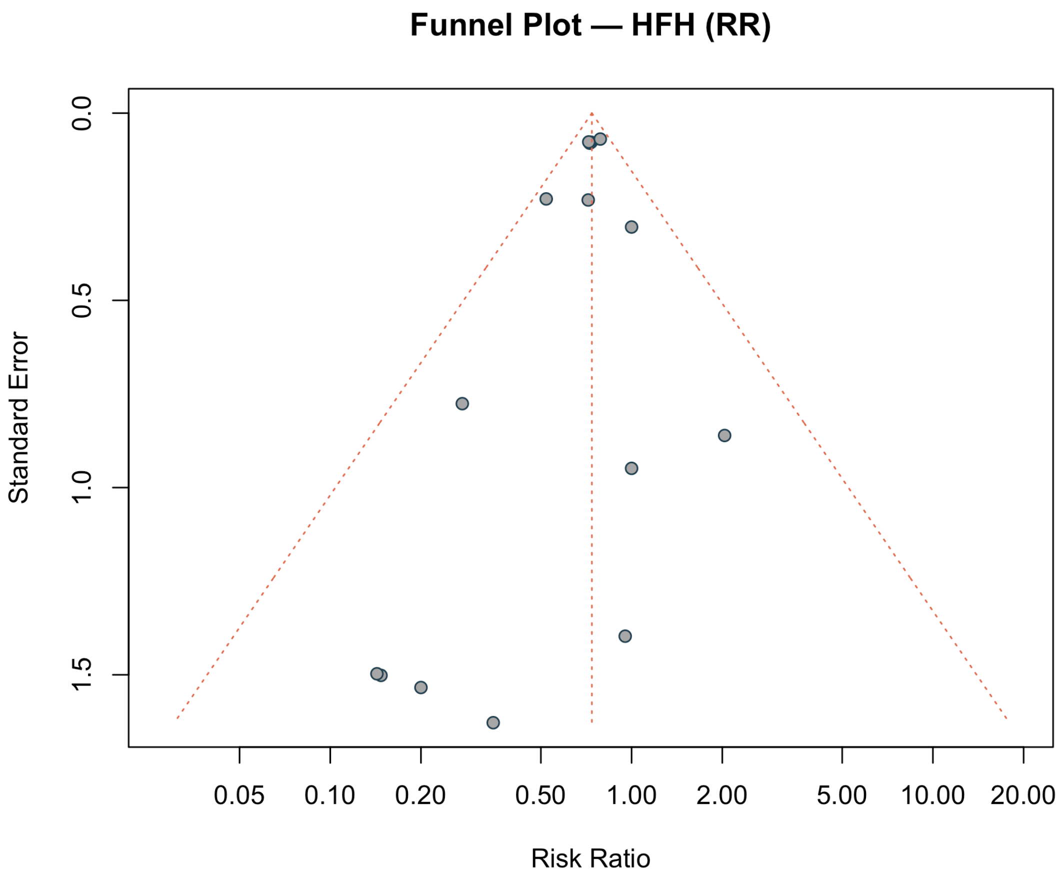

#### eFigure 40: Funnel Plot — CVD

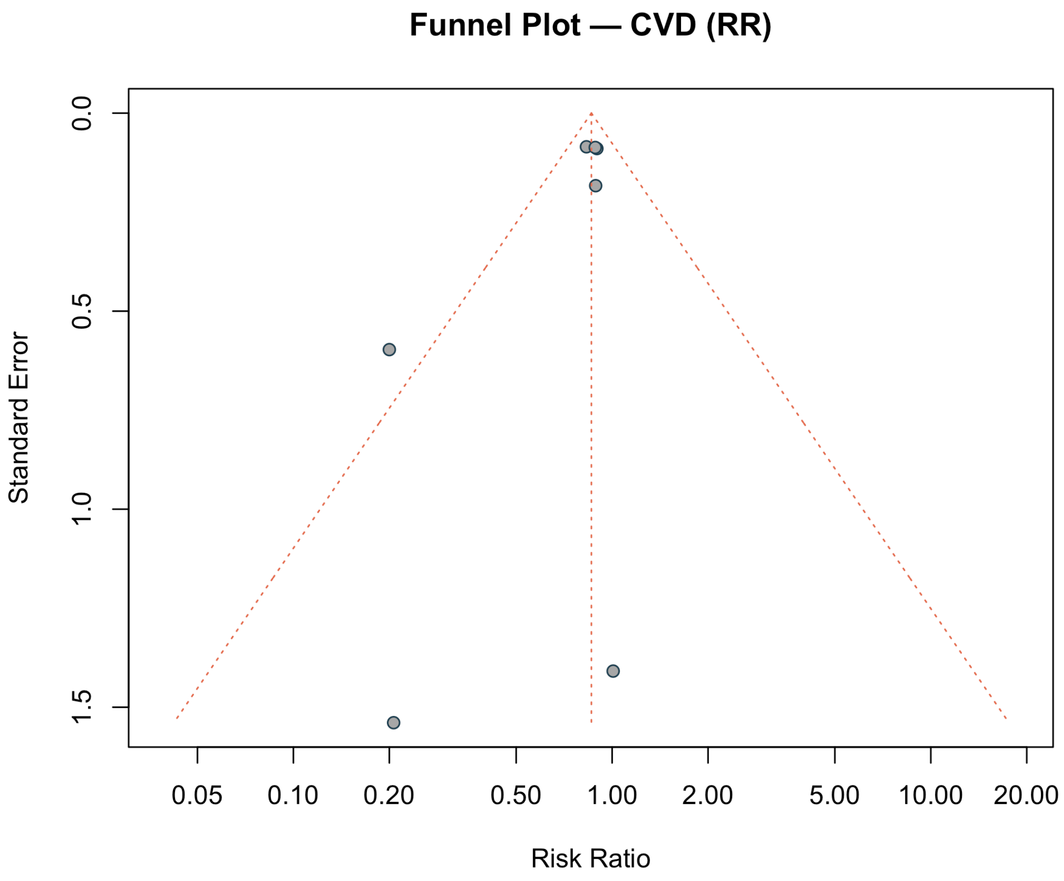

#### eFigure 41: Funnel Plot — Composite

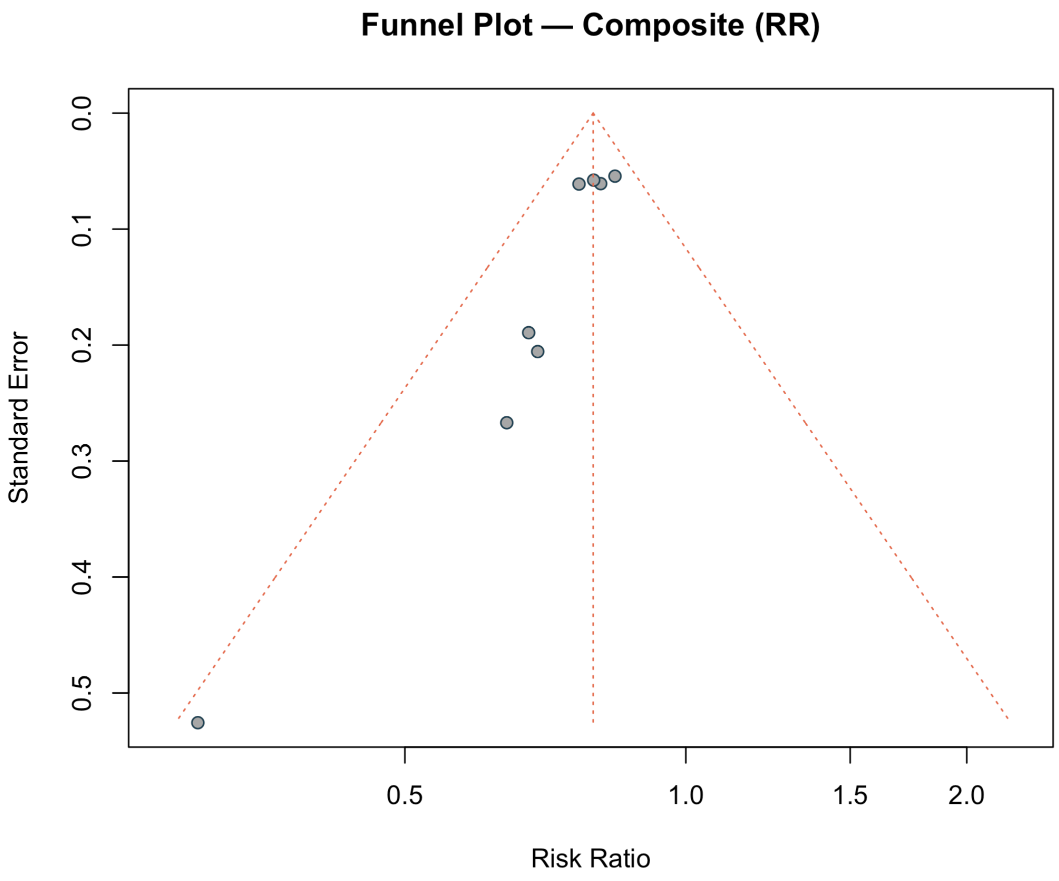

#### eFigure 42: Funnel Plot — SAE

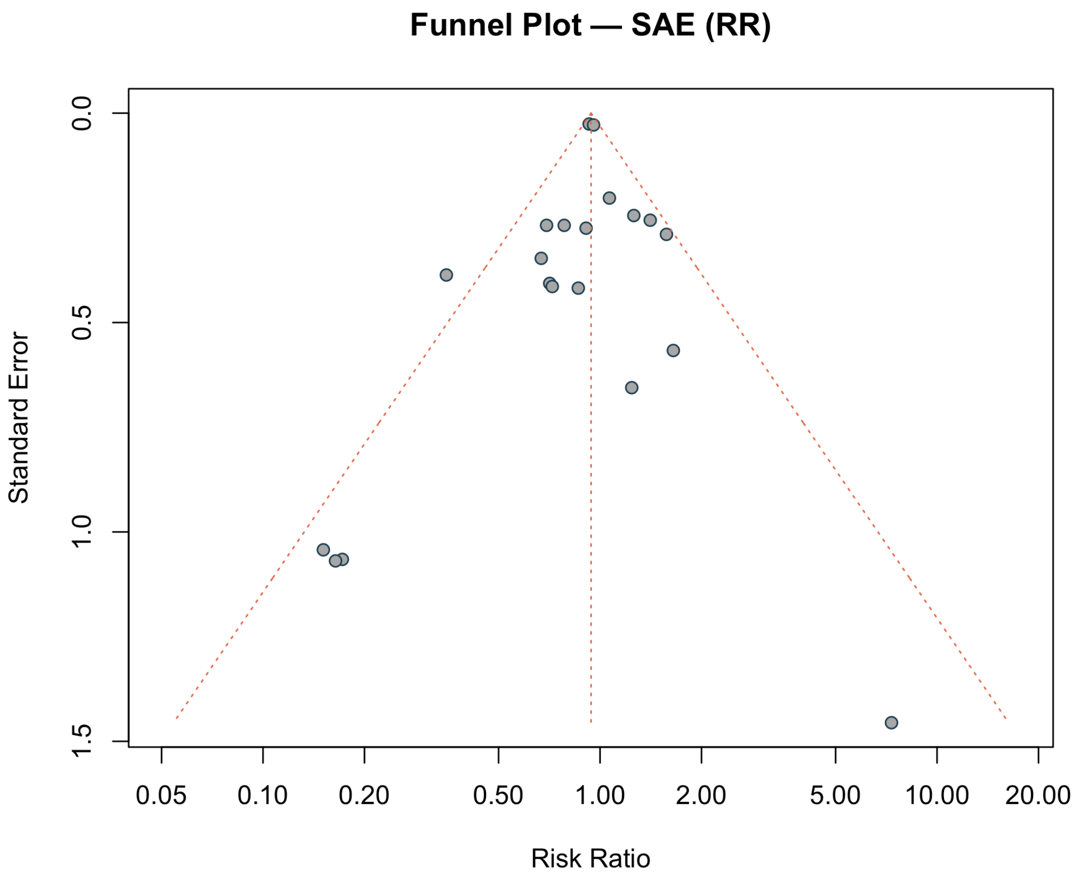

#### eFigure 43: Trim-and-Fill — ACM

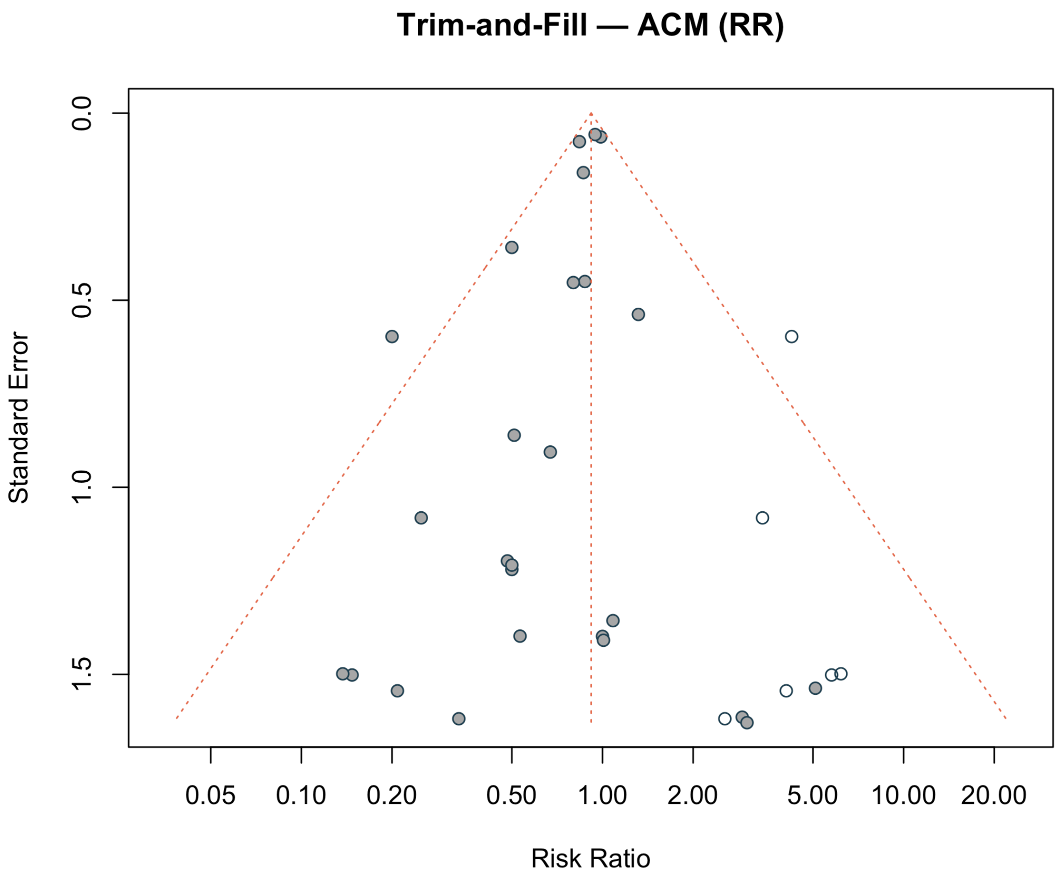

#### eFigure 44: Trim-and-Fill — HFH

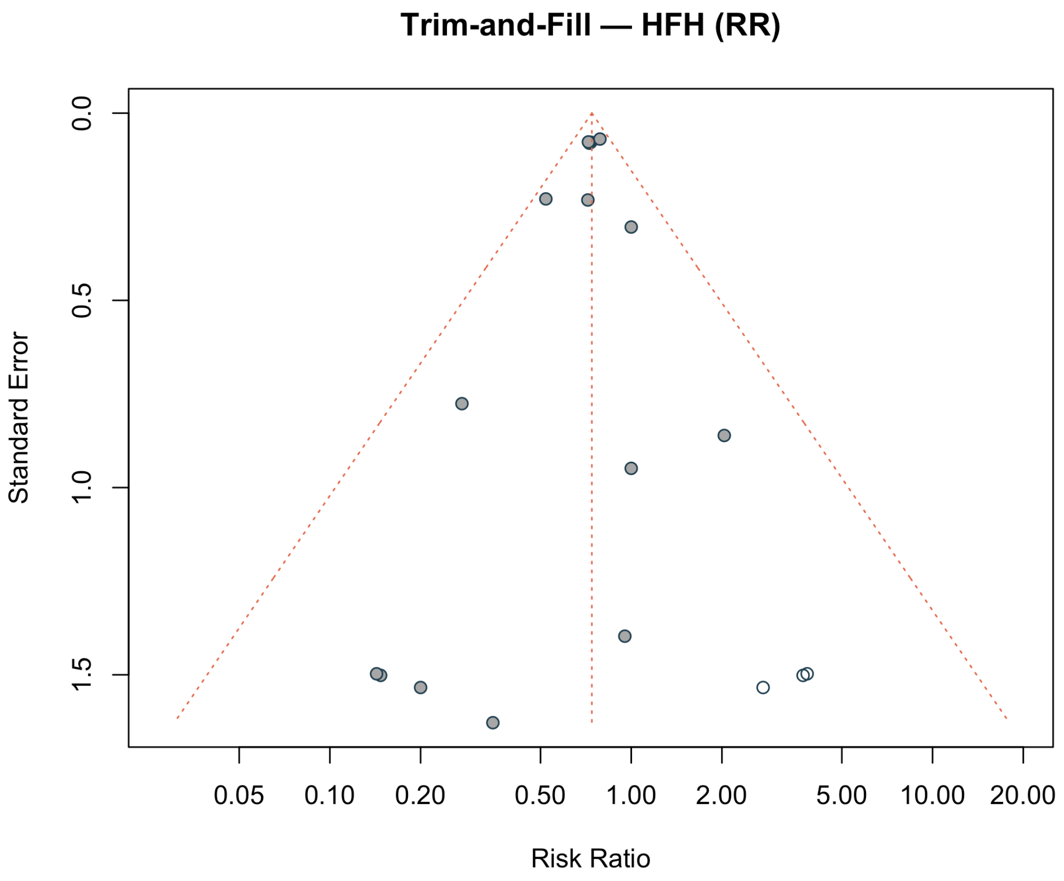

#### eFigure 45: Trim-and-Fill — SAE

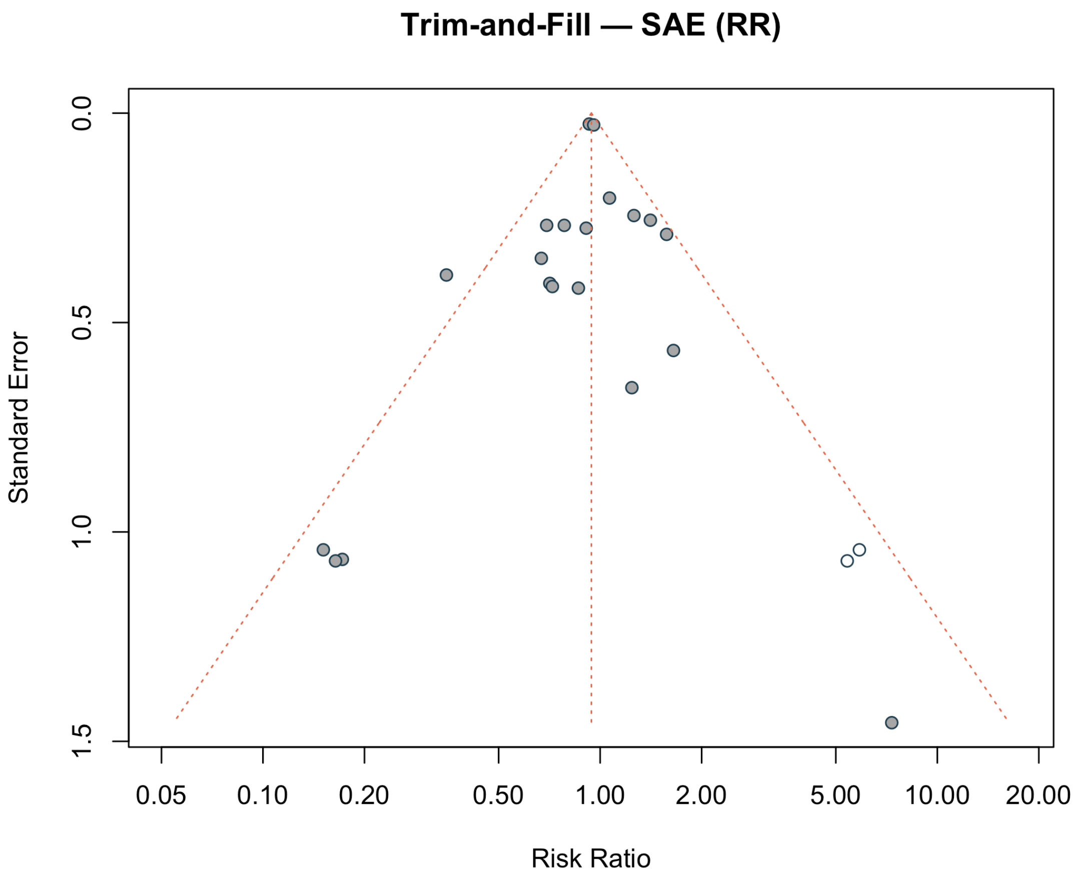

### Sensitivity Analyses

#### eFigure 46: Leave-One-Out — ACM

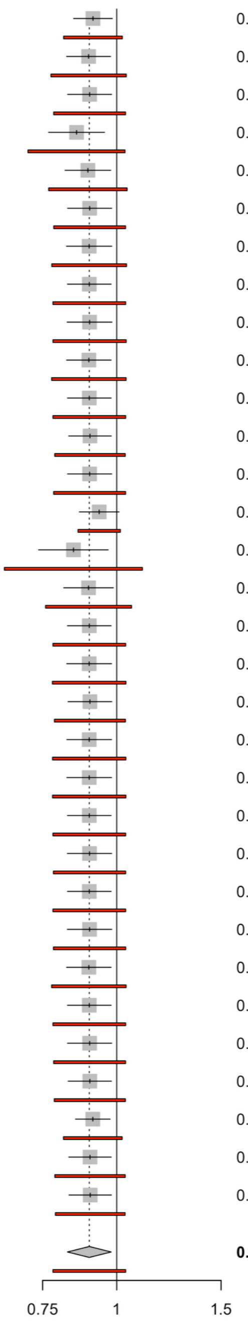


| Study | RR | 95%-CI/PI | P-value | Tau2 | Tau | I2 |
|---|---|---|---|---|---|---|
| Omitting Voors 2022 | 0.91 | [0.85; 0.98]<br>[0.81; 1.02] | 0.0181 | 0.0014 | 0.0374 | 0% |
| Omitting Lee 2021 | 0.90 | [0.82; 0.98]<br>[0.77; 1.04] | 0.0142 | 0.0030 | 0.0546 | 0% |
| Omitting Santos-Gallego 2021 | 0.90 | [0.83; 0.98]<br>[0.78; 1.04] | 0.0178 | 0.0026 | 0.0512 | 0% |
| Omitting Anker 2021 | 0.86 | [0.77; 0.95]<br>[0.71; 1.03] | 0.0068 | 0.0048 | 0.0694 | 0% |
| Omitting Charaya 2023 | 0.89 | [0.82; 0.98]<br>[0.77; 1.04] | 0.0154 | 0.0033 | 0.0575 | 0% |
| Omitting Schulze 2022 | 0.90 | [0.83; 0.98]<br>[0.78; 1.04] | 0.0182 | 0.0026 | 0.0511 | 0% |
| Omitting Abdullaev 2024 | 0.90 | [0.82; 0.98]<br>[0.78; 1.04] | 0.0191 | 0.0029 | 0.0539 | 0% |
| Omitting Ueda 2021 | 0.90 | [0.83; 0.98]<br>[0.78; 1.04] | 0.0158 | 0.0027 | 0.0522 | 0% |
| Omitting Lin 2024 | 0.90 | [0.82; 0.98]<br>[0.78; 1.04] | 0.0196 | 0.0027 | 0.0524 | 0% |
| Omitting Nassif_2020 | 0.90 | [0.82; 0.98]<br>[0.78; 1.04] | 0.0163 | 0.0029 | 0.0538 | 0% |
| Omitting Bhushan_2023 | 0.90 | [0.83; 0.98]<br>[0.78; 1.04] | 0.0158 | 0.0027 | 0.0522 | 0% |
| Omitting EFFORT | 0.90 | [0.83; 0.98]<br>[0.79; 1.03] | 0.0159 | 0.0025 | 0.0498 | 0% |
| Omitting Nassif_2021 | 0.90 | [0.83; 0.98]<br>[0.78; 1.04] | 0.0183 | 0.0026 | 0.0513 | 0% |
| Omitting McMurray_2019 | 0.93 | [0.86; 1.01]<br>[0.86; 1.01] | 0.0813 | 0 | 0 | 0% |
| Omitting Solomon_2022 | 0.85 | [0.74; 0.97]<br>[0.65; 1.11] | 0.0168 | 0.0114 | 0.1069 | 0% |
| Omitting Bhatt_2021 | 0.90 | [0.81; 0.99]<br>[0.76; 1.06] | 0.0278 | 0.0040 | 0.0636 | 0% |
| Omitting Rau_2021 | 0.90 | [0.83; 0.98]<br>[0.78; 1.04] | 0.0158 | 0.0027 | 0.0522 | 0% |
| Omitting Gojaseni 2024 | 0.90 | [0.82; 0.98]<br>[0.78; 1.04] | 0.0180 | 0.0028 | 0.0528 | 0% |
| Omitting Nassif 2021 | 0.90 | [0.83; 0.98]<br>[0.78; 1.03] | 0.0184 | 0.0025 | 0.0503 | 0% |
| Omitting Palau 2022 | 0.90 | [0.82; 0.98]<br>[0.78; 1.04] | 0.0181 | 0.0028 | 0.0526 | 0% |
| Omitting Nassif 2019 | 0.90 | [0.82; 0.98]<br>[0.78; 1.04] | 0.0181 | 0.0028 | 0.0526 | 0% |
| Omitting Jensen 2020 | 0.90 | [0.83; 0.98]<br>[0.78; 1.04] | 0.0158 | 0.0027 | 0.0522 | 0% |
| Omitting de Boer 2020 | 0.90 | [0.83; 0.98]<br>[0.78; 1.04] | 0.0183 | 0.0027 | 0.0516 | 0% |
| Omitting Ovchinnikov 2021 | 0.90 | [0.83; 0.98]<br>[0.78; 1.04] | 0.0158 | 0.0027 | 0.0522 | 0% |
| Omitting NCT03448419 (EMPERIAL-Reduced) | 0.90 | [0.83; 0.98]<br>[0.78; 1.04] | 0.0186 | 0.0027 | 0.0516 | 0% |
| Omitting Abraham 2021 | 0.90 | [0.82; 0.98]<br>[0.78; 1.04] | 0.0162 | 0.0029 | 0.0538 | 0% |
| Omitting Reis 2022 | 0.90 | [0.83; 0.98]<br>[0.78; 1.04] | 0.0158 | 0.0027 | 0.0522 | 0% |
| Omitting Mocan 2025 | 0.90 | [0.83; 0.98]<br>[0.78; 1.04] | 0.0183 | 0.0026 | 0.0512 | 0% |
| Omitting Tanaka 2020 | 0.90 | [0.83; 0.98]<br>[0.78; 1.03] | 0.0169 | 0.0025 | 0.0505 | 0% |
| Omitting Golubovskaya 2025a | 0.91 | [0.85; 0.97]<br>[0.81; 1.02] | 0.0088 | 0.0014 | 0.0375 | 0% |
| Omitting Zhou 2024 | 0.90 | [0.83; 0.98]<br>[0.79; 1.03] | 0.0156 | 0.0025 | 0.0497 | 0% |
| Omitting Shoshina 2026 | 0.90 | [0.83; 0.98]<br>[0.79; 1.03] | 0.0164 | 0.0024 | 0.0490 | 0% |
| **Random effects model** | **0.90** | **[0.83; 0.98]** | **0.0158** | **0.0027** | **0.0522** | **0%** |
| **Prediction interval** | | **[0.78; 1.04]** | | | | |

### eFigure 47: Leave-One-Out — HFH

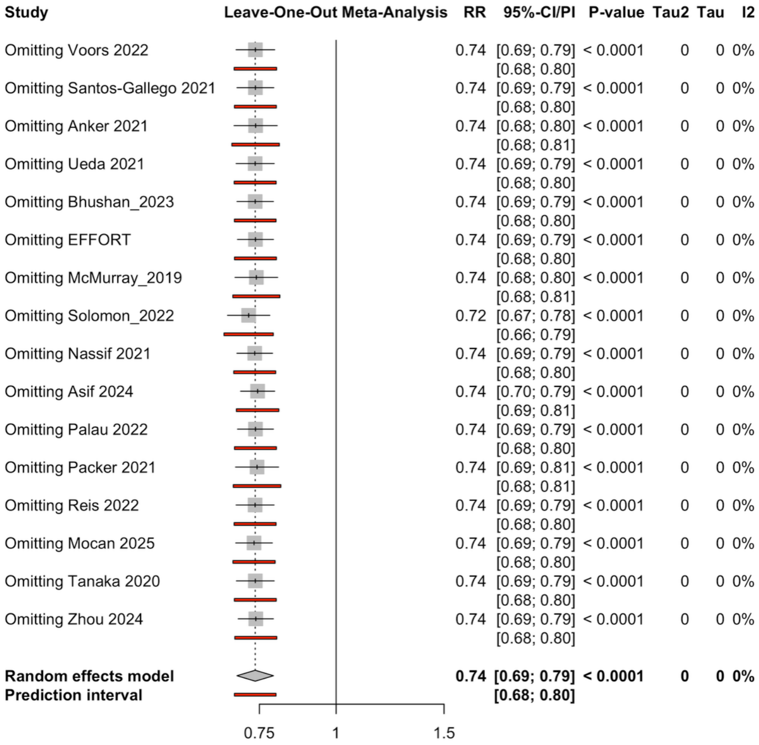

### eFigure 48: Influence Diagnostics — ACM

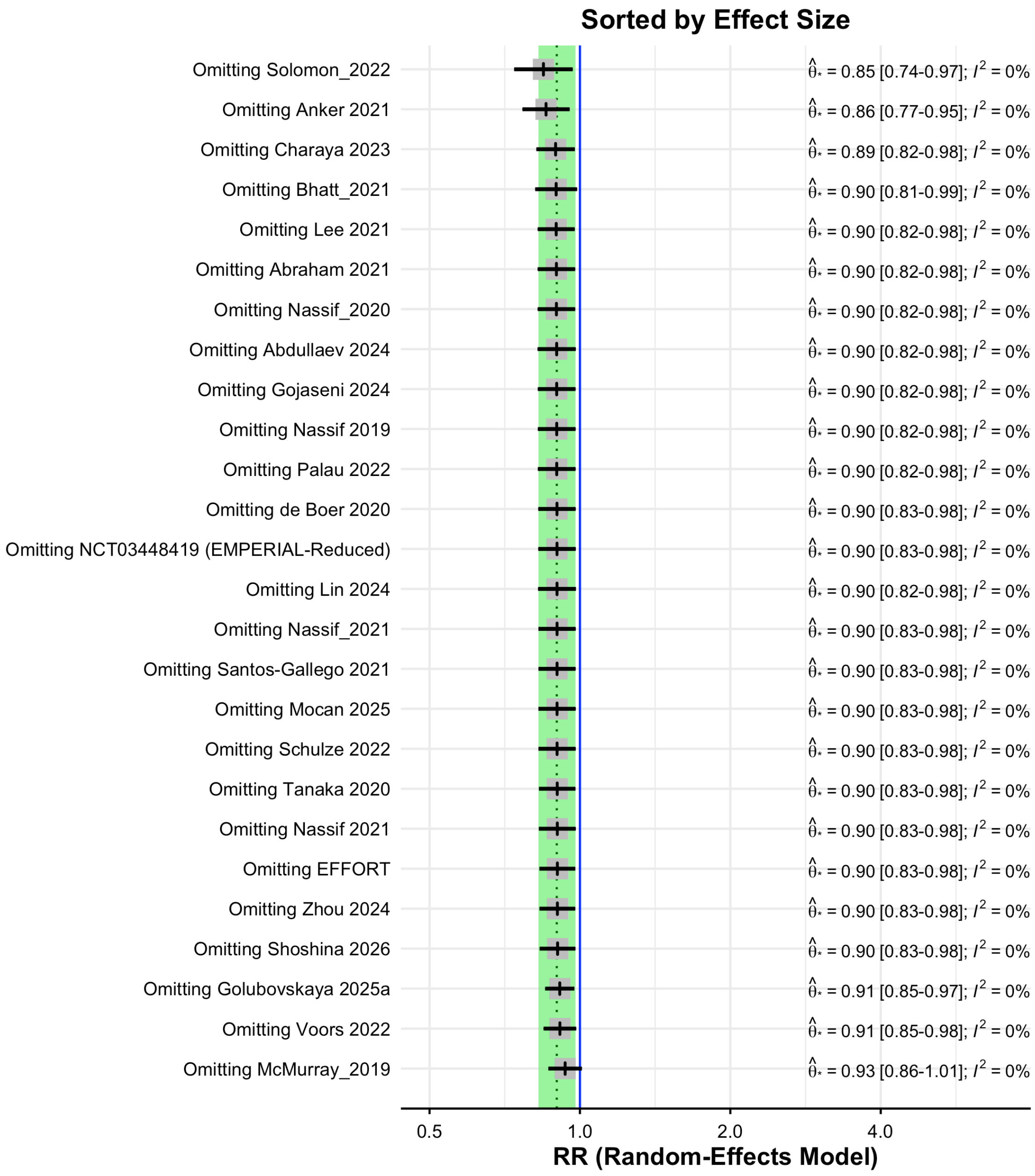

#### eFigure 49: Influence Diagnostics — HFH

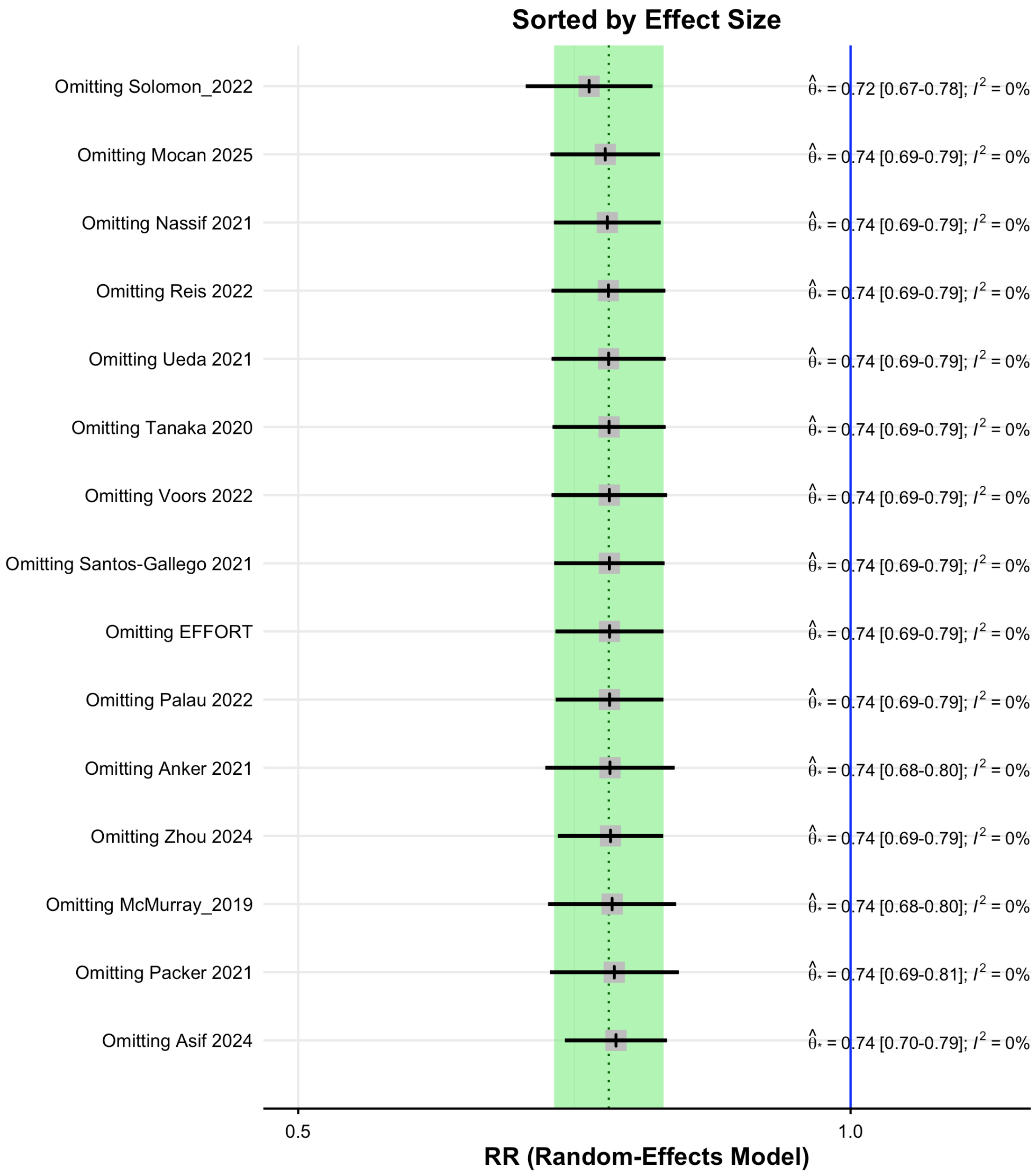

### Heterogeneity

#### eFigure 50: Baujat Plot — ACM

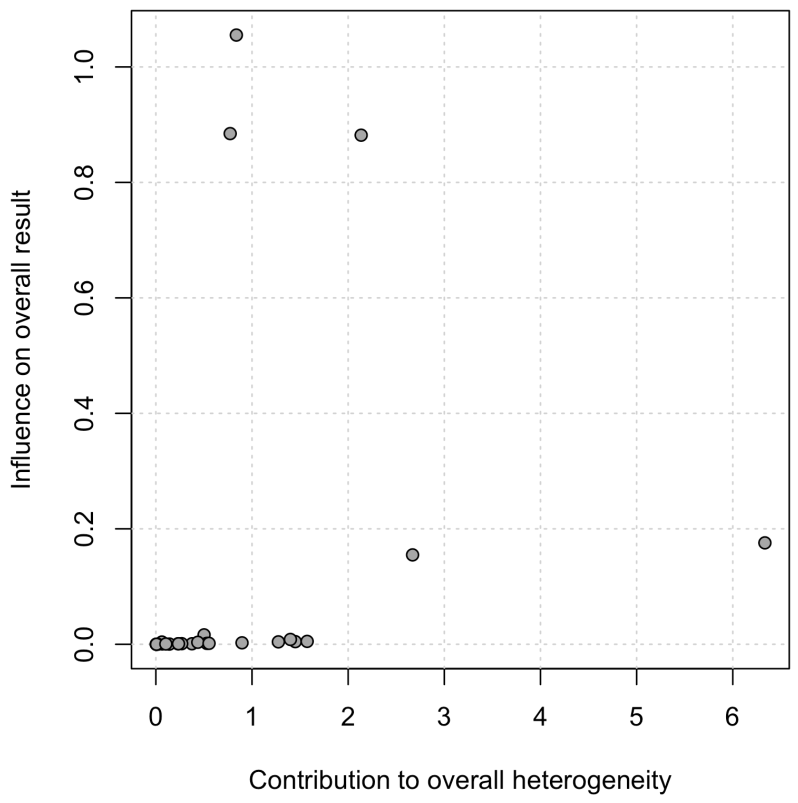

#### eFigure 51: Baujat Plot — HFH

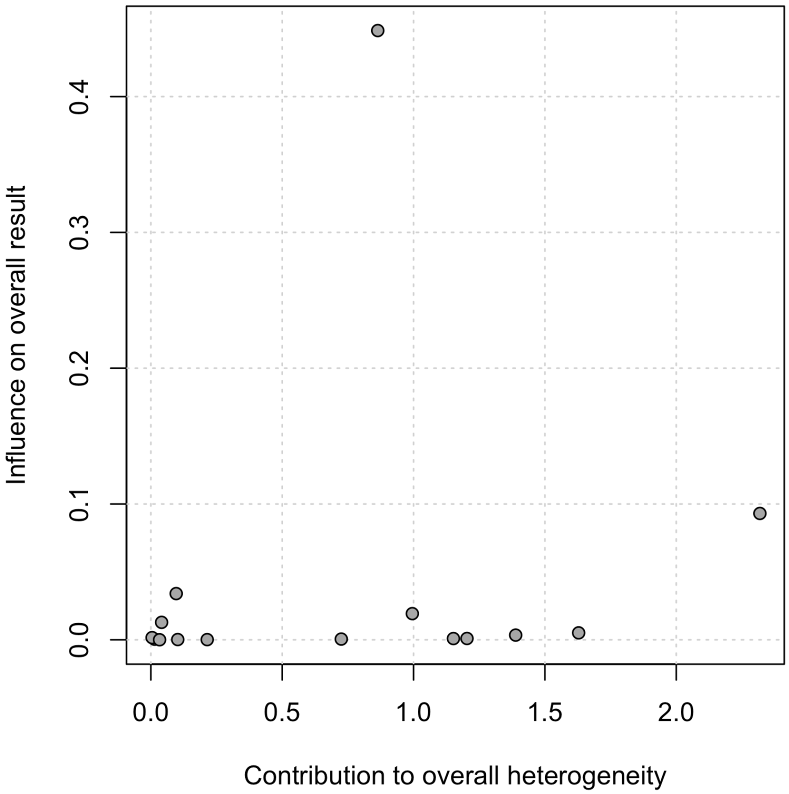

### Cumulative Meta-Analysis

#### eFigure 52: Cumulative — ACM

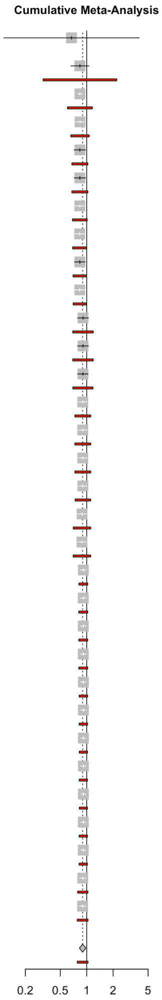

| Study | RR | 95%-CI/PI | P-value | Tau2 | Tau | I2 |
|---|---|---|---|---|---|---|
| Adding NCT03448419 (EMPERIAL-Reduced) (k=1) | 0.67 | [0.11; 3.96] | 0.6596 | . | . | . |
| Adding McMurray_2019 (k=2) | 0.84 | [0.66; 1.06]<br>[0.32; 2.20] | 0.0663 | 0 | 0 | 0% |
| Adding Nassif 2019 (k=3) | 0.84 | [0.79; 0.89]<br>[0.60; 1.16] | 0.0070 | 0 | 0 | 0% |
| Adding Nassif_2020 (k=4) | 0.84 | [0.75; 0.94]<br>[0.66; 1.07] | 0.0165 | 0 | 0 | 0% |
| Adding EFFORT (k=5) | 0.84 | [0.72; 0.97]<br>[0.68; 1.03] | 0.0289 | 0 | 0 | 0% |
| Adding Jensen 2020 (k=6) | 0.84 | [0.72; 0.97]<br>[0.68; 1.03] | 0.0289 | 0 | 0 | 0% |
| Adding de Boer 2020 (k=7) | 0.83 | [0.74; 0.95]<br>[0.69; 1.01] | 0.0144 | 0 | 0 | 0% |
| Adding Tanaka 2020 (k=8) | 0.83 | [0.73; 0.95]<br>[0.69; 1.00] | 0.0130 | 0 | 0 | 0% |
| Adding Lee 2021 (k=9) | 0.84 | [0.73; 0.96]<br>[0.70; 1.00] | 0.0190 | 0 | 0 | 0% |
| Adding Santos-Gallego 2021 (k=10) | 0.83 | [0.73; 0.95]<br>[0.70; 0.99] | 0.0133 | 0 | 0 | 0% |
| Adding Anker 2021 (k=11) | 0.91 | [0.79; 1.05]<br>[0.69; 1.19] | 0.1597 | 0.0079 | 0.0889 | 0% |
| Adding Ueda 2021 (k=12) | 0.91 | [0.79; 1.05]<br>[0.69; 1.19] | 0.1597 | 0.0079 | 0.0889 | 0% |
| Adding Nassif_2021 (k=13) | 0.91 | [0.79; 1.04]<br>[0.69; 1.18] | 0.1347 | 0.0079 | 0.0890 | 0% |
| Adding Bhatt_2021 (k=14) | 0.90 | [0.81; 1.01]<br>[0.73; 1.11] | 0.0607 | 0.0049 | 0.0696 | 0% |
| Adding Rau_2021 (k=15) | 0.90 | [0.81; 1.01]<br>[0.73; 1.11] | 0.0607 | 0.0049 | 0.0696 | 0% |
| Adding Ovchinnikov 2021 (k=16) | 0.90 | [0.81; 1.01]<br>[0.73; 1.11] | 0.0607 | 0.0049 | 0.0696 | 0% |
| Adding Abraham 2021 (k=17) | 0.90 | [0.81; 1.01]<br>[0.74; 1.11] | 0.0614 | 0.0047 | 0.0689 | 0% |
| Adding Voors 2022 (k=18) | 0.88 | [0.78; 1.00]<br>[0.70; 1.11] | 0.0508 | 0.0067 | 0.0818 | 0% |
| Adding Schulze 2022 (k=19) | 0.88 | [0.78; 0.99]<br>[0.70; 1.11] | 0.0419 | 0.0068 | 0.0826 | 0% |
| Adding Solomon_2022 (k=20) | 0.92 | [0.85; 0.99]<br>[0.81; 1.03] | 0.0247 | 0.0013 | 0.0366 | 0% |
| Adding Nassif 2021 (k=21) | 0.91 | [0.85; 0.98]<br>[0.81; 1.03] | 0.0209 | 0.0014 | 0.0379 | 0% |
| Adding Palau 2022 (k=22) | 0.91 | [0.85; 0.98]<br>[0.81; 1.03] | 0.0172 | 0.0014 | 0.0376 | 0% |
| Adding Reis 2022 (k=23) | 0.91 | [0.85; 0.98]<br>[0.81; 1.03] | 0.0172 | 0.0014 | 0.0376 | 0% |
| Adding Charaya 2023 (k=24) | 0.92 | [0.86; 0.98]<br>[0.82; 1.02] | 0.0171 | 0.0012 | 0.0344 | 0% |
| Adding Bhushan_2023 (k=25) | 0.92 | [0.86; 0.98]<br>[0.82; 1.02] | 0.0171 | 0.0012 | 0.0344 | 0% |
| Adding Abdullaev 2024 (k=26) | 0.92 | [0.86; 0.98]<br>[0.82; 1.02] | 0.0134 | 0.0011 | 0.0332 | 0% |
| Adding Lin 2024 (k=27) | 0.92 | [0.86; 0.98]<br>[0.82; 1.02] | 0.0103 | 0.0011 | 0.0330 | 0% |
| Adding Gojaseni 2024 (k=28) | 0.92 | [0.86; 0.98]<br>[0.82; 1.02] | 0.0086 | 0.0011 | 0.0327 | 0% |
| Adding Zhou 2024 (k=29) | 0.91 | [0.86; 0.98]<br>[0.82; 1.02] | 0.0099 | 0.0012 | 0.0344 | 0% |
| Adding Mocan 2025 (k=30) | 0.91 | [0.86; 0.97]<br>[0.82; 1.02] | 0.0084 | 0.0012 | 0.0351 | 0% |
| Adding Golubovskaya 2025a (k=31) | 0.90 | [0.83; 0.98]<br>[0.79; 1.03] | 0.0164 | 0.0024 | 0.0490 | 0% |
| Adding Shoshina 2026 (k=32) | 0.90 | [0.83; 0.98]<br>[0.78; 1.04] | 0.0158 | 0.0027 | 0.0522 | 0% |
| **Random effects model** | **0.90** | **[0.83; 0.98]** | **0.0158** | **0.0027** | **0.0522** | **0%** |
| **Prediction interval** | | **[0.78; 1.04]** | | | | |

### eFigure 53: Cumulative — HFH

| Study | RR | 95%-CI/PI | P-value | Tau2 | Tau | I2 |
|---|---|---|---|---|---|---|
| Adding McMurray_2019 (k=1) | 0.73 | [0.62; 0.85] | < 0.0001 | . | . | . |
| Adding EFFORT (k=2) | 0.66 | [0.01; 76.31] | 0.4685 | 0.1418 | 0.3766 | 11.1% |
| | | [0.00; 559.79] | | | | |
| Adding Tanaka 2020 (k=3) | 0.72 | [0.55; 0.96] | 0.0378 | 0 | 0 | 0% |
| | | [0.51; 1.02] | | | | |
| Adding Santos-Gallego 2021 (k=4) | 0.61 | [0.26; 1.40] | 0.1530 | 0.1367 | 0.3697 | 0% |
| | | [0.12; 3.07] | | | | |
| Adding Anker 2021 (k=5) | 0.73 | [0.65; 0.81] | 0.0013 | 0 | 0 | 0% |
| | | [0.62; 0.85] | | | | |
| Adding Ueda 2021 (k=6) | 0.73 | [0.66; 0.80] | 0.0003 | 0 | 0 | 0% |
| | | [0.63; 0.84] | | | | |
| Adding Packer 2021 (k=7) | 0.73 | [0.68; 0.77] | < 0.0001 | 0 | 0 | 0% |
| | | [0.65; 0.81] | | | | |
| Adding Voors 2022 (k=8) | 0.72 | [0.68; 0.77] | < 0.0001 | 0 | 0 | 0% |
| | | [0.65; 0.80] | | | | |
| Adding Solomon_2022 (k=9) | 0.74 | [0.70; 0.78] | < 0.0001 | 0 | 0 | 0% |
| | | [0.68; 0.81] | | | | |
| Adding Nassif 2021 (k=10) | 0.74 | [0.70; 0.79] | < 0.0001 | 0 | 0 | 0% |
| | | [0.68; 0.81] | | | | |
| Adding Palau 2022 (k=11) | 0.74 | [0.70; 0.79] | < 0.0001 | 0 | 0 | 0% |
| | | [0.68; 0.81] | | | | |
| Adding Reis 2022 (k=12) | 0.74 | [0.70; 0.79] | < 0.0001 | 0 | 0 | 0% |
| | | [0.68; 0.81] | | | | |
| Adding Bhushan_2023 (k=13) | 0.74 | [0.70; 0.79] | < 0.0001 | 0 | 0 | 0% |
| | | [0.68; 0.81] | | | | |
| Adding Asif 2024 (k=14) | 0.74 | [0.69; 0.79] | < 0.0001 | 0 | 0 | 0% |
| | | [0.68; 0.80] | | | | |
| Adding Zhou 2024 (k=15) | 0.74 | [0.69; 0.79] | < 0.0001 | 0 | 0 | 0% |
| | | [0.68; 0.80] | | | | |
| Adding Mocan 2025 (k=16) | 0.74 | [0.69; 0.79] | < 0.0001 | 0 | 0 | 0% |
| | | [0.68; 0.80] | | | | |
| **Random effects model** | **0.74** | **[0.69; 0.79]** | **< 0.0001** | **0** | **0** | **0%** |
| **Prediction interval** | | **[0.68; 0.80]** | | | | |

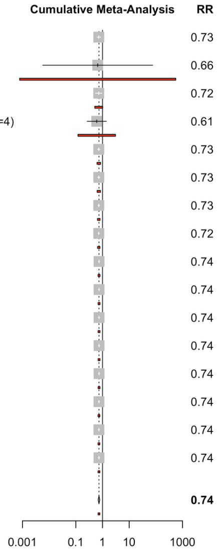

### eFigure 54: Cumulative — CVD

| Study | RR | 95%-CI/PI | P-value | Tau2 | Tau | I2 |
|---|---|---|---|---|---|---|
| Adding McMurray_2019 (k=1) | 0.83 | [0.70; 0.98] | 0.0292 | . | . | . |
| Adding Nassif 2019 (k=2) | 0.83 | [0.72; 0.96]<br>[0.28; 2.44] | 0.0399 | 0 | 0 | 0% |
| Adding EFFORT (k=3) | 0.83 | [0.65; 1.05]<br>[0.57; 1.19] | 0.0748 | 0 | 0 | 0% |
| Adding Anker 2021 (k=4) | 0.86 | [0.76; 0.97]<br>[0.71; 1.05] | 0.0314 | 0 | 0 | 0% |
| Adding Ueda 2021 (k=5) | 0.86 | [0.76; 0.97]<br>[0.71; 1.05] | 0.0314 | 0 | 0 | 0% |
| Adding Nassif_2021 (k=6) | 0.86 | [0.76; 0.97]<br>[0.71; 1.05] | 0.0314 | 0 | 0 | 0% |
| Adding Bhatt_2021 (k=7) | 0.86 | [0.79; 0.94]<br>[0.73; 1.01] | 0.0108 | 0 | 0 | 0% |
| Adding Solomon_2022 (k=8) | 0.87 | [0.82; 0.93]<br>[0.77; 0.98] | 0.0025 | 0 | 0 | 0% |
| Adding Pastore 2024 (k=9) | 0.87 | [0.82; 0.93]<br>[0.77; 0.98] | 0.0025 | 0 | 0 | 0% |
| Adding Golubovskaya 2025a (k=10) | 0.86 | [0.76; 0.98]<br>[0.77; 0.97] | 0.0311 | 0 | 0 | 18.4% |
| **Random effects model** | **0.86** | **[0.76; 0.98]** | **0.0311** | **0** | **0** | **18.4%** |
| **Prediction interval** | | **[0.77; 0.97]** | | | | |

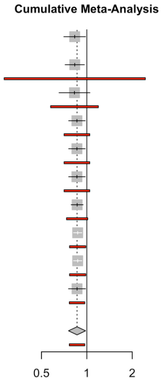

### eFigure 55: Cumulative — Composite

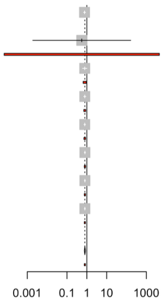


| Study | RR | 95%-CI/PI | P-value | Tau2 | Tau | I2 |
|---|---|---|---|---|---|---|
| Adding McMurray_2019 (k=1) | 0.77 | [0.68; 0.87] | < 0.0001 | . | . | . |
| Adding Damman 2020 (k=2) | 0.55 | [0.00; 163.13] | 0.4135 | 0.3021 | 0.5497 | 68.3% |
| | | [0.00; 4516.86] | | | | |
| Adding Anker 2021 (k=3) | 0.78 | [0.61; 1.01] | 0.0539 | < 0.0001 | 0.0025 | 46.6% |
| | | [0.65; 0.94] | | | | |
| Adding Packer 2021 (k=4) | 0.79 | [0.70; 0.89] | 0.0088 | < 0.0001 | 0.0014 | 21.0% |
| | | [0.71; 0.88] | | | | |
| Adding Voors 2022 (k=5) | 0.79 | [0.71; 0.87] | 0.0023 | < 0.0001 | 0.0012 | 4.2% |
| | | [0.71; 0.86] | | | | |
| Adding Solomon_2022 (k=6) | 0.80 | [0.74; 0.86] | 0.0007 | < 0.0001 | 0.0008 | 4.6% |
| | | [0.74; 0.86] | | | | |
| Adding Lin 2024 (k=7) | 0.80 | [0.74; 0.86] | 0.0002 | < 0.0001 | 0.0016 | 0% |
| | | [0.74; 0.86] | | | | |
| Adding Asif 2024 (k=8) | 0.80 | [0.75; 0.85] | < 0.0001 | < 0.0001 | 0.0006 | 0% |
| | | [0.74; 0.85] | | | | |
| **Random effects model** | **0.80** | **[0.75; 0.85]** | **< 0.0001** | **< 0.0001** | **0.0006** | **0%** |
| **Prediction interval** | | **[0.74; 0.85]** | | | | |

### eFigure 56: Cumulative — SAE

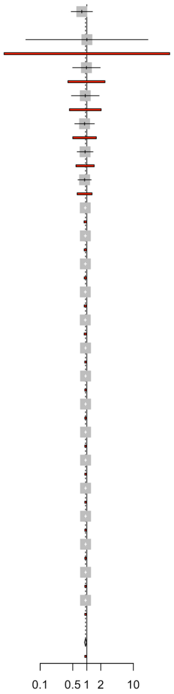


| Study | RR | 95%-CI/PI | P-value | Tau2 | Tau | I2 |
|---|---|---|---|---|---|---|
| Adding NCT03448419 (EMPERIAL-Reduced) (k=1) | 0.78 | [0.46; 1.32] | 0.3608 | . | . | . |
| Adding Nassif 2019 (k=2) | 1.01 | [0.05; 20.55] / [0.02; 60.20] | 0.9851 | 0.0473 | 0.2175 | 41.8% |
| Adding Nassif_2020 (k=3) | 0.98 | [0.50; 1.94] / [0.39; 2.46] | 0.9266 | 0.0129 | 0.1135 | 0% |
| Adding EFFORT (k=4) | 0.93 | [0.47; 1.84] / [0.42; 2.03] | 0.7535 | 0.0249 | 0.1579 | 33.0% |
| Adding Damman 2020 (k=5) | 0.90 | [0.55; 1.46] / [0.50; 1.60] | 0.5655 | 0.0162 | 0.1273 | 18.7% |
| Adding de Boer 2020 (k=6) | 0.92 | [0.62; 1.36] / [0.59; 1.44] | 0.6079 | 0.0070 | 0.0840 | 2.6% |
| Adding Tanaka 2020 (k=7) | 0.90 | [0.65; 1.24] / [0.62; 1.30] | 0.4495 | 0.0026 | 0.0511 | 0% |
| Adding Anker 2021 (k=8) | 0.93 | [0.88; 0.98] / [0.87; 0.99] | 0.0128 | < 0.0001 | 0.0010 | 0% |
| Adding Nassif_2021 (k=9) | 0.93 | [0.88; 0.99] / [0.88; 0.99] | 0.0243 | < 0.0001 | 0.0003 | 1.9% |
| Adding Rau_2021 (k=10) | 0.93 | [0.88; 0.99] / [0.88; 0.99] | 0.0233 | < 0.0001 | 0.0004 | 1.8% |
| Adding Abraham 2021 (k=11) | 0.93 | [0.88; 0.98] / [0.88; 0.98] | 0.0182 | < 0.0001 | 0.0006 | 3.6% |
| Adding Schulze 2022 (k=12) | 0.93 | [0.88; 0.98] / [0.88; 0.98] | 0.0144 | < 0.0001 | 0.0021 | 2.4% |
| Adding Solomon_2022 (k=13) | 0.94 | [0.90; 0.98] / [0.90; 0.98] | 0.0070 | < 0.0001 | 0.0016 | 0% |
| Adding Nassif 2021 (k=14) | 0.94 | [0.90; 0.99] / [0.90; 0.98] | 0.0124 | < 0.0001 | 0.0024 | 13.6% |
| Adding Akasaka 2022 (k=15) | 0.94 | [0.90; 0.99] / [0.91; 0.98] | 0.0140 | < 0.0001 | 0.0014 | 21.0% |
| Adding Borlaug 2023 (k=16) | 0.94 | [0.90; 0.99] / [0.91; 0.98] | 0.0152 | < 0.0001 | 0.0010 | 23.9% |
| Adding Bhushan_2023 (k=17) | 0.94 | [0.90; 0.99] / [0.91; 0.98] | 0.0152 | < 0.0001 | 0.0010 | 23.9% |
| Adding Hundertmark 2023 (k=18) | 0.94 | [0.90; 0.99] / [0.91; 0.98] | 0.0174 | < 0.0001 | 0.0022 | 29.8% |
| Adding McMurray 2024 (k=19) | 0.94 | [0.90; 0.99] / [0.91; 0.98] | 0.0159 | < 0.0001 | 0.0018 | 26.6% |
| Adding Lin 2024 (k=20) | 0.94 | [0.90; 0.99] / [0.91; 0.98] | 0.0125 | < 0.0001 | 0.0003 | 22.3% |
| Adding Xie 2024 (k=21) | 0.94 | [0.90; 0.99] / [0.91; 0.98] | 0.0125 | < 0.0001 | 0.0003 | 22.3% |
| Adding Golubovskaya 2025a (k=22) | 0.94 | [0.90; 0.99] / [0.90; 0.98] | 0.0174 | < 0.0001 | 0.0028 | 36.1% |
| **Random effects model** | **0.94** | **[0.90; 0.99]** | **0.0174** | **< 0.0001** | **0.0028** | **36.1%** |
| **Prediction interval** | | **[0.90; 0.98]** | | | | |

### Additional Funnel Plots

#### eFigure 57: Funnel Plot — NT-proBNP

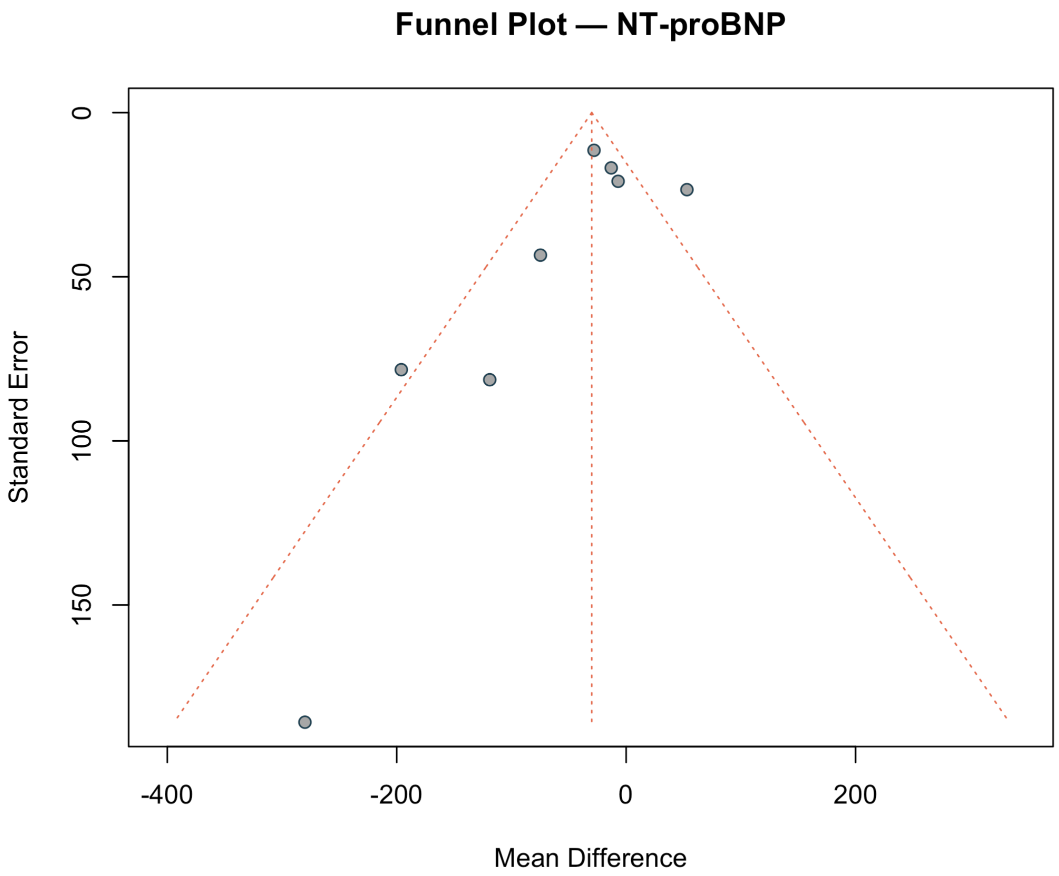

#### eFigure 58: Funnel Plot — eGFR

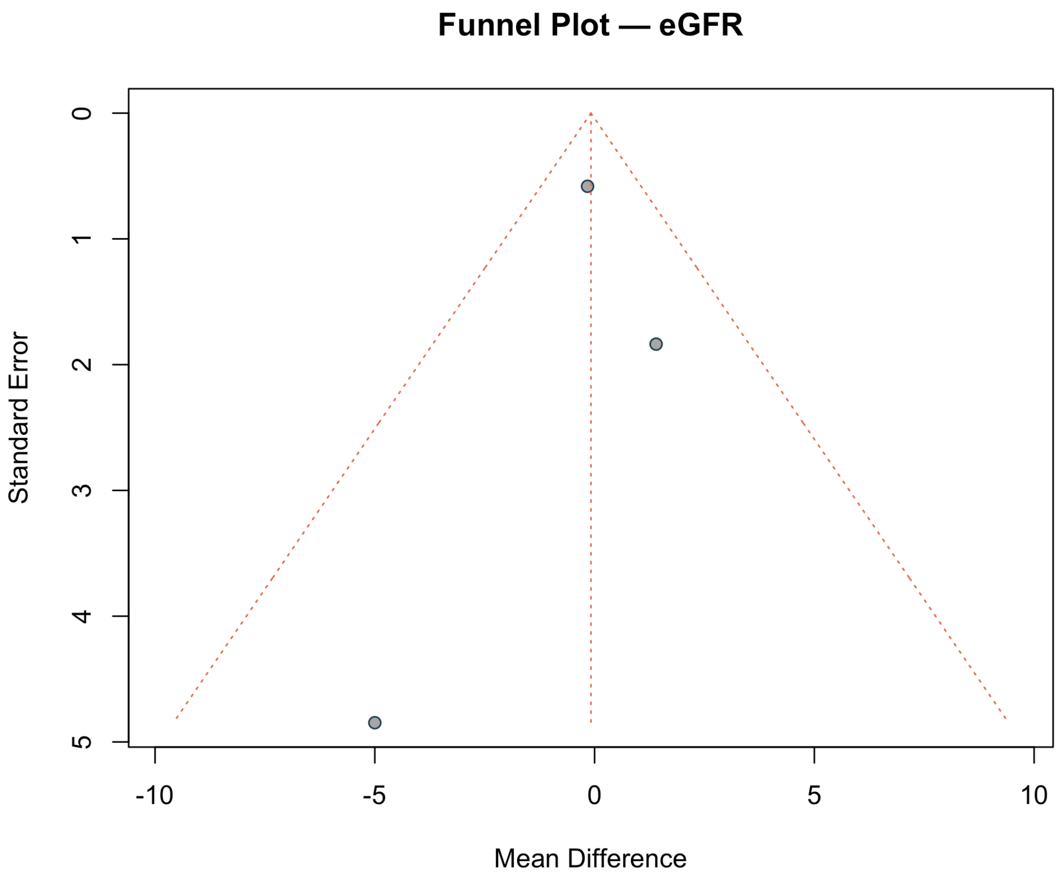

#### eFigure 59: Funnel Plot — SBP

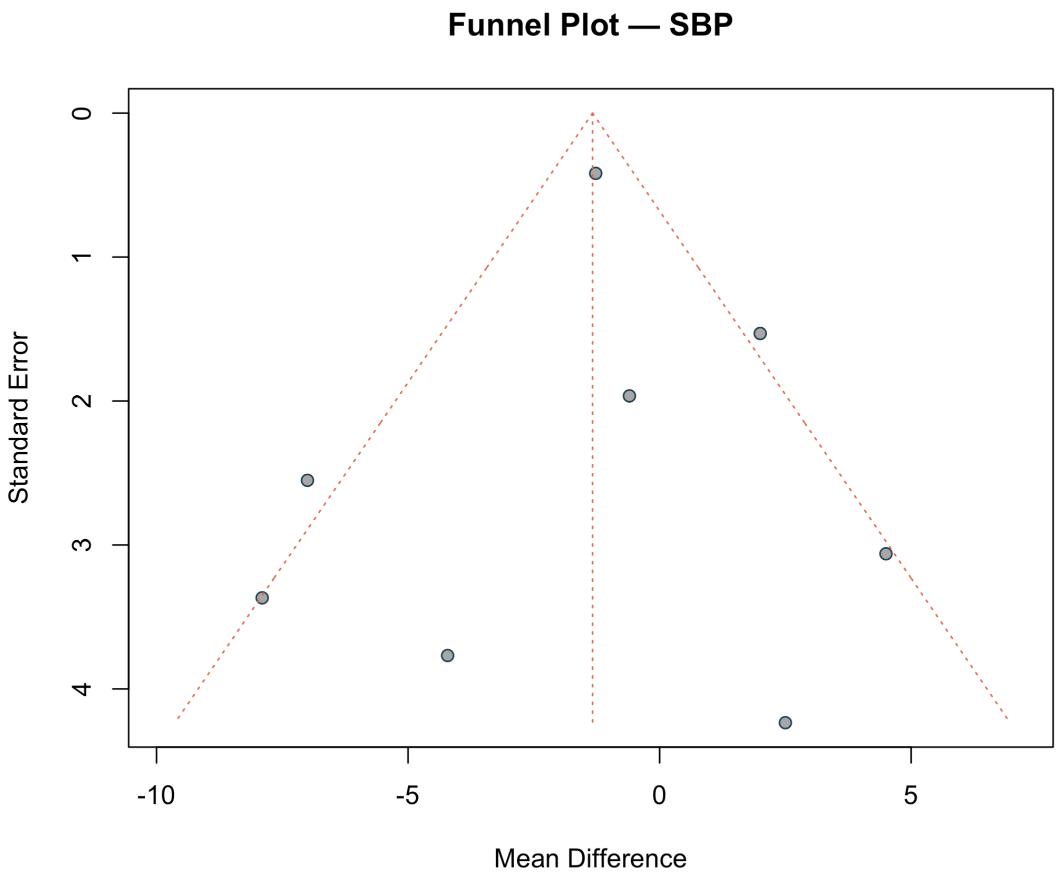
